# Lorazepam stimulates IL-6 production and is associated with poor survival outcomes in pancreatic cancer

**DOI:** 10.1101/2023.03.01.23286581

**Authors:** Abigail C. Cornwell, Arwen A. Tisdale, Swati Venkat, Kathryn E. Maraszek, Abdulrahman A. Alahmari, Anthony George, Kristopher Attwood, Madison George, Donald Rempinski, Janusz Franco-Barraza, Mark D. Parker, Eduardo Cortes Gomez, Christos Fountzilas, Edna Cukierman, Nina G. Steele, Michael E. Feigin

## Abstract

**Purpose:** This research investigates the association between benzodiazepines (BZDs) and cancer patient survival outcomes. Due to the high prevalence of BZD use in pancreatic cancer patients, we evaluated the effect of commonly prescribed BZDs on the pancreatic cancer tumor microenvironment and cancer-associated fibroblast (CAF) signaling.

**Experimental Design:** Multivariate Cox regression modeling was used to retrospectively measure associations between Roswell Park cancer patient survival outcomes and BZD prescription records. Immunohistochemistry, H&E, Masson’s trichrome, *in situ* hybridization, and RNA sequencing were used to evaluate the impact of lorazepam (LOR) on the PDAC tumor microenvironment, using murine pancreatic cancer models. ELISA and qPCR were used to determine the impact of BZDs on IL-6 expression/secretion by human immortalized pancreatic CAFs. PRESTO-Tango assays, reanalysis of PDAC single cell sequencing/TCGA datasets, and GPR68 CRISPRi knockdown CAF cells were used to mechanistically determine the impact of BZDs on CAF-specific GPR68 signaling.

**Results:** LOR is associated with worse progression-free survival (PFS) while alprazolam (ALP) is associated with improved PFS, in pancreatic cancer patients receiving chemotherapy. LOR promotes desmoplasia (fibrosis and extracellular matrix protein deposition), inflammatory signaling, IL-6 expression/secretion in CAFs, and ischemic necrosis. LOR promotes inflammatory signaling and IL-6 secretion by CAFs through activation of GPR68. GPR68 is preferentially expressed on human PDAC CAFs, and n-unsubstituted BZDs significantly increase GPR68 activation under acidic conditions. LOR increases IL-6 expression and secretion in CAFs in a pH and GPR68-dependent manner. Conversely, ALP, and other GPR68 non-activator BZDs decrease IL-6 in human CAFs in a pH and GPR68-independent manner. Across many cancer types, LOR is associated with worse survival outcomes relative to ALP and patients not receiving BZDs.

**Conclusion:** We demonstrate that LOR stimulates fibrosis and inflammatory signaling, promotes ischemic necrosis, and is associated with decreased pancreatic cancer patient survival.

## Introduction

Pancreatic cancer is a recalcitrant disease with the poorest five-year survival rate (12%) relative to all cancers assessed by the American Cancer Society from 2012-2018 (1). In the United States, pancreatic cancer is projected to be the second leading cause of cancer-related death by 2030, despite accounting for only ∼3% of all estimated new cancer cases (2). Over 90% of patients with pancreatic cancer present with pancreatic adenocarcinoma (PDAC), which is associated with the worst clinical outcomes (3). This disease is often lethal because patients present with non-specific symptoms such as weight loss, abdominal pain, and fatigue, and are consequently diagnosed at late stages. Complete surgical resection is the only curative therapy. However, at diagnosis only 20% of patients are surgical candidates (4).

A unique feature further driving this deadly disease is the presence of a dense, desmoplastic (fibrotic) stroma that impedes drug delivery. The PDAC tumor microenvironment (TME), which is composed of cancer-associated fibroblasts (CAFs), immune cells, and extracellular matrix (ECM) proteins, can comprise up to 90% of the tumor volume and plays important roles in PDAC development, progression, and therapeutic resistance (5). CAFs are plastic, highly heterogeneous cells, with both tumor-promoting and tumor-restraining roles (6). The two most well-characterized CAF subtypes are myofibroblastic CAFs (myCAFs) and inflammatory CAFs (iCAFs) (7). myCAFs preferentially express α-SMA and are thought to be tumor restraining. iCAFs secrete high levels of inflammatory cytokines, most notably interleukin-6 (IL-6), and are thought to be pro-tumorigenic due to the fact IL-6 is associated with worse survival outcomes (8). CAFs influence tumor cell growth, angiogenesis, metastasis, ECM remodeling, and immune cell signaling and function by secreting ECM proteins, growth factors, chemokines, and cytokines (6). Therefore, understanding how CAFs develop, undergo subtype switching, and interact with tumor and immune cells, subsequently modulating therapy response, is fundamental to improving PDAC patient survival.

The role of palliative care medicine in influencing the TME and cancer patient outcomes is also vitally important. Cancer is a devastating diagnosis, associated with emotional distress, anxiety, and depression (9). Harsh surgical, radiological, and chemotherapeutic interventions can induce numerous side-effects, including nausea, anxiety, fatigue, and insomnia (10). To combat these cancer-associated effects, patients are frequently prescribed an array of palliative care drugs such as aspirin, cannabinoids, antihistamines, selective serotonin reuptake inhibitors (SSRIs), opioids, and benzodiazepines (BZDs). There is a growing appreciation that many commonly prescribed drugs can either positively or negatively impact cancer risk, tumor progression, and chemotherapeutic efficacy (11). Many of these interactions are being tested experimentally, providing insight into clinical observations, and opening new avenues to improve patient outcomes. This is a highly significant problem due to the vast majority of patients who are taking these medications, and our general lack of knowledge regarding their impact on the cancer phenotype (11).

In this study, we report the novel discovery that lorazepam (LOR, Ativan®) and alprazolam (ALP, Xanax®), BZDs frequently prescribed to cancer patients to treat anxiety, impact patient survival outcomes across the cancer spectrum. We employ a combination of *in vivo* and *in vitro* models to mechanistically determine the effects of LOR and ALP on the PDAC TME. Specifically, we find that LOR promotes IL-6 secretion from CAFs, and drives ischemic necrosis and desmoplasia in mouse models of PDAC. To our knowledge, this is the first study to demonstrate that the commonly prescribed BZD lorazepam modifies the TME and has potential clinical implications when prescribing BZDs to cancer patients.

## Results

### Lorazepam is associated with poor survival outcomes in pancreatic cancer patients

To determine how frequently benzodiazepines (BZDs) are prescribed to cancer patients, we broadly examined BZD use in Roswell Park Comprehensive Cancer Center patients. We specifically assessed patients with primary cancers of the prostate, pancreas, ovary, kidney, head and neck, corpus uteri, colon, breast, brain, and those with invasive nevi/melanoma. Across all cancer types, 30.9% of patients had a record of BZD usage (Fig. 1A). Female patients had an equal or higher record of BZD prescriptions relative to males (34.2% vs. 27.4%) across all cancer types (Supplemental Fig. S1A). Pancreatic cancer patients had the highest record of BZD usage, with 40.6% of patients prescribed at least one BZD (Fig. 1A). Due to the high frequency of BZD use, we assessed the impact of BZDs on pancreatic cancer patient survival outcomes. We first evaluated how BZD prescription records correlated with survival outcomes in Roswell Park pancreatic cancer patients treated with chemotherapy from 2004-2020. Pancreatic cancer patients with a BZD prescription record had no significant difference in progression-free survival (PFS) (Supplemental Fig. S1B) but were associated with significantly improved disease-specific survival (DSS) relative to those without prescription records of BZDs (Supplemental Fig. S1C). Improved DSS can be partially attributed to imbalances in patient demographic and clinical characteristics; patients prescribed BZDs were significantly more likely to be white, younger, and were less likely to receive radiation therapy or surgery compared to non-BZD users (Table 1). Therefore, we performed covariate adjusted analyses to account for age, sex, race, clinical stage, additional treatments, and progressive disease. With these factors considered, DSS was significantly improved in patients prescribed BZDs ([HR: 0.70 (0.60, 0.82)]) (Supplemental Fig. S1D).

**Figure 1.**
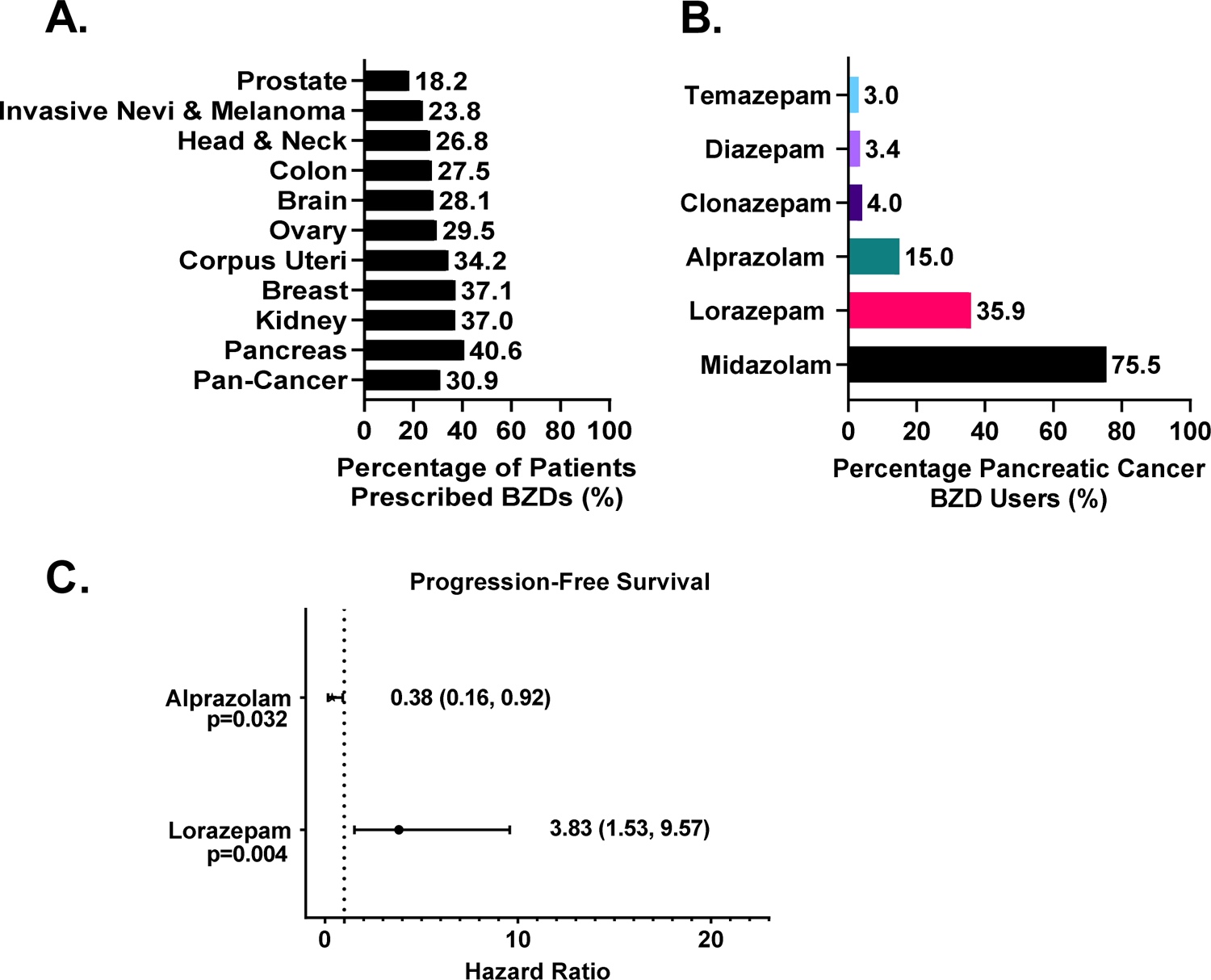
Lorazepam is associated with poor survival outcomes in pancreatic cancer patients. **(A)** Percentage of Roswell Park patients with a prescription record of benzodiazepines (BZDs) by cancer type. **(B)** Percentage of pancreatic cancer patients prescribed BZDs that are receiving the top six most commonly prescribed BZDs. **(C)** Covariate adjusted analysis evaluating the impact of lorazepam (n=40) or alprazolam (n=27) prescription records on pancreatic cancer patient progression-free survival accounting for age, sex, race, clinical stage, additional treatments, and progressive disease relative to no record of BZDs (n=69). Pan-cancer analyses refers to the combined average of all cancer types in the nSight database. <u>Statistics</u>: To account for potential imbalances in patient demographic and clinical characteristics, multivariable Cox regression models were used to evaluate the association between group (i.e. BZD usage) and the survival outcomes while adjusting for: age, sex, race, clinical stage, and additional treatments. Hazard ratios for BZD, with 95% confidence intervals, were obtained from model estimates. All analyses were conducted in SAS v9.4 (Cary, NC) at a significance level of 0.05.

We then sought to investigate if any specific commonly prescribed BZDs were associated with significant differences in survival. The most commonly prescribed BZD in pancreatic cancer, and all other cancer types with the exception of brain cancer, was midazolam, a short-acting (half-life 2-5 hr) agent often used as a sedative prior to surgery or medical procedures (Supplemental Fig. S1E) (12). The intermediate-acting (half-life 6-24 hr) BZDs lorazepam (LOR) and alprazolam (ALP) were the second and third most commonly prescribed BZDs to pancreatic cancer patients, respectively (Fig. 1B). LOR and ALP are frequently prescribed to pancreatic cancer patients to treat anxiety and anticipatory nausea prior to chemotherapy (10, 13). Due to the frequency of use and the longer-acting effect of LOR and ALP relative to midazolam, we assessed the impact of LOR and ALP on pancreatic cancer patient survival outcomes (Table 2-3). We performed covariate adjusted analyses to account for age, sex, race, clinical stage, and additional treatments (Table 4). Strikingly, LOR was associated with significantly worse PFS ([HR: 3.83 (1.53, 9.57)]) relative to patients not prescribed BZDs (Fig. 1C). In contrast, ALP was associated with significantly improved PFS ([HR: 0.38 (0.16, 0.92)]) relative to patients not prescribed BZDs (Fig. 1C). Collectively, we find that BZDs are commonly prescribed to pancreatic cancer patients. Importantly, specific BZD choice is associated with positive (ALP) or negative (LOR) survival outcomes.

### Lorazepam promotes ischemic necrosis and desmoplasia in murine PDAC tumors

Due to the differential effect of LOR and ALP on pancreatic cancer patient survival, we sought to characterize how these BZDs impact the growth and histology of murine pancreatic ductal adenocarcinoma (PDAC), the most common and deadly form of pancreatic cancer. We subcutaneously implanted LSL-KrasG12D/+; LSL-Trp53R172H/+; Pdx-1-Cre (KPC) tumor pieces into strain-matched, immunocompetent C57BL/6 mice (Fig. 2A). Our model accurately recapitulated the histology of the KPC spontaneous tumor as demonstrated by H&E staining (Fig 2B). The stromal compartment was maintained as indicated by α-SMA and vimentin staining, and the epithelial compartment was well-differentiated as evidenced by CK19 staining (Fig. 2B). To elucidate the effect of LOR and ALP on tumor growth, we treated C57BL/6 mice bearing KPC subcutaneous syngeneic allograft tumors with 0.5 mg/kg LOR or ALP daily until the tumors reached 2,000 mm^3^ or the mice reached endpoint criteria (Supplemental Fig. S2A). All the mice used in this study were female to match the sex of the syngeneic allograft tumor and there were no significant differences in the age, weight, and enrollment tumor size of the mice (Supplementary Fig. S2B-D). We did not observe significant differences in tumor growth or survival of the mice (Supplemental Fig. S2E-G). However, upon histological examination, we observed the presence of ischemic necrosis in tumors from LOR-treated mice (Supplemental Figure S2H, I). Next, we examined collagen deposition and found a significant increase upon BZD treatment, which was again most striking in the LOR-treated mice (Supplementary Fig. S2J, K). This experiment suggested that LOR may remodel the PDAC TME.

**Figure 2.**
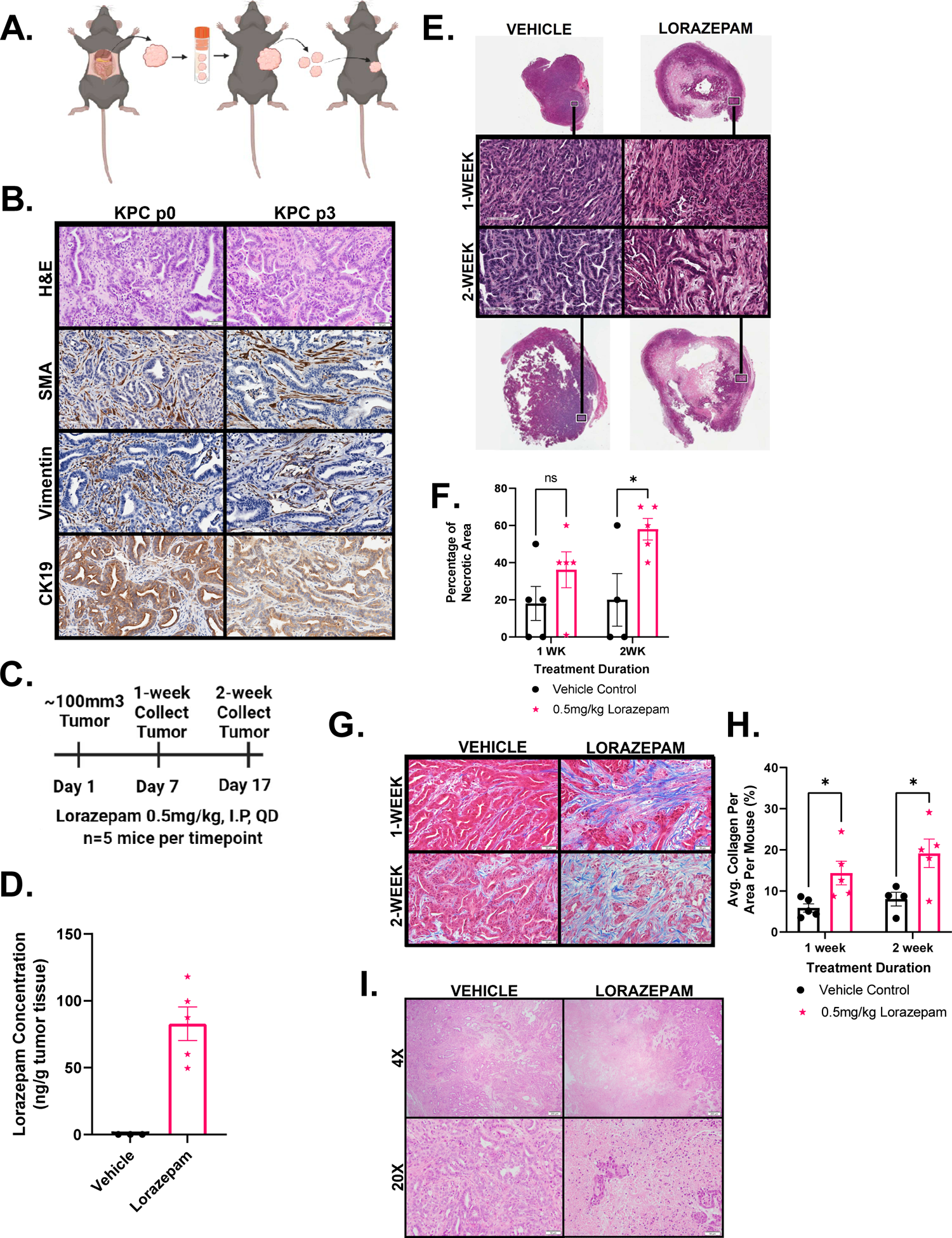
Lorazepam promotes ischemic necrosis and desmoplasia in murine PDAC tumors. **(A)** Schematic of subcutaneous LSL-KrasG12D/+; LSL-Trp53R172H/+; Pdx-1-Cre (KPC) syngeneic allograft model generation. **(B)** Comparison (top to bottom) of H&E, α-SMA IHC (20x), vimentin IHC (20x), and CK19 IHC (20x) in the KPC spontaneous tumor (left) and the p3 KPC syngeneic allograft derived from the KPC spontaneous tumor (right). **(C)** Experimental schematic of short-term LOR (n=5/arm) or vehicle treatment (n=4-5/arm). **(D)** Scatter plot with bar (mean with SEM) of LOR concentration per mouse quantified by liquid chromatography-mass spectroscopy (LC-MS) in the two-week LOR (n=5) or vehicle (n=3) treated subcutaneous KPC syngeneic allograft tumors collected two hr post-dosing. **(E)** Representative Aperio scanned H&E section of 1-week (top) and two-week (bottom) vehicle (left) and LOR (right) treated mice, representative zoomed in 20x images (black and white box) of 1-week (second row) and 2-week (third row) vehicle (left) and LOR (right) treated mice. **(F)** Quantification of the percentage of necrotic area per slide. **(G)** Representative 20x Masson’s trichrome images of 1-week (top) and 2-week (bottom) treated mice. **(H)** Quantification of the percentage of collagen per area. Image J color deconvolution plugin was used to quantify collagen area/20x field of 5 randomly selected images per mouse in a blinded manner. **(I)** Representative 4x (top) and 20x (bottom) H&E image of KPC spontaneous tumors treated with 0.5 mg/kg vehicle (left) or LOR (right) for two weeks (n=2-3/arm). <u>Statistics</u>: Groups were compared by mixed effects analysis with Bonferroni’s multiple comparison test, Black=Vehicle, Pink=0.5 mg/kg LOR.

To more definitively assess the impact of LOR on the TME, we performed a short-term treatment study. When the syngeneic subcutaneous allograft tumors reached 100 mm^3^, we treated the mice daily for 1-week or 2-weeks with 0.5 mg/kg LOR or vehicle (Fig. 2C). As noted in the previous study, all of the mice were female and there were no significant differences in murine age, weight, and enrollment tumor size (Supplementary Fig. S2L-N). To ensure therapeutic relevance, our dosing scheme was based on previous murine studies assessing the anxiolytic impact of LOR (14). We performed pharmacokinetic studies on endpoint tumors and found LOR concentrations of 49.6-118 ng/g, 2 hrs post-dosing (Fig. 2D). These concentrations were comparable to those observed in the brains of male CD-1 mice 1 hr post-intraperitoneal injection with 0.1-0.3 mg/kg LOR, supporting that the drug deposited in the tumor tissue at therapeutically relevant quantities (15). We performed H&E staining to identify histologic changes resulting from LOR treatment. Control tumors were differentiated with a well-defined stromal compartment (Fig. 2E). In contrast, LOR-treated tumors were more poorly differentiated, had increased stromal area, and had a significant increase in ischemic necrosis in the center of the tumors (Fig. 2E, F). LOR treatment did not impact endpoint tumor weight or tumor volume, supporting that increasing levels of necrosis was independent of tumor size (Supplemental Fig. S2O, P). Tumor size was likely maintained by the presence of rapidly proliferating tumor cells on the leading edge of the LOR-treated tumors, as indicated by Ki67 staining (Supplementary Fig. S2Q). Strikingly, we observed significant increases in collagen deposition at the 1 and 2-week timepoints (Fig. 2G, H), indicating that LOR-treatment increases desmoplasia. We did not observe any significant changes in collagen fiber integrated density, length, width, or straightness by second harmonic generation imaging (Supplemental Fig. S2R-U). Therefore, LOR promotes collagen deposition but not collagen remodeling. Next, we sought to extend these findings to the spontaneous KPC model. We treated KPC mice bearing 100 mm^3^ tumors daily with LOR (0.5 mg/kg) or vehicle for two weeks. Consistent with the transplant model, LOR treatment resulted in ischemic necrosis in KPC mice (Fig. 2I). Aggregately, these results support that LOR promotes desmoplasia within the PDAC tumor microenvironment.

### Lorazepam promotes inflammatory response and extracellular matrix signature in PDAC tumors

To assess transcriptional changes associated with LOR treatment, we performed RNA sequencing on the 2-week vehicle and LOR-treated subcutaneous syngeneic allograft tumors (Fig. 3A). There were 370 significantly upregulated genes and 617 significantly downregulated genes associated with LOR treatment. Consistent with increased stromal area and desmoplasia, we found a significant upregulation of extracellular matrix (ECM)-related genes, including *Serpinb2, Il6, Fgf7, Lox, Col6a4, Iga11, Pdpn,* and *Fap* in the LOR-treated tumors (Fig. 3A, B). We also observed a significant downregulation of the epithelial-related genes *Muc5ac* and *Gata3* (Fig. 3A, B). We performed pathway analysis to assess the top signaling pathways enriched upon LOR treatment. Among the top ten upregulated Kyoto Encyclopedia of Genes and Genomes (KEGG) pathways were Interferon Gamma Response, Interferon Alpha Response, Epithelial Mesenchymal Transition, TNF-alpha Signaling via NF-κB, Hypoxia, Complement, and IL-6/JAK/STAT3 Signaling (Fig. 3C-3F). These pathways, and IL-6, are highly enriched in the pro-inflammatory iCAF subpopulation (7, 16). While IL-6 has been reported to be associated with iCAFs, recent work has highlighted the extreme heterogeneity of CAF subtypes in the PDAC TME, and IL-6 is broadly expressed across multiple CAF subpopulations in murine PDAC models. Therefore, we determined if the LOR-induced IL-6 was produced in CAFs. To determine if upregulated IL-6 mRNA expression was produced by CAFs, we used RNAscope to perform RNA *in situ* hybridization (ISH) using an *Il6* probe, coupled with IHC for SMA. Intriguingly, we found that LOR was associated with a significantly higher number of IL-6 positive CAFs in both the KPC syngeneic and KPC spontaneous models (Fig. 3G, H; Supplemental Fig. S3A, B). These results indicate that LOR increases inflammatory signaling by CAFs and ECM-related gene expression in murine models of PDAC.

**Figure 3.**
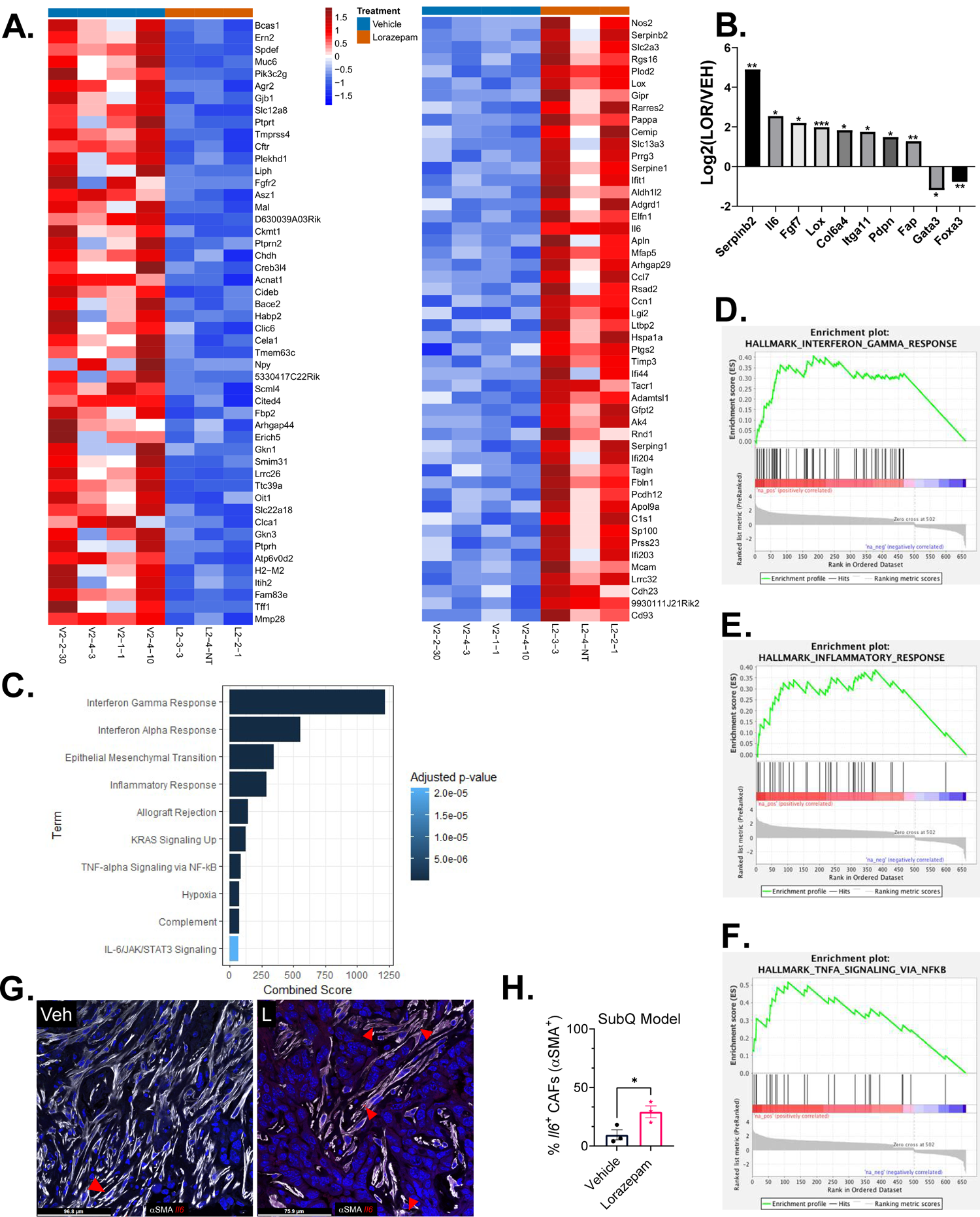
Lorazepam promotes inflammatory response and extracellular matrix signature in PDAC tumors. **(A)** Heat map of top 50 downregulated (left) and upregulated (right) genes in the 2-week LOR-treated (orange bar) subcutaneously implanted KPC tumors relative to the vehicle-treated (blue bar) tumors. **(B)** Differentially expressed extracellular matrix-related genes and epithelial genes in the 2-week LOR treated mice relative to the vehicle-treated mice. <u>Statistics</u>: adjusted p-value of log2 fold change of LOR/VEH. **(C)** Enrichr combined scores of the top 10 Enriched KEGG Terms in the two-week LOR-treated tumors relative to vehicle. **(D-F)** Enrichment plots of (D) Hallmark_Interferon_Gamma_Response (adjusted p-value 2.23E-36), (E) Hallmark_Inflammatory Response (adjusted p-value 1.98E-16), and (F) Hallmark_TNFA_Signaling_via_NFKB (adjusted p-value 5.57E-08). **(G)** Representative 40x RNAscope images of IL6+/SMA+ cells in the two-week treated vehicle (left) and LOR-treated subcutaneously implanted KPC tumors (n=3/arm). **(H)** Quantification of (G).

### GPR68 is preferentially expressed on human PDAC CAFs

We next sought to determine the molecular mechanism by which LOR regulates IL-6 production. First, we assessed the expression of common BZD targets in PDAC tumors, including the pentameric GABA-A receptors, the proton-sensing G-protein coupled receptor (GPCR) GPR68, and the translocator protein (TSPO, also known as the peripheral benzodiazepine receptor). We queried human PDAC single cell sequencing data from Peng *et al.* (17) and found that PDAC CAFs preferentially express *Gpr68* and the GABA-A receptor subunits *Gabra1*, *Gabrb2*, *Gabrg2*, and *Gabrr1* (Fig. 4A, Supplemental Fig. 4A).

**Figure 4.**
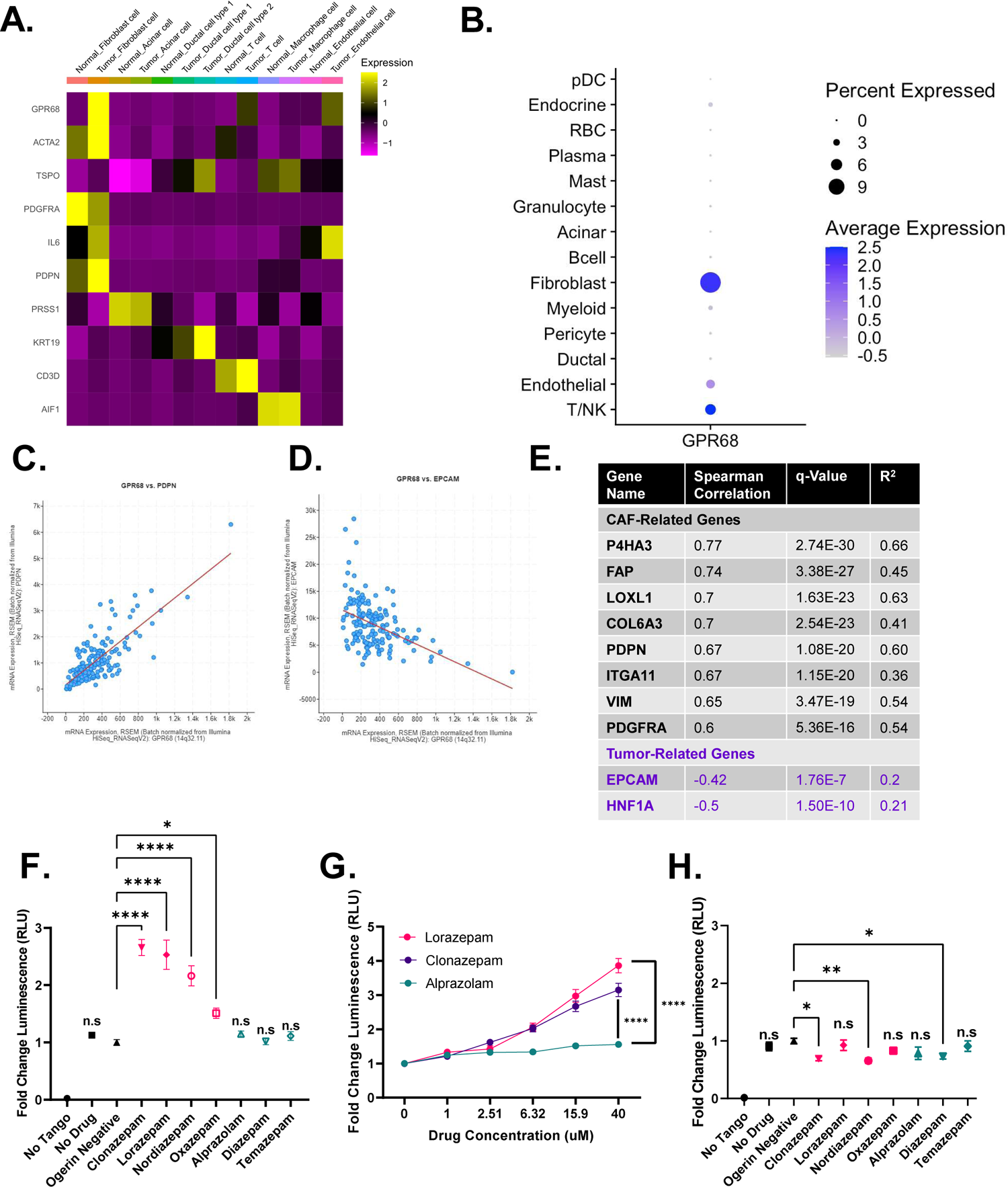
N-unsubstituted benzodiazepines potentiate activation of GPR68, a receptor preferentially expressed on human PDAC CAFs. **(A)** Heat map of *GPR68* and *TSPO* expression by cell type from the Peng *et al.* (17) human pancreatic ductal adenocarcinoma tumor single cell sequencing dataset. Yellow represents upregulated gene expression relative to other cell types within a row. **(B)** Dot plot visualization of *GPR68* gene expression level (color intensity) and frequency (size of dot) in different cell populations of human PDAC samples from Steele *et al.* (20). **(C,D)** Correlation plot of (C) *GPR68* and *PDPN*, and (D) *GPR68* and *EPCAM* in the human PDAC Pan-Cancer Atlas (TCGA dataset). **(E)** Summary table of the Spearman correlation of CAF-related genes with *GPR68* in the human PDAC Pan-Cancer Atlas (TCGA dataset). **(F-H)** PRESTO-Tango Assay for GPR68 activation (F) pH 6.8 BZD screen, (G) pH 6.8 dose-response curve for LOR, CLZ, and ALP, and (H) pH 7.4 BZD screen. Each plot represents the normalized average of 2-3 biological replicates. <u>Statistics</u>: BZD screens were analyzed by ordinary one-way ANOVA with Dunnett’s multiple comparison test, dose response curves were analyzed by two-way ANOVA with Holm-Šídák’s multiple comparisons test.

We chose to focus on GPR68, an acid-sensing receptor, for two reasons. First, activation of GPR68 in pancreatic CAFs is known to upregulate IL-6 secretion under acidic conditions (18). Second, n-unsubstituted BZDs (Supplemental Fig. S4B), such as LOR and clonazepam (CLZ), are strong positive allosteric modulators of GPR68, meaning they potentiate GPR68 activation only under acidic conditions. Conversely, n-substituted BZDs, including ALP, do not activate GPR68 (Supplemental Fig. S4B) (19). Therefore, we hypothesized that LOR increases inflammatory signaling by promoting GPR68 activation in CAFs. To further support that *GPR68* is preferentially expressed in CAFs, we analyzed human PDAC single cell sequencing data from Steele *et al.* (20). As observed in the Peng *et al.* (2019) dataset, *GPR68* was most highly enriched in human PDAC CAFs (Fig. 4B). Furthermore, there is a strong, significant positive correlation between *GPR68* and CAF-related genes, such as podoplanin (*PDPN*), and a strong, significant negative correlation between *GPR68* and epithelial-related genes, such as epithelial cellular adhesion molecule (*EPCAM*) in the human PDAC Pan-Cancer Atlas TCGA dataset (Fig. 4C-4E). To ensure that murine PDAC CAFs also express *Gpr68*, we reanalyzed single cell sequencing data from Kemp *et al.* (21). Similar to the human PDAC dataset, *Gpr68* was preferentially expressed on KPC tumor fibroblasts, T-cells, and endothelial cells (Supplemental Fig. S4C). We confirmed that murine CAFs express *Gpr68* by performing RNA ISH on KPC tumors and our syngeneic allograft tumors (Supplemental Fig. S4D). In addition to being expressed on CAFs, reanalysis of the CAF cluster in the human PDAC single cell sequencing by Steele *et al.* (20) indicated that *Gpr68* is not highly expressed on pericytes (RGS5 marker), supporting that it is a fibroblast-specific marker (Supplemental Fig. S4E-S4G). To determine the relationship between *GPR68* expression and PDAC progression, we reanalyzed *GPR68* expression by disease stage in the human PDAC single cell sequencing by Steele *et al.* (20). *GPR68* was not expressed strongly in the normal human pancreas but was expressed on PDAC primary tumors and PDAC metastases, supporting its likely role in disease pathogenesis (Supplemental Fig. S4H-J).

### N-unsubstituted benzodiazepines potentiate activation of GPR68

To identify which commonly prescribed BZDs were the strongest GPR68 activators, we performed PRESTO-Tango assays at pH 6.8, the optimal pH for GPR68 activation. This luciferase-based assay measures GPCR activity in a G-protein-independent manner. We found that at pH 6.8, the n-unsubstituted BZDs (LOR, CLZ, nordiazepam, and oxazepam) promoted GPR68 activation. In contrast, the n-substituted BZDs (ALP, diazepam, and temazepam) did not promote GPR68 activation (Fig. 4F). GPR68 activation by the n-unsubstituted BZDs LOR and CLZ was dose-dependent at pH 6.8, while the n-substituted BZD ALP did not activate GPR68 at any dose (Fig. 4G). When we re-screened the BZDs at pH 7.4 (a pH where GPR68 is not active), there was no significant increase in GPR68 activation by any BZD, supporting that n-unsubstituted BZDs are positive allosteric modulators of GPR68 (Fig. 4H).

Next, we sought to determine if murine PDAC tumors had a pH in the relevant range to support GPR68 activation. We assessed the pH of orthotopically implanted syngeneic KPC tumors (n=2), adjacent normal pancreas from the orthotopic model (n=1), bilaterally implanted subcutaneous KPC tumors (n=4), and the corresponding pancreata of the subcutaneously implanted tumors (n=2) using an H^+^ sensitive microelectrode. In the subcutaneous model, the normal pancreata had an average pH of 6.9568 +/- 0.1559. The tumors (weighing 0.985 g, 0.331 g, 0.214 g, and 0.078 g) were significantly more acidic, with an average pH of 6.7270 +/- 0.2292 (Supplemental Fig. S4K-S4M). Additionally, the subcutaneous tumors were well-differentiated with a clearly defined stromal compartment (Supplemental Fig. S4N). For the orthotopic model, the adjacent normal pancreas had a pH of 6.8833 (Supplemental Fig. S4O). Similar to the subcutaneous tumors, the orthotopic tumors (weighing 1.448 g and 1.713 g) were significantly more acidic than the normal pancreas with a pH of 6.6056 +/- 0.2313 and were well-differentiated with a well-defined stromal compartment (Supplemental Fig. S4P-S4R). Taken together, these results support that GPR68, a receptor preferentially expressed on PDAC CAFs, is activated by n-unsubstituted BZDs under acidic conditions present in the PDAC TME.

### Lorazepam promotes IL-6 secretion by human PDAC CAFs in a GPR68-dependent manner

Insel *et al.* (22) previously established that GPR68 activation in human CAFs increases IL-6 secretion in a cAMP-PKA-pCREB-dependent manner. We hypothesized that n-unsubstituted BZDs, including LOR, would increase IL-6 expression in CAFs in a GPR68-dependent and pH-dependent manner. First, we treated immortalized human CAFs with LOR for 3 hr at pH 6.8, and observed a significant increase in phospho-CREB (p-CREB) protein levels by western blot (Fig. 5A). Next, we assessed the role of LOR in regulating IL-6 expression. To determine if LOR modulated *Il6* mRNA expression, we treated immortalized human CAFs with LOR at pH 6.8 and performed qPCR. LOR significantly increased *Il6* expression at 24 hr (Fig. 5B). Similarly, *Il6* mRNA expression was significantly increased upon LOR treatment in human primary pancreatic CAFs (Fig. 5C). *Il6* mRNA expression was also significantly upregulated in the LOR-treated KPC syngeneic allograft tumors at the 2-week timepoint (Fig. 3B). Next, we performed an IL-6 ELISA which revealed that 24 hr LOR treatment significantly increased IL-6 protein secretion in immortalized human CAFs at pH 6.8 (Fig. 5D). Then, we evaluated whether GPR68 overexpression would promote even higher levels of IL-6 secretion. GPR68 overexpression in human immortalized CAFs significantly increased IL-6 secretion by LOR (Fig. 5E). In fact, 24 hr LOR treatment of human immortalized CAFs with GPR68 overexpression produced such high levels of IL-6 that the readings were too high to register (data not shown). To determine if LOR-mediated IL-6 secretion by CAFs was GPR68-dependent, we knocked down GPR68 in human immortalized CAFs using CRISPRi (Supplemental Fig. S5A). As expected, GPR68 knockdown potently decreased IL-6 levels (Supplemental Fig S5B). We then treated the control and GPR68 knockdown CAFs with LOR, CLZ, ALP, or DMSO at pH 6.8. GPR68 knockdown prevented LOR and CLZ from increasing IL-6 secretion at pH 6.8 (Fig. 5F). To determine if all GPR68 activator BZDs increase IL-6 secretion, we treated immortalized human CAFs with a panel of the most commonly prescribed BZDs at pH 6.8 and pH 8.0 for 24 hrs, collected the conditioned media, and performed an IL-6 ELISA. At pH 6.8, n-unsubstituted BZDs (GPR68 activators) significantly increased IL-6 secretion (Fig. 5G). Unexpectedly, n-substituted BZDs (non-activators) significantly decreased IL-6 secretion (Fig. 5G). When we performed the ELISA at pH 8.0, there was no significant increase in IL-6 secretion by the n-unsubstituted BZDs. This supports the contention that n-unsubstituted BZDs promote IL-6 secretion through GPR68 in CAFs (Fig. 5H). In contrast, at pH 8.0, n-substituted BZDs continued to significantly decrease IL-6 secretion, suggesting that this is occurring in a GPR68-independent manner (Fig. 5H). In fact, ALP still potently decreased IL-6 in the presence of GPR68 knockdown (Fig. 5F). We compared GPR68 activation by PRESTO-Tango with the ability of each BZD to increase IL-6 levels to further establish GPR68 dependence. We found that there was a direct correlation between the degree of GPR68 activation and the increase in IL-6 secretion by n-unsubstituted BZDs (Fig. 5I). There was no correlation between decreased IL-6 secretion and GPR68 activation by the n-substituted BZDs (Fig. 5J). To determine the relationship between GPR68 and IL-6 *in vivo*, we performed RNA ISH using *Il6* and *Gpr68* probes, and SMA IHC. In KPC tumors, LOR treatment significantly increased the number of triple positive (*Il6+/Gpr68+/*SMA*+*) cells, supporting that GPR68 increases IL-6 secretion by CAFs *in vivo* (Fig. 5K, L). In summary, these results indicate that BZDs differentially affect IL-6 secretion based on the structure of the BZD. N-unsubstituted BZDs promote IL-6 secretion under acidic conditions in a GPR68-dependent manner while n-substituted BZDs decrease IL-6 secretion in a pH and GPR68-independent manner.

**Figure 5.**
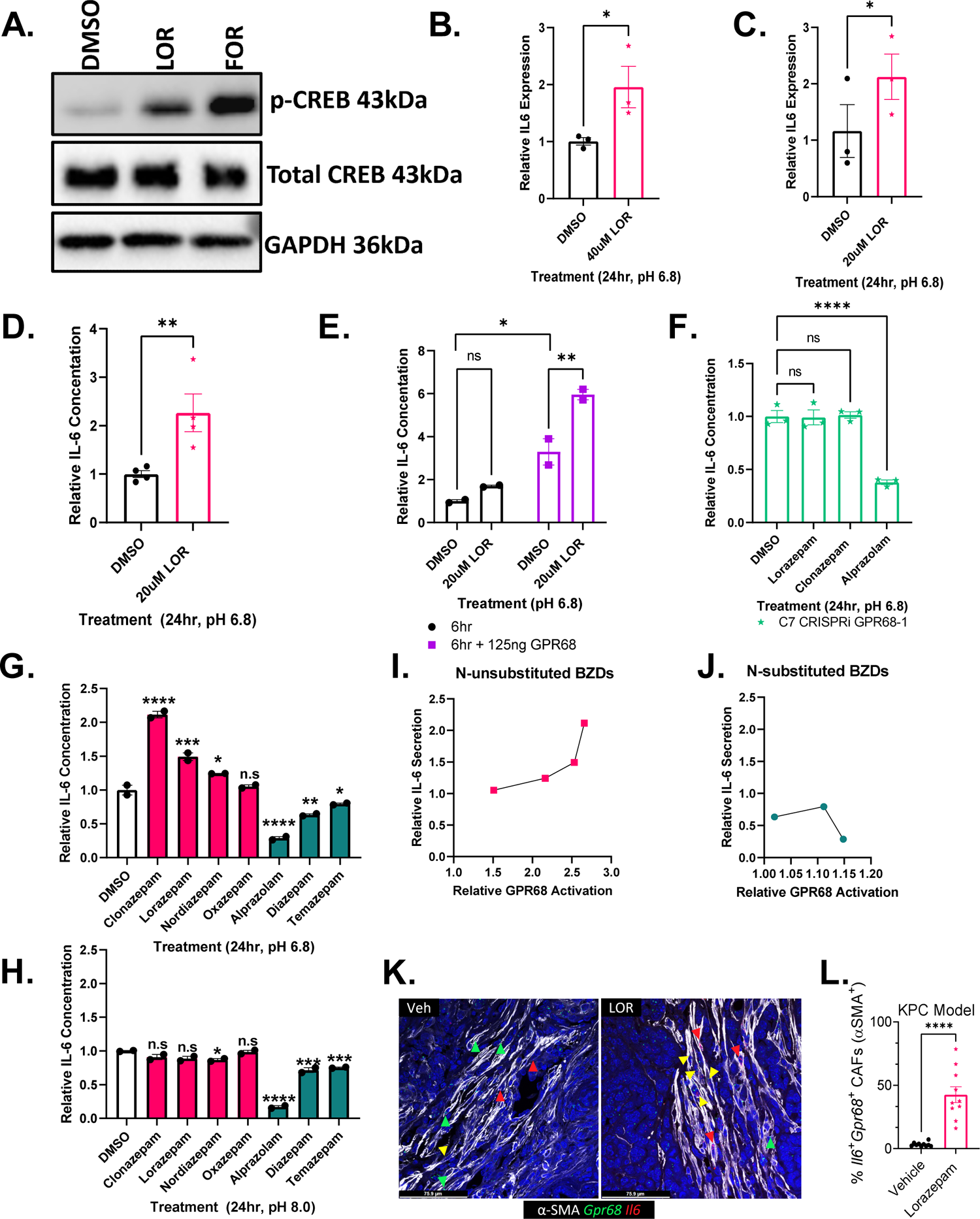
Lorazepam increases IL-6 secretion by human PDAC CAFs in a GPR68-dependent manner. **(A)** Western blot of immortalized human PDAC CAFs treated with LOR or forskolin (positive control) at pH 6.8 for 3 hr. **(B)** *Il6* qPCR of immortalized human PDAC CAFs treated with 40 μM LOR at pH 6.8 for 24 hr. **(C)** *Il6* qPCR of primary human PDAC CAFs treated with 20 μM LOR at pH 6.8 for 24 hr. **(D)** IL-6 ELISA of conditioned media from immortalized human PDAC CAFs treated with BZDs (20 μM) or DMSO control for 24 hr at pH 6.8. **(E)** IL-6 ELISA of conditioned media from immortalized human PDAC CAFs treated with 20 μM LOR or DMSO control for 6 hr in the presence or absence of GPR68 overexpression. **(F)** IL-6 ELISA of GPR68 knockdown immortalized human PDAC CAFs treated with LOR, CLZ, ALP, or DMSO control for 24 hr at pH 6.8. **(G-H)** IL-6 ELISA of conditioned media from immortalized human PDAC CAFs treated with BZDs (20 uM) or DMSO control for 24 hr at (G) pH 6.8 or (H) pH 8.0. Pink represents n-unsubstituted BZDs, teal represents n-substituted BZDs. **(I, J)** Correlation plot of relative GPR68 activation of each BZD by PRESTO-Tango relative to IL-6 secretion by IL-6 ELISA for (I) n-unsubstituted BZDs and (J) n-substituted BZDs at pH 6.8. **(K)** Representative 40x RNAscope images of IL6+/GPR68+/SMA+ cells in the two-week treated vehicle (left) and LOR-treated KPC tumors. **(L)** Quantification of (K). All experiments are representative of 2-4 biological replicates. <u>Statistics</u>: To analyze two groups, paired/unpaired one-tailed t-tests were performed. For analysis of multiple groups, we performed ordinary one-way ANOVA with Bonferroni’s multiple comparison test. In the case of multiple groups with two independent variables, groups were compared by two-way ANOVA with with Holm-Šídák’s multiple comparisons test.

### Lorazepam is associated with worse patient survival across multiple cancer types

Based on the differential effect of BZDs on IL-6 secretion by CAFs (Fig. 5), and the established role of IL-6 in promoting worse clinical outcomes (23–25), we compared overall survival (OS) differences in Roswell Park patients (2000–2022) prescribed LOR or ALP relative to patients with no record of BZDs treated for primary cancers of the brain (Table 5), breast (Table 6), corpus uteri (Table 7), head and neck (Table 8), skin (Table 9), kidney (Table 10), ovary (Table 11), colon (Table 12), and prostate (Table 13). LOR and ALP are commonly prescribed to patients with these cancer types (Supplemental Fig. S6A, S6B). We calculated hazard ratios accounting for sex (where applicable), clinical grade, and clinical stage. LOR was associated with significantly worse OS and progression-free survival (PFS) in prostate cancer [HR OS: 2.160 (1.589, 2.936), HR PFS: 1.899 (1.433, 2.517)], ovarian cancer [HR OS: 1.521 (1.212, 1.907), HR PFS: 1.464 (1.174, 1.826)], invasive nevi/melanoma [HR OS: 1.978 (1.519, 2.576), HR PFS: 2.195 (1.699, 2.835)], head and neck cancer [HR OS: 1.629 (1.304, 2.035), HR PFS: 1.635 (1.313, 2.036)], colon cancer [HR OS: 1.620 (1.317, 1.993, HR PFS: 1.782 (1.457, 2.179)], and breast cancer [HR OS: 1.248 (1.050, 1.484), HR PFS: 1.345 (1.138, 1.591)] relative to patients not prescribed BZDs (Fig. 6A, Supplemental Fig S6C). In contrast, ALP was infrequently associated with significant differences in survival outcomes, with the exception of hormonal cancers where there was significantly worse OS and PFS in breast cancer [HR OS: 1.867 (1.528, 2.281), HR PFS: 1.850 (1.523, 2.248)], worse OS in prostate cancer [HR OS: 1.464 (1.038, 2.064)], and worse PFS in uterine cancer patients [HR PFS: 1.668 (1.051, 2.646)] (Fig. 6B, Supplemental Fig. S6D). Intriguingly, LOR was associated with significantly improved OS in patients with brain cancer (Fig. 6A). LOR and ALP did not correlate with altered survival outcomes in kidney cancer (Fig 6A, B, Table 10). The Kaplan-Meier curves for OS and PFS for melanoma (Fig. 6C, Supplemental Fig. S6E), prostate cancer (Fig. 6D, Supplemental Fig. S6F), and ovarian cancer (Fig. 6E, Supplemental Fig. S6G) clearly demonstrate that LOR correlates with worse survival outcomes relative to patients prescribed ALP or those with no record of BZD use. Overall, we find that LOR is associated with poor survival outcomes across multiple cancer types.

**Figure 6.**
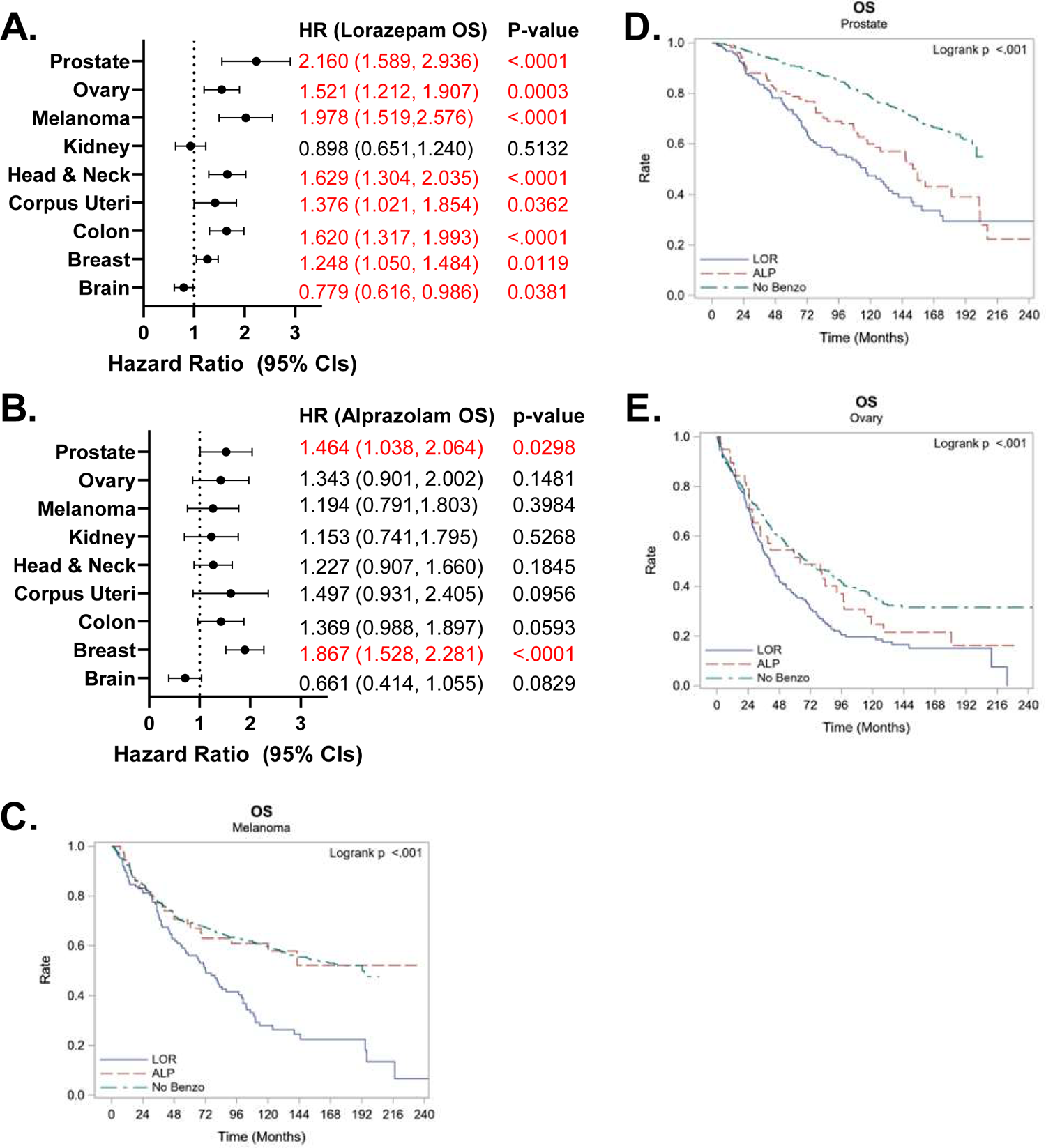
Lorazepam is associated with worse patient survival across multiple cancer types. **(A,B)** Association between prescription or infusion records of (A) LOR or (B) ALP and OS by cancer type in Roswell Park patients with a diagnostic date from 2000-2022, significant values are highlighted in red. **(C-E)** Kaplan Meier curve comparing OS in Roswell Park patients with prescription or infusion records of LOR or ALP, or those with no history of BZD use treated for primary (C) invasive nevi or melanoma, (D) prostate cancer, or (E) ovarian cancer. <u>Statistics</u>: Multivariate Cox regression modeling was performed to measure associations between survival outcomes and cohort. Models were adjusted for sex (where applicable), clinical grade, and clinical stage. HR and corresponding 95% CIs were provided for individual LOR and ALP groups, with ‘No Benzo’ as the referent group. Type 3 Test was used and an overall p-value measuring the association between survival and cohort was provided. CI: confidence interval, HR: Hazard ratio, OS: overall survival.

## Discussion

We provide evidence that the commonly prescribed anti-anxiety drug LOR promotes desmoplasia in the PDAC tumor microenvironment (Figs. 2, 3), IL-6 secretion from CAFs (Fig. 5), and is associated with poor cancer patient survival outcomes (Figs. 1, 6). Retrospective epidemiological studies found that LOR was associated with worse progression-free survival (PFS) while ALP was associated with improved PFS in pancreatic cancer patients (Fig. 1). LOR promotes desmoplasia (Fig. 2), inflammatory signaling (Fig. 3), IL-6 expression in CAFs (Fig. 3, 5) and ischemic necrosis in murine PDAC models (Fig. 2). LOR is likely promoting inflammatory signaling and IL-6 secretion by CAFs through activation of GPR68. GPR68 is preferentially expressed on human PDAC CAFs and n-unsubstituted BZDs significantly increase GPR68 activation under acidic conditions (Fig. 4). LOR increases IL-6 expression and secretion in human immortalized CAFs in a pH and GPR68-dependent manner (Fig. 5). Conversely, ALP, and other GPR68 non-activator BZDs decrease IL-6 in human immortalized CAFs in a pH and GPR68-independent manner (Fig. 5). We propose that LOR stimulates fibrosis and inflammatory signaling, promoting desmoplasia and ischemic necrosis, decreasing pancreatic cancer patient survival. Across many cancer types, LOR is associated with worse survival outcomes, supporting a pro-tumorigenic role (Fig. 6).

In the context of cancer, BZDs are commonly used in palliative care (26). Zabora *et al.* (27) assessed psychological distress in patients with cancer and found that pancreatic cancer produced the highest scores for anxiety and depression. Approximately 48% of pancreatic cancer patients had symptoms of an anxiety-related disorder (28). A larger, more recent, cross-sectional study by Clark *et al.* (29) found that approximately 30% of pancreatic cancer patients had distressing levels of anxiety. Wilson *et al.* (30) compiled Canadian survey data and found that two-thirds of cancer patients with depression or anxiety are prescribed BZDs. Our epidemiology studies further corroborated these findings, indicating that across multiple cancer types, 30.9% of Roswell Park patients received at least one BZD, with pancreatic cancer patients having the highest rate of BZD prescriptions relative to the cancer types evaluated (40.6%). High usage of BZDs is concerning because many epidemiological studies have found that BZDs increase the risk of cancer (31–36). However, few experimental studies have been performed to mechanistically link BZDs to increased cancer risk. Studies in mice and rats have shown that diazepam and oxazepam can spontaneously induce liver cancer and clobazam can induce thyroid cancer (37–39). These studies support that BZD use may promote cancer development but no study has definitively addressed the association between BZDs and human cancer progression.

To our knowledge, our research is the first retrospective cohort study to assess the association between BZDs and cancer patient survival, accounting for potential confounding variables, including disease stage (Fig. 6). We are also the first to perform a comprehensive analysis regarding the association between BZDs and pancreatic cancer survival outcomes (Fig. 1). Previously, O’Donnell et al. (40) performed a systematic review to determine the relationship between BZDs and cancer patient survival. Their cohort was primarily late stage cancer patients receiving the short-acting sedative BZD, midazolam. Unsurprisingly, they did not observe significant survival differences.

Experimentally, few studies have quantified the effect of commonly prescribed BZDs on cancer progression and the TME. Oshima *et al.* studied the impact of the short-acting BZD midazolam on LSL-Kras^G12D/+^; Trp53*^flox/flox^*; Pdx-1^cre/+^ (KPPC) mice (41). They found that midazolam slowed tumor growth/proliferation, decreased inflammatory cytokine production (including IL-6), and reduced the number of α-SMA+ cells. The phenotype was reversed by PK11195, a TSPO antagonist, suggesting that inhibition of inflammatory cytokine production is a TSPO-dependent process. Our studies are the first to test physiologically relevant doses of LOR in immunocompetent cancer models with intact stroma (Fig. 2). Fafalios *et al.* (42) found that LOR decreased prostate cancer cell growth *in vivo*. Their study used very high LOR concentrations (40 mg/kg) and differences in tumor growth were only observed at very large tumor volumes in immunocompromised mice. Additionally, they suggested that LOR was exerting its biological effects via TSPO; however, a more recent study by Huang *et al.* (19) assessed the off-target activities of LOR using radioligand binding assays and found that LOR did not bind TSPO. Therefore, it is necessary to determine the off-target activities of a panel of BZDs to be certain which ones bind TSPO and whether they function as agonists or antagonists in the context of cancer. Previous studies in rats injected intravenously with W-256 carcinosarcoma cells indicate that ALP inhibits lung metastases in a central BZD receptor-dependent manner (43). Additionally, ALP, LOR, and CLZ enhance or suppress immune function in cancer and non-cancer settings (44–48). We are the first to comprehensively assess the impact of commonly prescribed BZDs on interleukin-6 (IL-6) signaling by CAFs (Fig. 5).

IL-6 plays important roles in pancreatic cancer development and progression (49). Inhibition of IL-6 improves the efficacy of PD-L1 immunotherapy in mouse models (50). Conversely, high IL-6 levels are associated with lower survival and decreased gemcitabine efficacy in PDAC patients (8). We show that there is a strong association between BZDs and survival outcomes in PDAC patients receiving chemotherapy. Additional epidemiology studies should be performed to determine if BZDs are associated with altered survival outcome in cancer patients receiving immunotherapy drugs.

IL-6 is also associated with a specific subset of pancreatic CAFs, known as inflammatory CAFs or iCAFs, characterized by high expression of inflammatory cytokines (51). Due to the pro-tumorigenic nature of IL-6, this subtype is presumed to be associated with poor survival outcomes relative to myofibroblastic CAFs (myCAFs), which are characterized by high levels of alpha-smooth muscle actin (α-SMA) (7). Interestingly, pathway analysis of our LOR-treated tumors overlapped significantly with iCAF-related signaling pathways (Figure 3C-3F), supporting that LOR may increase the level of iCAFs (7, 16). It is well established that CAF subtypes are plastic (51). We identify a significant increase in IL6+/SMA+ cell populations in murine PDAC tumors, suggesting that LOR may promote CAF subtype plasticity (Figure 3G, 3H, Supplemental Fig. S3A, S3B).

Although BZDs have previously been shown to alter IL-6, we are the first to show that BZDs alter IL-6 secretion in a pH and GPR68-dependent manner (Figure 5). BZDs produce therapeutic effects by binding GABA-A receptors, particularly *α1β2γ2* GABA-A receptors, which were GABA subunits we found to be preferentially expressed on human pancreatic CAFs (Supplemental Fig. S4A) (52). BZDs prescribed for anxiety have similar affinities for the different GABA-A receptor subtypes but differ in potency, half-life, and how the drugs are metabolized (13). As of 2016, there were 14 FDA-approved BZDs (53). New BZDs are being synthesized every year, for both licit and illicit purposes (54), highlighting the strong need to fully understand how these drugs impact human physiology and disease pathology. GABA-A (muscimol) and GABA-B (baclofen) receptor agonism is known to decrease stress-induced plasma IL-6 in murine models (55). In other contexts, BZDs have been shown to differentially affect IL-6. For example, similar to findings by Oshima *et al.* (41), midazolam decreased *Il6* expression in peripheral blood mononuclear cells. However, in that study, TSPO agonism and CLZ (which does not bind TSPO) did not downregulate *Il6* (56), suggesting that IL-6 modulation is through an alternative mechanism. Additional studies are required to determine the mechanism by which n-substituted BZDs, such as ALP, which do not activate GPR68, decrease IL-6 levels in PDAC CAFs.

An off-target effect of n-unsubstituted BZDs is positive allosteric modulation of GPR68 (Fig. 4). GPR68 activation is known to increase IL-6 and IL-8 in various cell types (57–60). GPR68 knockdown inhibits IL-6 secretion by CAFs in a G_s_-cAMP-PKA-CREB-dependent manner (18). Our studies are the first to determine how BZDs influence GPR68 signaling in pancreatic CAFs. To our knowledge, we are also the first to determine the pH of subcutaneous and orthotopically implanted KPC tumors using a microelectrode pH meter. High *et al.* (61) measured the pH of murine pancreatic tumors from cerulean-treated K-ras^LSL.G12D/+^; Pdx-1-Cre (KC) mice using acidoCEST magnetic resonance imaging (MRI), a non-invasive method of measuring extracellular pH. Similar to our findings (Supplemental Fig S4K-S4R), when pancreatitis was likely present, the pancreatic pH was 6.85-6.92, which is more acidic than mice without cerulean treatment (pH 6.92-7.05). Tumor-bearing mice had the most acidic pH of approximately 6.75-6.79 (5-8 weeks post-cerulean treatment). Acidic pH can alter the TME by modulating enzymatic function, as well as by promoting epithelial to mesenchymal transition, metastasis, and T cell anergy (62–65), common features of pancreatic cancer. To ensure that pancreatic cancer research is physiologically relevant, it is vital that *in vitro* models accurately mimic the acidic pH conditions observed *in vivo*.

In addition to impacting inflammation, GPR68 regulates fibrosis and mechanosensing, important factors in promoting pancreatic cancer development and progression. GPR68 promotes fibrosis and pro-fibrotic cytokine production in ileum grafts, airway smooth muscle cells, and human PDAC CAFs (18, 66, 67). An unbiased screen revealed GPR68 as a fibroblast-specific drug target in colon cancer (57). Knockdown of GPR68 in bone marrow-derived mesenchymal stem cells (BMSCs), which have previously been shown to convert to CAFs, slowed tumor growth when subcutaneously co-injected with tumor cells into nude mice, further supporting the CAF-specific importance of GPR68 in cancer (68). Additionally, mechanosensing and acid-sensing are vital to fibrosis and cancer cell survival. GPR68 senses and responds to membrane stretch and shear stress, regulating blood vessel dilation/remodeling (69–71). Wei *et al.* (71) proposed that GPR68 will likely be a potential drug target for solid cancers and fibrotic diseases, thus the role of GPR68 in pancreatic cancer, which is very fibrotic, is highly relevant. The role of GPR68 in sensing membrane stretch may even aid its roles in metastasis and may dictate the morphological alterations in pancreatic stellate cell (PSC) activation to CAFs. Future studies should be performed to assess the effect of BZDs on the tumor vasculature, PSC activation, and metastasis.

Future work should also assess the tumor intrinsic, GPR68-independent effect of BZDs. We found that TSPO and the GABA-A receptor subunits *GABRA2*, *GABRA3*, *GABRA4*, and *GABRQ* are preferentially expressed in tumor ductal cell type 2, the more aggressive subset of PDAC tumor cells, as determined by human PDAC single cell sequencing data (17) (Fig. 4A, Supplemental Fig. S4A). Our data strongly support that LOR is likely impacting ischemic necrosis and desmoplasia in a CAF-dependent manner (Fig. 2). However, GPR68-independent mechanisms can influence the TME, as evidenced by the presence of ischemic necrosis and increased collagen levels in ALP-treated mice (Supplemental Fig. S2H-2K). Additionally, in our pan-cancer analysis not all cancer types are as dependent on CAFs as PDAC, but dramatic differences in survival outcomes are still observed. By comparing the cell and tissue-specific roles of GPR68, TSPO, and GABA-A receptors and determining which receptor type is likely playing an important role in each cancer type we can begin to delineate the exact mechanism by which BZDs are impacting patient survival across different cancer types.

In summary, we have interrogated the role of BZDs on the PDAC TME and patient survival. We made the significant, novel discovery that certain types of BZDs may negatively impact cancer patient survival, while others may be beneficial. Due to the frequency that BZDs are prescribed, this is an issue that could impact a large percentage of cancer patients. Performing prospective clinical trials and additional experimental studies to determine whether BZDs impact therapeutic efficacy, is vital. Physicians could improve patient outcomes by optimizing BZD prescribing practice to maximize cancer patient survival while providing necessary palliative care. Additionally, this research provides a platform to guide others interested in determining how commonly prescribed drugs influence the tumor microenvironment via on-target or off-target mechanisms.

## Methods

### Benzodiazepine Prescription Frequency

We used Roswell Park Comprehensive Cancer Center’s web-based tool, nSight™, which allows users to explore and analyze clinical data. We compared BZD prescription records (alprazolam, lorazepam, chlordiazepoxide, clobazam, clonazepam, clorazepate, diazepam, estazolam, flurazepam, midazolam, oxazepam, temazepam, and triazolam) in Roswell Park patients with primary cancers of the prostate, pancreas, ovary, kidney, head and neck, corpus uteri, colon, breast, brain, and those with invasive nevi/melanomas. Pan-cancer analysis assessed all Roswell Park patients. Patients with multiple primary cancers were excluded. The data were acquired on February 3, 2023.

### Pancreatic Cancer Epidemiology Study

This study assesses the effect of BZD prescription on the survival outcomes of Roswell Park pancreatic cancer patients treated with chemotherapy from 2004-2020. Patients who did not receive chemotherapy (n=4) or had clinical stage 0 disease (n=2) were removed. Patient characteristics were summarized by BZD use (overall and by type, Tables 1-3) using the mean, median, and standard deviation for quantitative variables; and using frequencies and relative frequencies for categorical variables. Comparisons were made using the Mann-Whitney U or Kruskal-Wallis tests for quantitative variables, and Fisher’s exact or Chi-square tests for categorical variables. The time-to-event outcomes were summarized by group using standard Kaplan-Meier methods, where the 1/3-year rates and medians were estimated with 95% confidence intervals. Associations were evaluated using the log-rank test. Overall survival (OS) is defined as the time from first chemotherapy until death due to any cause or last follow-up. Disease-specific survival is defined as the time from chemotherapy until death due to cancer or last follow-up. Progression-free survival (PFS) is only defined in those who were disease-free (i.e. non-persistent disease), and is the time from chemotherapy until recurrence, death from disease, or last follow-up. Disease-free survival (DFS) is defined as the time from chemotherapy until persistent disease, recurrence, death from disease, or last follow-up. To account for potential imbalances in patient demographic and clinical characteristics, multivariable Cox regression models were used to evaluate the association between group (i.e. BZD usage) and the survival outcomes while adjusting for: age, sex, race, clinical stage, additional treatments, and progressive disease (for OS and DSS only). Hazard ratios for BZD, with 95% confidence intervals, were obtained from model estimates. All analyses were conducted in SAS v9.4 (Cary, NC) at a significance level of 0.05.

### LSL-Kras^G12D/+^; LSL-Trp53^R172H/+^; Pdx-1-Cre (KPC) Subcutaneous Syngeneic Allograft Long-Term Study

A subcutaneously passaged KPC002 allograft derived from a female KPC mouse was stored in freezing media (50% RPMI, 40% FBS, 10% DMSO) in liquid nitrogen. The p3 allograft tissue was passaged once in strain-matched C57BL/6 female mice by dipping the tumor tissue (2-3 mm in size) in Corning Matrigel (Cat. #356231) and implanting the tissue bilaterally into the flank of each mouse. The tumor tissue was harvested 2 weeks later. ∼0.55 mm^3^ tumor pieces were implanted into the left flank of 24 C57BL/6 female mice. When the tumors reached 100-200 mm^3^ the mice were enrolled into the study. Each mouse was treated with 0.5 mg/kg lorazepam or DMSO control (0.25% DMSO in a sodium chloride solution (0.9%), Sigma Cat. #S8776) daily by intraperitoneal (IP) injection. A 50 μg/mL lorazepam was prepared fresh by diluting a 20 mg/mL stock of lorazepam (Sigma-Aldrich, Cat. #L1764) or alprazolam (Sigma-Aldrich, Cat. #A8800) dissolved in DMSO in a sodium chloride solution (0.9%), Sigma Cat. #S8776) and each mouse received 0.01 mL/g. Mice were weighed daily, and tumor growth was measured biweekly using Fisherbrand Traceable Digital Calipers (0-150 mm). When the tumors measured 2,000 mm^3^ or other endpoint criteria were reached, the mice were sacrificed two hrs after drug administration.

### KPC Subcutaneous Syngeneic Allograft Short-Term Study

A subcutaneously passaged KPC002 allograft derived from a female KPC mouse was stored in freezing media (50% RPMI, 40% FBS, 10% DMSO) in liquid nitrogen. The p2 allograft tissue was passaged once in strain-matched C57BL/6 female mice by dipping the tumor tissue (2-3 mm in size) in Corning Matrigel (Cat. #356231) and implanting the tissue bilaterally into the flank of each mouse. The tumor tissue was harvested 2 weeks later. ∼0.55 mm^3^ tumor pieces were implanted into the left flank of 20 C57BL/6 female mice. When the tumors reached 100-200 mm^3^ the mice were enrolled into the study. Each mouse was treated daily with 0.5 mg/kg lorazepam or DMSO control (0.2% DMSO in a sodium chloride solution (0.9%), Sigma Cat. #S8776) by intraperitoneal (IP) injection. A 50 μg/mL lorazepam was prepared fresh by diluting a 25 mg/mL stock of lorazepam (Sigma-Aldrich, L1764, LOT#035F0115) dissolved in DMSO in a sodium chloride solution (0.9%, Sigma Cat. #S8776) and each mouse received 0.01 mL/g. Mice were weighed daily, and tumor growth was measured daily using Fisherbrand Traceable Digital Calipers (0-150 mm). After 1 or 2 weeks the mice were sacrificed two hrs after drug administration.

### LC-MS Analysis of Subcutaneous Syngeneic KPC Allograft Tumors

141.9-255.6 mg mouse tumor pieces (2-week timepoint, 2 hours post-dosing) were snap frozen in homogenizing tubes and stored at −80°C. Prior to analysis the tumors were homogenized in 25% methanol. Calibrators, quality controls, plasma blanks, and study samples were thawed and vortexed for 5-10 seconds. To separate 1.5 mL microcentrifuge tubes, 50 μL of spiking solution was added to 50 μL of blank plasma for calibrators A-I and quality controls. 50 μL of 50% methanol was added to 50 μL plasma blank with internal standard, plasma blank, reagent blank (water), and study samples. 200 μL of WIS was added to each sample (or 100% methanol to plasma blank and reagent blank) using a repeater pipet and vortexed for ∼10 seconds. Samples were allowed to digest for 10 min in the refrigerator or on wet ice. Samples were vortexed for ∼10 seconds and centrifuged at 13,500 rpm for 10 min at 4°C. 150 μL of each sample was transferred to the autosampler vial and 5.00 μL were injected into the LC-MS/MS (Sciex 5500 QTrap) system.

### H&E

Freshly isolated tumors were fixed in 10% neutral buffered formalin solution (Sigma-Aldrich, Cat. # HT501128) for 24 hr prior to processing. Tumor processing was performed in the Experimental Tumor Model (ETM) Shared Resource using a HistoCore Arcadia H (Leica) embedder and sliced in 5 µm sections using a RM2235 (Leica) microtome. FFPE unstained slides were rehydrated as follows: xylene: 5 min (repeat 3 times), 100% ethanol: 10 min, 95% ethanol: 10 min (repeat twice), 70% ethanol: 10 min, distilled water 5 min. The slides were then placed in hematoxylin for 2 min, rinsed with cold running tap water for ∼3min, dipped twice in 1% acid alcohol, rinsed with cold running tap water until tissue turned blue color. Next, the slides were placed in 95% ethanol for three min, eosin for 30 seconds, dipped in 95% ethanol 4-5 times, and dehydrated as follows: 95% ethanol: 3 min, 100% ethanol: 3 min, xylene: 3 min (repeat twice), xylene: 5 min. Slides were dried briefly and cover-slipped using Poly-Mount.

### Ischemic Necrosis Quantification

H&E slides were imaged using the ScanScope XT System and necrotic area relative to total area was determined in a blinded manner by a PDAC pathologist.

### Masson’s Trichrome

Freshly isolated tumors were fixed in 10% neutral buffered formalin solution (Sigma-Aldrich, Cat. #HT501128) for 24 hr prior to processing. Tumor processing was performed in the ETM Shared Resource using a HistoCore Arcadia H (Leica) embedder and sliced in 5 µm sections using a RM2235 (Leica) microtome. Tissue was rehydrated as follows, xylene: 3 min (repeat three times), 100% ethanol: 3 min (repeat three times), 95% ethanol: 3 min, 70% ethanol: 3 min, deionized water: 5 min. The Abcam trichrome stain kit (ab150686) was then used according to the manufacturer’s instructions. For step 5.9 the slides were rinsed in distilled water for 2 min and in step 5.12 the slides were rinsed in distilled water for 30 seconds. The slides were dehydrated as follows, 95% ethanol: 3 min (repeat twice), 100% ethanol: 3 min (repeat twice), and xylene: 5 min (repeat three times). The slides were dried briefly and cover-slipped using Poly-Mount.

### Immunohistochemistry

Freshly isolated tumors were fixed in 10% neutral buffered formalin solution (Sigma-Aldrich, Cat. #HT501128) for 24 hr prior to processing. All immunohistochemistry processing and staining was performed in the ETM Shared Resource using an AutoStainer Plus (Dako) using the antibodies alpha-smooth muscle actin (Sigma, Cat. #A5228), vimentin (Cell Signaling, Cat. #5741S), cytokeratin-19 (Abcam, Cat. #ab15463), and Ki67 (Abcam, Cat. #ab15580).

### Second Harmonic Generation (SHG) of Polarized Light Detection and Analysis

As previously reported (72), imaging of SHG signal from collagen bundles was performed with a Leica SP8 DIVE confocal/multiphoton microscope system (Leica Microsystems, Inc., Mannheim, Germany), using a 25X HC FLUOTAR L 25x/0.95NA W VISIR water-immersion objective. H&E stained specimens were excited at 850 nm employing an IR laser Chameleon Vision II (Coherent Inc., Santa Clara, CA), and blackguard SHG emitted signal was collected using a non-descanned detector configured to record wavelengths between 410-440 nm. Under pathologist supervision, two different areas containing tumor and stromal tissue were selected from three different animals of each cohort. Using the automated Leica Application Suite X 3.5.5 software, 2-4 regions of interest (ROI) from each area, were set up for SHG signal collection using identical settings and recorded as monochromatic, 16-bit image stacks of 5 μm depth (Z total distance). Image processing and digital analyses were conducted via FIJI (ImageJ 1.52p; https://fiji.sc/) software (73). Raw image stack files were tri-dimensionally reconstituted as two-dimensional maximal projection 16-bit images. For all images, signal to noise identical thresholds were set. Resultant SHG positive-signal pixels were used to calculate integrated densities (e.g., SHG signal/SHG area). SHG integrated density data were standardized to the mean value obtained from vehicle cohort. [FJ1] Results represent SHG arbitrary units compared to control tissues. Additionally, CT-FIRE (V2.0 Beta; https://eliceirilab.org/software/ctfire/) software (74) was used for individual collagen fiber (SHG-positive) architecture analyses. Following the pipeline described by the authors in the provided manual document, SHG images were loaded in batches organized by cohorts. Using similar settings for both groups, single collagen fibers were analyzed for length, width, and straightness. A threshold for fibers with a minimum of 10 μm length was set to reduce error from smaller objects detected. Readouts were plotted in graphs, expressed in micron units for length, width parameters, and arbitrary units for fiber straightness.

### KPC Short-Term Lorazepam Study

Male and female KPC mice (n= 2-3/arm) were enrolled when their tumors reached 100-150 mm^3^, as measured by MRI (Translational Imaging Shared Resource, Roswell Park). All experimental MRI studies used a 4.7T MR scanner (Roswell Park) dedicated for preclinical research. Baseline MRI scans were acquired prior to treatment. Each KPC mouse was treated daily with 0.5 mg/kg lorazepam or DMSO control (0.2% DMSO in a sodium chloride solution (0.9%), Sigma Cat. #S8776) by intraperitoneal (IP) injection. A 50 μg/mL lorazepam was prepared fresh by diluting a 25 mg/mL stock of lorazepam (Sigma-Aldrich, L1764) dissolved in DMSO in a sodium chloride solution (0.9%), Sigma Cat. #S8776) and each mouse received 0.01 mL/g. Mice were weighed daily and were monitored for hunching, anemia, labored breathing, and decreased activity. Follow-up MR imaging were performed at 1 and 2 weeks to assess tumor growth. Multi-slice high-resolution T2-weighted images were acquired for visualization of tumor extent *in vivo*.

### RNA Sequencing of Subcutaneous Syngeneic KPC Allograft Tumors

Heat maps were generated using a regularized-log transformation (DSEQ2-implement) from raw counts. Each individual gene is row normalized to highlight and examine the differentially expressed genes. Pheatmap package (v1.0.12) from R was used to produce all DE-related heatmaps. As previously described in Venkat et al. (16) Gene Set Enrichment Analysis (GSEA) and Enrichr were used to perform pathway analysis using the MSigDB hallmark, KEGG and Reactome Gene sets (75, 76) (Edward, Subramanian). Enrichment of the input genes (LOR/VEH) in Enrichr was computed using the Fisher’s exact test and p-values were adjusted using the Benjamini-Hochberg correction (FDR < .01).

### RNAscope Multiplex Fluorescent Detection with Immunofluorescence

Tumor processing was performed in the Experimental Tumor Models (ETM) Shared Resource using a HistoCore Arcadia H (Leica) embedder and sliced in 5 µm sections using a RM2235 (Leica) microtome. Chosen slides were warmed at 65°C for 60 min, cooled 10 min, deparaffinized with xylene for 2 x 5 min, dehydrated in 100% ethanol for 2 x 1 minute, and washed with 0.1% Tween-20 RNAse-free 1x phosphate-buffered saline (PBST) three times. RNAscope Multiplex Fluorescent Detection was performed according to modified instructions provided by the Pasca Di Magliano lab (77). Briefly, slides were incubated with hydrogen peroxide (H_2_O_2_) for 10 min at room temperature followed by target retrieval at 98°C for 15 min. Slides were then blocked with Co-Detection antibody diluent for 30 min and incubated with Primary Antibody solutions overnight at 4°C.

The following day, tissue sections were fixed with formalin, treated with Protease Plus Reagent for 11 min at 40°C, and washed with PBST three times. RNAscope probes (if any) were then added for a 2 hr incubation at 40°C. Following two washes with RNAscope wash buffer at each step, signal for each of the probes was amplified with AMP 1, 2, and 3 reagents, horseradish peroxidase, and tyramide signal amplification kit at 40°C. Slides were then stained with DAPI for 15 min at room temperature and incubated with appropriate secondary antibody solution for 45 min at room temperature before being mounted with ProLong Diamond Antifade.

| Antibody/Probe | Company | Catalog # | Dilution |
| --- | --- | --- | --- |
| αSMA | Sigma | A2547 | 1:400 |
| Mm-*Gpr68* | ACD<br>Biosystems | 319321 | According to<br>manufacturer |
| Mm-*Il6*-C2 | ACD<br>Biosystems | 315891-C2 | According to<br>manufacturer |

### RNAscope Imaging Analysis

Images were obtained using confocal microscopy and exported as multiple-image LIFs for analysis in HALO-v3.5 software (Indica Labs). For each slide, five representative confocal microscopy images were obtained, totaling in 10 images. Images were imported directly into the HALO software for analysis. Images were analyzed with HALO image analysis software (Indica labs) using the Indica-Labs FISH-IF module. After cells were detected based on nuclear recognition (DAPI stain), the fluorescence intensity of the cytoplasmic areas of each cell was measured. A mean intensity threshold above background was used to determine positivity for each fluorochrome within the cytoplasm, thereby defining cells as either positive or negative for each marker. The positive cell data were then used to define colocalized populations. The percentage of *α*SMA, *Il6*, and/or *Gpr68* positive cells were calculated by fluorescence positive cell counts, divided by total DAPI positive nuclei. The number of cells was quantified by the HALO programming system and recorded. Percent positive cell values were imported into Excel (Microsoft) for graphing and statistical analysis. Statistics: two-tailed unpaired t test.

### Re-Analysis of Single Cell Sequencing Data

Peng et al. (17) dataset was processed and analysed as described in Venkat et al. (16). In brief, single cell RNA-seq FASTQ files of human PDAC tumors (n= 24) and normal human pancreata (n= 11) were downloaded from the Genome Sequence Archive (GSA) (Accession: CRA001160, Bioproject: PRJCA001063). Files were aligned to the hg19 genome with Cell Ranger 3.1.0 using standard parameters (78). 21 of the human PDAC tumors and all 11 normal human pancreata has proper chemistry and alignment and were used for downstream analyses. Annotated cells with 200-6,000 genes/cell (upper limit to exclude possible doublets) were filtered to remove cells with > 10% mitochondrial counts and genes occurring in < 3 cells, yielding a final count of 10,345 normal pancreas cells and 22,053 PDAC cells. Analyses were carried out in R 4.0.4. Differentially expressed genes between the subclusters were identified using the FindMarkers function in Seurat4 (79).

Steele et al. (20) and Kemp et al.(21) datasets were processed as previously described. In brief, h5 files were imported into R, and processed with the Seurat package (79, 80). Data were normalized and integrated for batch correction. PCA clustering and UMAP visualization was performed to generate unbiased clusters. Populations were labeled based on established lineage markers (20, 21). Feature plots or Dot plots were generated to visualize specific gene expression profiles.

### GPR68 Correlation Analysis

cBioPortal was used to assess GPR68 correlation with CAF and epithelial markers in the Pancreatic Adenocarcinoma (TCGA, Pan-Cancer Atlas) dataset (n=175 patients/samples).

### Measuring pH of Murine Pancreas and Pancreatic Tumors

The fabrication of H+-sensitive microelectrodes and their use for measuring pH was performed as described in detail by Lee et al. (2013) (81). In brief, borosilicate glass (no. BF200-156-10, Sutter Instrument, Novato, CA) is pulled to a fine tip (∼1 megaohm resistance) using a model P-1000 micropipette puller (Sutter Instrument). To create an electrode that monitors the H+-sensitive electrical potential, VH, the tip of one electrode is filled with H+-selective ionophore cocktail B (Sigma Aldrich) and backfilled with a solution: 40 mM K2HPO4, 15 mM NaCl, pH 7.0. To monitor the reference electrical potential, Vref, a second microelectrode is filled with 3 M KCl. The true H+-selective signal is the subtracted signal (VH-Vref), acquired using an HiZ-223 dual channel electrometer (Warner Instruments) and digitized using a Digidata 1550 unit. The signal is converted to pH by a three-point calibration at pH 6.0, pH 7.5 and pH 8.0 using custom software (Courtesy of Dale Huffman and Walter Boron at Case Western Reserve University). The electrical potential of the fluid in the measurement chamber (PBS pH 7.50) is maintained at 0 mV using a bath clamp (no. 725I, Warner Instruments). Pancreatic tissue was sectioned into a 5 mm thick slice to allow submersion in the bath and was impaled with the Vref and VH electrodes. Vref did not deviate from 0 mV, demonstrating electrode placement in the extracellular milieu, while the measured pH dropped rapidly to a new level that plateaued after 5 min.

### Cell Culture

Human immortalized CAF (C7-TA-PSC) cells were a gift from Dr. Edna Cukierman (Fox Chase Cancer Center). HTLA cells were a gift from Dr. Brian Roth (University of North Carolina). All cell lines were routinely tested for mycoplasma using the Genome Modulation Services Shared Resource.

### Acidic Media Preparation

All acidic media preparation was based on a protocol by Dr. Tonio Pera (Thomas Jefferson University).

### HTLA Media

Followed the instructions for powdered DMEM (Sigma Aldrich, Cat. #D5030), when the media was fully dissolved 10% FBS, 12.5 mL 1 M HEPES, 2 μg/mL puromycin, 100 μg/mL hygromycin B, 1 mM sodium pyruvate, and 1% P/S was added to the media. The media was aliquoted into separate beakers and was adjusted to the appropriate pH using 10 N HCl/NaOH. pH was measured with a VWR Traceable pH/ORP meter (10539–802). Media was sterile filtered with a 0.22 μm pore size SteriCup (MilliporeSigma™ Stericup™ Quick Release-GV Vacuum Filtration System, 500 mL, Fisher Scientific, Cat. #S2GVU05RE).

### CAF Media

Followed the instructions for powdered DMEM (Sigma Aldrich D5030), when the media was fully dissolved 10% FBS, 12.5 mL 1 M HEPES, 1mM sodium pyruvate, and 1% P/S was added to the media. The media was aliquoted into separate beakers and was adjusted to the appropriate pH using 10 N HCl/NaOH. pH was measured with a VWR Traceable pH/ORP meter (Cat. #10539-802). Media was sterile filtered with a 0.22 μM pore size SteriCup (MilliporeSigma™ Stericup™ Quick Release-GV Vacuum Filtration System, 500mL, Fisher Scientific, S2GVU05RE).

### PRESTO-Tango Protocol

HTLA cells were maintained in DMEM supplemented with 10% FBS, 2 μg/mL puromycin, 100 μg/mL hygromycin B, and 1% P/S at 37⁰C in a 5% CO_2_ incubator. For acidic pH studies 37⁰C, 0% CO2 incubator was used (see acidic media preparation). For transfection, 400,000 HTLA cells/well were plated in a 6-well dish. The next day, Lipofectamine 3000 (L3000008, Thermo Scientific) was used according to the manufacturer’s instructions to transfect 500 ng GPR68-Tango (Addgene, Cat. #66371) construct per well. The transfection reagent remained on the cells overnight. Three wells were not transfected to serve as a negative control. On day 3, the cells were re-plated in a white flat bottom polystyrene TC-treated Corning 384-well plate (8,000 cells/well). A BioRad TC-20 automated cell counter was used to count the cells. On Day 4, the Tecan D300e digital drug dispenser was used to plate the desired drug concentrations using 10mM drug stocks resuspended in DMSO. DMSO concentration was normalized. On Day 5, the luminescence of each well was measured using Promega Bright-Glo Luciferase Assay System (Cat. #E2610) according to the manufacturer’s instructions.

### Western Blot

Protein lysis was performed following the Silva et al (82) rapid extraction method for mammalian cell culture. Proteins were transferred to nitrocellulose membranes (0.2 µm, Bio-Rad, Cat. #1620112) at a constant voltage of 100 V for 70 min at 4°C using Mini Trans-Blot® Cell (Bio-Rad). Membranes were blocked in TBS-T (Tris-buffered saline (TBS) with 0.5% v/v TWEEN-20, Sigma Aldrich) and 5% w/v non-fat dry milk (Blotting-Grade Blocker, Bio-Rad, Cat. #1706404). Primary antibodies were diluted in 5% milk in TBS-T and incubated overnight at 4°C phospho-CREB (Ser133) (87G3) monoclonal anti-rabbit antibody (Cell Signaling Technology, Cat. #9198S, 1:1,000 dilution), GAPDH anti-mouse monoclonal antibody (Proteintech, Cat. #60004-1-Ig, 1:20,000 dilution). Membranes were incubated with horseradish peroxidase-conjugated secondary antibodies (1:2,000 Donkey anti-rabbit; Fisher Scientific; Cat. #45-000-682, or 1:2,000 Goat anti-mouse Sigma Aldrich Cat. #A4416) for 45-90 min at room temperature. Pierce ECL Western Blotting Substrate (Thermo Scientific, Cat. #32106) was used for chemiluminescent detection. Signals were visualized and imaged using the ChemiDoc XRS+ System and Image Lab Software (Bio-Rad).

### qPCR

Cells were washed once with ice cold PBS then lysed and homogenized in TRIzol reagent according to the manufacturer’s protocol. RNA was isolated and DNase I treated using a Direct-zol RNA miniprep kit (Zymo research) according to the manufacturer’s protocol. RNA concentration and purity were measured using a Thermo Scientific NanoDrop 8000 Spectrophotometer. Any RNA with an A260/280 ratio below 1.9 or an A260/230 ratio below 1.9 were excluded from the analysis. RNA was aliquoted and stored at −80°C. 300-900 ng RNA was converted to 20 μL cDNA using the iScript cDNA synthesis kit (BioRad) according to the manufacturer’s instructions. The cDNA was diluted with nuclease-free water (∼15 ng/μL) and the qPCR was performed in 10 μL reactions using iTaq Universal SYBR green Supermix according to the manufacturer’s instructions using 0.5 μL primer and 1 μL cDNA per reaction. Thermal cycling was performed using a BioRad CFX Connect Realtime System. All primers were BioRad PrimePCR SYBR Green Assay primers. Gene expression analysis was performed using the ΔΔCt method.

### CRISPRi GPR68 knockdown cell generation

The knockdown cells were generated according to a modified protocol from Francescone et al. 2021 (83). The following GPR68 CRISPRi gRNA sequences were used (gRNA sequences were selected from the top guide RNA sequences for GPR68 as determined by Horlbeck et al. 2016 (84)):

1.1 **<u>CACC</u>**GGGAGGGAGAGCTGGGATCG
1.2 **<u>AAAC</u>**CGATCCCAGCTCTCCCTCCC

#### Generation of lentiviral vectors

Designed guide sequences (Integrated DNA Technologies) were cloned into the lentiviral vector CRISPRi-Puro (gifted from the Cukierman Lab: modified from Addgene Plasmid #71236 to contain a “stuffer” to promote gRNA cloning efficiency). 8 μg CRISPRi-Puro plasmid was linearized and dephosphorylated with 2 μL BSMBI enzyme and 5 μL NE buffer 3.1 diluted in distilled water for a final volume of 50 μL. The mixture was placed into Eppendorf Thermomixer C (55°C, 300 rpm, 3 hr) then 1 μL of CIP was added and incubated for 1 hr (55°C, 300rpm). After linearization, the digested plasmid was loaded into an agarose gel (0.6%) and the higher molecular weight band was gel purified using an Invitrogen PureLink Quick Gel Extraction Kit (Thermo Fisher Scientific) according to the manufacturer’s instructions. The guide RNA oligos were phosphorylated and annealed: 1 μL Oligo 1 (100 uM), 1 μL Oligo 2 (100 uM), 1 μL 10x T4 ligation buffer (NEB), 6.5 μL ddH20, and 0.5 μL T4 PNK (10 μL total volume). The phosphorylation/annealing mixture was placed into the BioRad T100 Thermocycler: 37°C (30 min), 95°C (5 min), then ramped down to 25°C at 5°C/min, then diluted 1:200 with ddH20. The annealed and phosphorylated guide sequences were ligated into the linearized and dephosphorylated CRISPRi-Puro plasmid as follows: 25 ng linearized CRISPRi-Puro plasmid, 1 μL 1:200 annealed guides, 1 μL 10x T4 ligase buffer, and 1 μL T4 ligase (10 μL total volume) was incubated at room temperature for 30 min. 3 μL of the ligation reaction was transformed into 25 μL of Stbl3 competent cells (NEB) by keeping the mixture on ice for 10 min, heat shocking at 42°C for ∼1 minute, placing on ice for 10 min, adding 100 μL sterile LB to each tube, and incubating for 30 min in the Eppendorf Thermomixer C (37°C, 300 rpm). The entire mixture was plated on LB-AMP plates (100 μg/mL Ampicillin), 2-3 colonies from each plate were miniprepped using the Thermo Scientific GeneJet miniprep kit according to the manufacturer’s instructions. The plasmid DNA was sequenced by Eurofins Genomics using the hU6-F primer: GAGGGCCTATTTCCCATGATT. Lentiviruses were produced as follows: <u>Day 1</u> transfect 293T cells (∼75% confluent, 10 cm plate, 6 mL fresh complete media) with 2 μg of the CRISPRi-Puro plasmid containing the appropriate guide (and CRISPRi-Puro uncut as a control), 1.5 μg psPAX2, and 0.5 μg pMD2.G using Lipofectamine 3000 according to the manufacturer’s instructions. <u>Day 2</u> Gently add 4 mL fresh complete media to each plate and incubate for 24 hr. <u>Day 3</u> Collect virus and replace with 10 mL fresh media, filter (0.45 μm), aliquot, and store in −80°C. <u>Day 4</u> Collect virus, filter (0.45 μm), aliquot, and store in −80°C.

#### Lentiviral reverse transduction (based on Addgene protocol)

60,000 C7-TA-PSC cells per mL of media containing 10 μg/mL polybrene were prepared. Lentiviral media was rapidly thawed, diluted, and mixed with 60,000 cells in 1 mL of media, the virus was left on the cells for 48 hr and then replaced with fresh complete media. A no virus control was made for selection purposes. 72 hr after the reverse transduction puromycin selection was performed (2 μg/mL).

### Human IL-6 ELISA

For the GPR68 overexpression ELISA <u>Day 1</u>: 1 mL of media containing 28,000 C7-TA-PSC immortalized human CAFs were plated into each well of a 12-well plate. <u>Day 2</u>: Wells were transfected with 125 ng GPR68 cDNA or a no DNA control using Lipofectamine 3000 according to the manufacturer’s instructions for a 12-well plate. <u>Day 3</u>: 20 μM benzodiazepine/DMSO control were bulk prepared in pH 6.8 media and 1 mL per well was added (24 hr timepoint), 6 hr timepoint wells received pH 6.8 media, the plate was kept in the 37°C, 0% CO2 incubator. <u>Day 4</u>: 20 μM benzodiazepine/DMSO control were bulk prepared in pH 6.8 media and 1 mL per well was added to the 6 hr timepoint wells, the plate was kept in the 37°C, 0% CO2 incubator. The media was collected from the wells, centrifuged at 1,000 rpm, 4°C, 3 min, and the supernatant was transferred to freshly labelled tubes. 100 μL of each sample as well as 100 μL of each standard (0-1,000 pg/mL, prepared according to the manufacturer’s instructions for cell culture supernatants) were plated into the wells of the ELISA test strips and incubated overnight, 4°C, with gentle rocking (Sigma-Aldrich, RAB0306, Human IL-6 ELISA Kit). <u>Day 5</u>: Finished ELISA according to the manufacturer’s instructions. For the ELISAs without GPR68 expression 45,000-50,000 C7-TA-PSC immortalized human CAF cells per well were plated in 12-well plates, 20 μM BZDs were added on Day 2, 24 hr later the conditioned media was collected and centrifuged, as described above. Statistics: One or two-way ANOVA with Bonferroni multiple comparisons or Holm-Šídák’s multiple comparisons test, respectively.

### Pan-Cancer Epidemiology Study

All statistics were performed using SAS version 9.4 (SAS Institute Inc., Cary, NC). All analyses were performed within disease site (brain, breast, corpus uteri, head and neck, melanoma, kidney, ovary, pancreas, colon, and prostate). Only patients with a diagnostic date starting at the year 2000 were used for this analysis. Within disease site, patient characteristics were summarized by cohort (LOR, ALP, No Benzo). Frequencies and relative frequencies were provided for categorical variables and compared using chi-square test. P-values were provided. The overall and progression-free survival summaries were summarized by cohort using standard Kaplan-Meier methods. The median survival rate, Kaplan-Meier curves, and log-rank p-values were provided. Time to progression was calculated from ‘recurrence days from Dx’ if recurrence occurred. Otherwise, overall survival time was used. Multivariate Cox regression modelling was performed to measure associations between survival outcomes and cohort. Models were adjusted for sex (where applicable), clinical grade, and clinical stage. Hazard ratios and corresponding 95% confidence intervals were provided for individual LOR and ALP groups, with ‘No Benzo’ as the referent group. Type 3 Test was used and an overall p-value measuring the association between survival and cohort was provided.

### Statistical Analysis

Statistics were performed in GraphPad Prism 9.3.1. Unless otherwise noted, p < 0.05 was considered statistically significant. All statistical methods and p-values are described in the figure legends. Asterisks on the graphs denote statistically significant differences: * represents p-values < 0.05, ** represents p-values < 0.01, *** represents p-values < 0.001, **** represents p-values < 0.0001.

## Data Availability

All data and code will be available at https://github.com/feiginlab.

## Acknowledgments

We thank Dr. Ralph Francescone for providing advice and reagents for generating the CRISPRi knockdown cell lines, Dr. Agnieszka Witkiewicz for providing human primary stellate cells, Michael Habitzruther for performing KPC murine imaging, Dr. Tonio Pera for protocols to prepare acidic pH media, and Richard A. Pasternack for technical support with pH measurements. Research reported in this publication was supported by Roswell Park Comprehensive Cancer Center and the National Cancer Institute (NCI) of the National Institutes of Health (NIH) under Award Numbers P30CA016056 and F31CA260942, and by seed funding from the Roswell Park Alliance Foundation. NCI grant P30CA016056 supported the use of Roswell Park Comprehensive Cancer Center’s Pathology Network, Biomedical Research Informatics, Biostatistics and Statistical Genomics, Genome Modulation Services, Translational Imaging, Experimental Tumor Models, Drug Discovery Core, and Bioanalytics, Metabolomics, and Pharmacokinetics Shared Resources. Additional support from NIH (R01EY028580, to M.P) and the 5th AHEPA Cancer Research Foundation Inc, as well as R01CA269660, S10ODO23666 and P30CA06927 (Core Microscopy Facility) to J.F.B and E.C. C.F receives funding from the National Comprehensive Cancer Network Foundation, National Comprehensive Cancer Network Oncology Research Program, Taiho Oncology, and Pfizer Inc (all to RPCC).

## Tables

Table 1. Pancreatic cancer patient characteristics by benzodiazepine prescription records

Table 2. Pancreatic cancer patient characteristics by lorazepam prescription records

Table 3. Pancreatic cancer patient characteristics by alprazolam prescription records

Table 4. Pancreatic cancer time-to-event outcomes: Multivariate Summaries

Table 5. Brain cancer patient characteristics, median survival, and Multivariate Cox Regression modeling

Table 6. Breast cancer patient characteristics, median survival, and Multivariate Cox Regression modeling

Table 7. Corpus uterine cancer patient characteristics, median survival, and Multivariate Cox Regression modeling

Table 8. Head and neck cancer patient characteristics, median survival, and Multivariate Cox Regression modeling

Table 9. Invasive nevi/melanoma patient characteristics, median survival, and Multivariate Cox Regression modeling

Table 10. Renal cancer patient characteristics, median survival, and Multivariate Cox Regression modeling

Table 11. Ovarian cancer patient characteristics, median survival, and Multivariate Cox Regression modeling

Table 12. Colon cancer patient characteristics, median survival, and Multivariate Cox Regression modeling

Table 13. Prostate cancer patient characteristics, median survival, and Multivariate Cox Regression modeling

## Supplemental Figure Legends

**Supplemental Figure S1. (A)** Number of patients by cancer type with a record of BZD prescriptions by cancer type in males (black bar, top) and females (pink bar, bottom). **(B)** Kaplan Meier curve comparing progression-free survival of pancreatic cancer patients at Roswell Park from 2004-2020 with a record of chemotherapy with (n=69) or without (n=219) a prescription record of BZDs (excluding midazolam). **(C)** Kaplan Meier curve comparing disease-specific survival of pancreatic cancer patients at Roswell Park from 2004-2020 with a record of chemotherapy with (n=357) or without (n=1093) a prescription record of BZDs (excluding midazolam). **(D)** Covariate adjusted analysis evaluating the impact of BZD prescription records on pancreatic cancer patient disease-specific survival accounting for age, sex, race, clinical stage, additional treatments, and progressive disease. **(E)** Percentage of invasive nevi/melanoma, brain, breast, colon, corpus uteri, head and neck, kidney, ovarian, pancreatic, and prostate cancer patients prescribed BZDs that are receiving midazolam. <u>Statistics</u>: See Figure 1.

**Supplemental Figure S2. (A)** Experimental schematic of long-term ALP (n=6), LOR (n=6), or vehicle-treatment (n=6) in the subcutaneous KPC syngeneic allograft model. **(B-D)** Enrollment (B) age, (C) weight, and (D) tumor volume in the long-term ALP, LOR, or vehicle-treatment in the subcutaneous KPC syngeneic allograft experiment. **(E-K)** (E) Tumor growth curves, (F) endpoint tumor weight, (G) Kaplan Meier curves, (H) representative 20x H&E images, (I) quantification of the percentage of necrotic area per slide in a blinded manner by a pathologist, (J) representative Masson’s trichrome images, (K) quantification of the percentage of collagen per slide in a blinded manner by a pathologist of the long-term LOR, ALP, or vehicle treatment study. **(L-N)** Enrollment (L) age, (M), weight, and (N) tumor size at enrollment in the short-term LOR subcutaneous KPC syngeneic allograft experiment (n=4-5/arm). **(O,P)** (O) Endpoint tumor weight and (P) tumor growth curves in the short-term LOR study. **(Q)** Representative 20x Ki67 IHC images of the edge of 1-week (left) and 2-week (right) treated LOR tumors. **(R-U)** Second harmonic generation (SHG) imaging of the two-week treated mice in the short-term LOR study (n=3/arm): (R) Integrated density, (S) collagen fiber length, (T) collagen fiber width, and (U) collagen fiber straightness. <u>Statistics</u>: For analysis of multiple groups, ordinary one-way ANOVA with Tukey’s multiple comparison test. In the case multiple groups with two independent variables, groups were compared by mixed effects analysis with Bonferroni’s multiple comparison test.

**Supplemental Figure S3. (A)** Representative 40x RNAscope images of IL6+/SMA+ cells in the two-week treated vehicle (left) and LOR-treated KPC tumors. **(B)** Quantification of (A).

**Supplemental Figure S4. (A)** Heat map of GABA receptor expression by cell type from the Peng *et al.* (17) human pancreatic ductal adenocarcinoma tumor single cell sequencing dataset. **(B)** Structure of lorazepam (left) and alprazolam (right), pink circle denotes the n-unsubstitution and the teal circle denotes the n-substitution. **(C)** Dot plot visualization of *Gpr68* gene expression level (color intensity) and frequency (size of dot) in different cell populations of murine PDAC samples from Kemp *et al.* (21). **(D)** Representative images of *Gpr68* expression by RNAscope in the two-week vehicle (left) treated subcutaneous KPC tumors and the KPC spontaneous tumor (right). **(E-G)** UMAP plots of (E) *GPR68*, (F) *RGS5* (pericyte marker), and (G) *DCN* (pan-CAF marker) in human PDAC CAF cluster reprocessed from the Steele *et al.* (20) single cell sequencing dataset. **(H-J)** UMAP plots of (H) DCN, (I) RGS5, and (J) *GPR68* expression in (left to right) normal human pancreas, PDAC metastasis, and primary PDAC tumors from the Steele *et al.* (20) single cell sequencing dataset. **(K,L)** Representative extracellular pH tracing of (K) Normal C57BL/6 murine pancreas and (L) Murine PDAC tumors isolated from C57BL/6 mouse subcutaneously implanted bilaterally with KPC tumor chunks. **(M)** Scatterplot with bar of the extracellular pH values from normal pancreas (n=2 biological replicates) and KPC subcutaneous tumors (n=4 biological replicates), dots represent independent pH readings. **(N)** Representative 10x H&E image of the subcutaneous KPC tumor from the pH experiment. **(O,P)** Representative extracellular pH tracing of (O) adjacent normal pancreas from C57BL/6 mice with orthotopically implanted KPC tumor pieces and (P) Murine PDAC tumors isolated from C57BL/6 mouse orthotopically implanted bilaterally with KPC tumor pieces. **(Q)** Scatterplot with bar of the extracellular pH values from normal pancreas (n=2 biological replicates) and KPC orthotopic tumors (n=2 biological replicates), dots represent independent pH readings. **(R)** Representative 10x H&E image of the orthotopic KPC tumor from the pH experiment. <u>Statistics</u>: pH values were compared using unpaired one-tailed t-tests.

**Supplemental Figure S5. (A)** *GPR68* knockdown by CRISPRi in human immortalized CAFs. **(B)** *IL6* mRNA expression in control and *GPR68* knockdown CAFs by qPCR.

**Supplemental Figure S6. (A,B)** Percentage of invasive nevi/melanoma, brain, breast, colon, corpus uteri, head and neck, kidney, ovarian, pancreatic, and prostate cancer patients prescribed or infused BZDs that are receiving (A) LOR or (B) LOR. Pan-cancer analyses refers to the combined average of cancer types in the nSight database. **(C,D)** Association between prescription records of (C) LOR or (D) ALP and PFS by cancer type. **(E-G)** Kaplan Meier curve comparing PFS in Roswell Park patients with prescription records of LOR or ALP, or those with no history of BZD use treated for primary (E) invasive nevi or melanoma, (F) prostate cancer, or (G) ovarian cancer. <u>Statistics</u>: Multivariate Cox regression modeling was performed to measure associations between survival outcomes and cohort. Models were adjusted for sex (where applicable), clinical grade, and clinical stage. HRs and corresponding 95% CIs were provided for individual LOR and ALP groups, with ‘No Benzo’ as the referent group. Type 3 Test was used and an overall p-value measuring the association between survival and cohort was provided. CI: confidence interval, HR: Hazard ratio, PFS: progression-free survival.

## Notes

### Competing Interest Statement

The authors have declared no competing interest.

### Author Declarations

IRB of Roswell Park Comprehensive Cancer Center gave ethical approval for this work.

## References

1. SEER*Explorer: An interactive website for SEER cancer statistics, National Cancer Institute [Internet]2022. Available from: https://seer.cancer.gov/statistics-network/explorer/application.html.

2. Rahib L, Smith BD, Aizenberg R, Rosenzweig AB, Fleshman JM, Matrisian LM. Projecting cancer incidence and deaths to 2030: the unexpected burden of thyroid, liver, and pancreas cancers in the United States. Cancer research. 2014;74(11):2913–21.

3. Sarantis P, Koustas E, Papadimitropoulou A, Papavassiliou AG, Karamouzis MV. Pancreatic ductal adenocarcinoma: Treatment hurdles, tumor microenvironment and immunotherapy. World journal of gastrointestinal oncology. 2020;12(2):173.

4. Wu YA, Oba A, Lin R, Watanabe S, Meguid C, Schulick RD, Del Chiaro M. Selecting surgical candidates with locally advanced pancreatic cancer: a review for modern pancreatology. Journal of Gastrointestinal Oncology. 2021;12(5):2475.

5. Hwang RF, Moore T, Arumugam T, Ramachandran V, Amos KD, Rivera A, Ji B, Evans DB, Logsdon CD. Cancer-associated stromal fibroblasts promote pancreatic tumor progression. Cancer research. 2008;68(3):918–26.

6. Sahai E, Astsaturov I, Cukierman E, DeNardo DG, Egeblad M, Evans RM, Fearon D, Greten FR, Hingorani SR, Hunter T. A framework for advancing our understanding of cancer-associated fibroblasts. Nature Reviews Cancer. 2020;20(3):174–86.

7. Öhlund D, Handly-Santana A, Biffi G, Elyada E, Almeida AS, Ponz-Sarvise M, Corbo V, Oni TE, Hearn SA, Lee EJ. Distinct populations of inflammatory fibroblasts and myofibroblasts in pancreatic cancer. Journal of Experimental Medicine. 2017;214(3):579–96.

8. Mitsunaga S, Ikeda M, Shimizu S, Ohno I, Furuse J, Inagaki M, Higashi S, Kato H, Terao K, Ochiai A. Serum levels of IL-6 and IL-1β can predict the efficacy of gemcitabine in patients with advanced pancreatic cancer. British journal of cancer. 2013;108(10):2063.

9. Miller K, Massie MJ. Depression and anxiety. The Cancer Journal. 2006;12(5):388–97.

10. Bektay MY, İzzettin FV. Oncology Pharmacy Practice: The Clinical Pharmacist’s Perspective2021.

11. Cornwell AC, Feigin ME. Unintended effects of GPCR-targeted drugs on the cancer phenotype. Trends in Pharmacological Sciences. 2020.

12. Tradounsky G. Seizures in palliative care. Canadian Family Physician. 2013;59(9):951–5.

13. Howard P, Twycross R, Shuster J, Mihalyo M, Wilcock A. Benzodiazepines. Journal of pain and symptom management. 2014;47(5):955–64.

14. Tang X, Yang L, Fishback NF, Sanford LD. Differential effects of lorazepam on sleep and activity in C57BL/6J and BALB/cJ strain mice. Journal of sleep research. 2009;18(3):365–73.

15. Miller LG, Greenblatt DJ, Paul SM, Shader RI. Benzodiazepine receptor occupancy in vivo: correlation with brain concentrations and pharmacodynamic actions. Journal of Pharmacology and Experimental Therapeutics. 1987;240(2):516–22.

16. Venkat S, Feigin ME. Alternative polyadenylation characterizes epithelial and fibroblast phenotypic heterogeneity in pancreatic ductal adenocarcinoma. bioRxiv. 2021.

17. Peng J, Sun B-F, Chen C-Y, Zhou J-Y, Chen Y-S, Chen H, Liu L, Huang D, Jiang J, Cui G-S. Single-cell RNA-seq highlights intra-tumoral heterogeneity and malignant progression in pancreatic ductal adenocarcinoma. Cell research. 2019;29(9):725–38.

18. Wiley SZ, Sriram K, Liang W, Chang SE, French R, McCann T, Sicklick J, Nishihara H, Lowy AM, Insel PA. GPR68, a proton-sensing GPCR, mediates interaction of cancer-associated fibroblasts and cancer cells. The FASEB Journal. 2018;32(3):1170–83.

19. Huang X-P, Karpiak J, Kroeze WK, Zhu H, Chen X, Moy SS, Saddoris KA, Nikolova VD, Farrell MS, Wang S. Allosteric ligands for the pharmacologically dark receptors GPR68 and GPR65. Nature. 2015;527(7579):477.

20. Steele NG, Carpenter ES, Kemp SB, Sirihorachai VR, The S, Delrosario L, Lazarus J, Amir E-aD, Gunchick V, Espinoza C. Multimodal mapping of the tumor and peripheral blood immune landscape in human pancreatic cancer. Nature Cancer. 2020;1(11):1097–112.

21. Kemp SB, Steele NG, Carpenter ES, Donahue KL, Bushnell GG, Morris AH, Orbach SM, Sirihorachai VR, Nwosu ZC, Espinoza C. Pancreatic cancer is marked by complement-high blood monocytes and tumor-associated macrophages. Life science alliance. 2021;4(6).

22. Insel PA, Sriram K, Wiley SZ, Wilderman A, Katakia T, McCann T, Yokouchi H, Zhang L, Corriden R, Liu D. GPCRomics: GPCR expression in cancer cells and tumors identifies new, potential biomarkers and therapeutic targets. Frontiers in pharmacology. 2018;9.

23. Michalaki V, Syrigos K, Charles P, Waxman J. Serum levels of IL-6 and TNF-α correlate with clinicopathological features and patient survival in patients with prostate cancer. British journal of cancer. 2004;90(12):2312–6.

24. Lane D, Matte I, Rancourt C, Piché A. Prognostic significance of IL-6 and IL-8 ascites levels in ovarian cancer patients. BMC cancer. 2011;11(1):1–6.

25. Duffy SA, Taylor JM, Terrell JE, Islam M, Li Y, Fowler KE, Wolf GT, Teknos TN. Interleukin-6 predicts recurrence and survival among head and neck cancer patients. Cancer: Interdisciplinary International Journal of the American Cancer Society. 2008;113(4):750–7.

26. Stark DPH, House A. Anxiety in cancer patients. British journal of cancer. 2000;83(10):1261–7.

27. Zabora J, BrintzenhofeSzoc K, Curbow B, Hooker C, Piantadosi S. The prevalence of psychological distress by cancer site. Psycho-Oncology: Journal of the Psychological, Social and Behavioral Dimensions of Cancer. 2001;10(1):19–28.

28. Green AI, Austin CP. Psychopathology of pancreatic cancer: a psychobiologic probe. Psychosomatics. 1993;34(3):208–21.

29. Clark KL, Loscalzo M, Trask PC, Zabora J, Philip EJ. Psychological distress in patients with pancreatic cancer—an understudied group. Psycho-oncology. 2010;19(12):1313–20.

30. Wilson KG, Chochinov HM, Skirko MG, Allard P, Chary S, Gagnon PR, Macmillan K, De Luca M, O’Shea F, Kuhl D. Depression and anxiety disorders in palliative cancer care. Journal of pain and symptom management. 2007;33(2):118–29.

31. Kripke DF, Langer RD, Kline LE. Hypnotics’ association with mortality or cancer: a matched cohort study. BMJ open. 2012;2(1):e000850.

32. Kao C-H, Sun L-M, Su K-P, Chang S-N, Sung F-C, Muo C-H, Liang J-A. Benzodiazepine use possibly increases cancer risk: a population-based retrospective cohort study in Taiwan. J Clin Psychiatry. 2012;73(4):e555–e60.

33. Iqbal U, Nguyen P-A, Syed-Abdul S, Yang H-C, Huang C-W, Jian W-S, Hsu M-H, Yen Y, Li Y-CJ. Is long-term use of benzodiazepine a risk for cancer? Medicine. 2015;94(6).

34. Harlow BL, Cramer DW. Self-reported use of antidepressants or benzodiazepine tranquilizers and risk of epithelial ovarian cancer: evidence from two combined case-control studies (Massachusetts, United States). Cancer Causes & Control. 1995;6(2):130–4.

35. Pottegård A, Friis S, Andersen M, Hallas J. Use of benzodiazepines or benzodiazepine related drugs and the risk of cancer: a population-based case-control study. British journal of clinical pharmacology. 2013;75(5):1356–64.

36. Kim HB, Myung SK, Park YC, Park B. Use of benzodiazepine and risk of cancer: A meta-analysis of observational studies. International journal of cancer. 2017;140(3):513–25.

37. Diwan BA, Rice JM, Ward JM. Tumor-promoting activity of benzodiazepine tranquilizers, diazepam and oxazepam, in mouse liver. Carcinogenesis. 1986;7(5):789–94.

38. Miyawaki I, Moriyasu M, Funabashi H, Yasuba M, Matsuoka N. Mechanism of clobazam-induced thyroidal oncogenesis in male rats. Toxicology letters. 2003;145(3):291–301.

39. Fox K, Lahcen R. Liver-cell adenomas and peliosis hepatis in mice associated with oxazepam. Research communications in chemical pathology and pharmacology. 1974;8(3):481–8.

40. O’Donnell SB, Nicholson MK, Boland JW. The association between benzodiazepines and survival in patients with cancer: a systematic review. Journal of Pain and Symptom Management. 2019;57(5):999–1008. e11.

41. Oshima Y, Sano M, Kajiwara I, Ichimaru Y, Itaya T, Kuramochi T, Hayashi E, Kim J, Kitajima O, Masugi Y. Midazolam exhibits antitumour and anti-inflammatory effects in a mouse model of pancreatic ductal adenocarcinoma. British journal of anaesthesia. 2022;128(4):679–90.

42. Fafalios A, Akhavan A, Parwani AV, Bies RR, McHugh KJ, Pflug BR. Translocator protein blockade reduces prostate tumor growth. Clinical Cancer Research. 2009;15(19):6177–84.

43. Freiregarabal M, Nuneziglesias M, Balboa J, Fernandezrial J, Garciavallejo L, Gonzalezpatino E, Reymendez M. Inhibitory effects of alprazolam on the enhancement of lung metastases induced by operative stress in rats. International Journal of Oncology. 1993;3(3):513–7.

44. Fride E, Skolnick P, Arora PK. Immunoenhancing effects of alprazolam in mice. Life sciences. 1990;47(26):2409–20.

45. Freire-Garabal M, Núñez MJ, Balboa J, Fernández-Rial J, Vallejo LG, González-Bahillo J, Rey-Méndez M. Effects of alprazolam on cellular immune response to surgical stress in mice. Cancer letters. 1993;73(2-3):155–60.

46. Elmesallamy G, Abass MA, Atta A, Refat NA. Differential effects of alprazolam and clonazepam on the immune system and blood vessels of non-stressed and stressed adult male albino rats. Mansoura Journal of Forensic Medicine and Clinical Toxicology. 2011;19(2):1–25.

47. Covelli V, Maffione AB, Greco B, Cannuscio B, Calvello R, Jirillo E. In vivo effects of alprazolam and lorazepam on the immune response in patients with migraine without aura. Immunopharmacology and immunotoxicology. 1993;15(4):415–28.

48. Ramirez K, Niraula A, Sheridan JF. GABAergic modulation with classical benzodiazepines prevent stress-induced neuro-immune dysregulation and behavioral alterations. Brain, behavior, and immunity. 2016;51:154–68.

49. Zhang Y, Yan W, Collins MA, Bednar F, Rakshit S, Zetter BR, Stanger BZ, Chung I, Rhim AD, di Magliano MP. Interleukin-6 is required for pancreatic cancer progression by promoting MAPK signaling activation and oxidative stress resistance. Cancer research. 2013.

50. Mace TA, Shakya R, Pitarresi JR, Swanson B, McQuinn CW, Loftus S, Nordquist E, Cruz-Monserrate Z, Yu L, Young G. IL-6 and PD-L1 antibody blockade combination therapy reduces tumour progression in murine models of pancreatic cancer. Gut. 2018;67(2):320–32.

51. Biffi G, Oni TE, Spielman B, Hao Y, Elyada E, Park Y, Preall J, Tuveson DA. IL1-Induced JAK/STAT Signaling Is Antagonized by TGFβ to Shape CAF Heterogeneity in Pancreatic Ductal AdenocarcinomaPathway Antagonism Shapes CAF Heterogeneity in PDAC. Cancer discovery. 2019;9(2):282–301.

52. Campo-Soria C, Chang Y, Weiss DS. Mechanism of action of benzodiazepines on GABAA receptors. British journal of pharmacology. 2006;148(7):984–90.

53. Lafleur J. Drug Class Review Benzodiazepines in the Treatment of Anxiety Disorder2016.

54. Brunetti P, Giorgetti R, Tagliabracci A, Huestis MA, Busardò FP. Designer benzodiazepines: a review of toxicology and public health risks. Pharmaceuticals. 2021;14(6):560.

55. Song D-K, Suh H-W, Huh S-O, Jung J-S, Ihn B-M, Choi I-G, Kim Y-H. Central GABAA and GABAB receptor modulation of basal and stress-induced plasma interleukin-6 levels in mice. Journal of Pharmacology and Experimental Therapeutics. 1998;287(1):144–9.

56. Miyawaki T, Sogawa N, Maeda S, Kohjitani A, Shimada M. Effect of midazolam on interleukin-6 mRNA expression in human peripheral blood mononuclear cells in the absence of lipopolysaccharide. Cytokine. 2001;15(6):320–7.

57. Horman SR, To J, Lamb J, Zoll JH, Leonetti N, Tu B, Moran R, Newlin R, Walker JR, Orth AP. Functional profiling of microtumors to identify cancer associated fibroblast-derived drug targets. Oncotarget. 2017;8(59):99913.

58. Chandra V, Karamitri A, Richards P, Cormier F, Ramond C, Jockers R, Armanet M, Albagli-Curiel O, Scharfmann R. Extracellular acidification stimulates GPR68 mediated IL-8 production in human pancreatic β cells. Scientific reports. 2016;6:25765.

59. Ichimonji I, Tomura H, Mogi C, Sato K, Aoki H, Hisada T, Dobashi K, Ishizuka T, Mori M, Okajima F. Extracellular acidification stimulates IL-6 production and Ca2+ mobilization through proton-sensing OGR1 receptors in human airway smooth muscle cells. American Journal of Physiology-Lung Cellular and Molecular Physiology. 2010;299(4):L567–L77.

60. Horiguchi K, Higuchi M, Yoshida S, Nakakura T, Tateno K, Hasegawa R, Takigami S, Ohsako S, Kato T, Kato Y. Proton receptor GPR68 expression in dendritic-cell-like S100β-positive cells of rat anterior pituitary gland: GPR68 induces interleukin-6 gene expression in extracellular acidification. Cell and tissue research. 2014;358(2):515–25.

61. High RA, Randtke EA, Jones KM, Lindeman LR, Ma JC, Zhang S, LeRoux LG, Pagel MD. Extracellular acidosis differentiates pancreatitis and pancreatic cancer in mouse models using acidoCEST MRI. Neoplasia. 2019;21(11):1085–90.

62. Zhu S, Zhou H-Y, Deng S-C, Deng S-J, He C, Li X, Chen J-Y, Jin Y, Hu Z-L, Wang F. ASIC1 and ASIC3 contribute to acidity-induced EMT of pancreatic cancer through activating Ca2+/RhoA pathway. Cell death & disease. 2017;8(5):e2806-e.

63. Lardner A. The effects of extracellular pH on immune function. Journal of leukocyte biology. 2001;69(4):522–30.

64. Huber V, Camisaschi C, Berzi A, Ferro S, Lugini L, Triulzi T, Tuccitto A, Tagliabue E, Castelli C, Rivoltini L, editors. Cancer acidity: An ultimate frontier of tumor immune escape and a novel target of immunomodulation. Seminars in cancer biology; 2017: Elsevier.

65. Swietach P, Vaughan-Jones RD, Harris AL, Hulikova A. The chemistry, physiology and pathology of pH in cancer. Philosophical Transactions of the Royal Society B: Biological Sciences. 2014;369(1638):20130099.

66. Hutter S, van Haaften WT, Hünerwadel A, Baebler K, Herfarth N, Raselli T, Mamie C, Misselwitz B, Rogler G, Weder B. Intestinal Activation of pH-Sensing Receptor OGR1 [GPR68] Contributes to Fibrogenesis. Journal of Crohn’s and Colitis. 2018;12(11):1348–58.

67. Matsuzaki S, Ishizuka T, Yamada H, Kamide Y, Hisada T, Ichimonji I, Aoki H, Yatomi M, Komachi M, Tsurumaki H. Extracellular acidification induces connective tissue growth factor production through proton-sensing receptor OGR1 in human airway smooth muscle cells. Biochemical and biophysical research communications. 2011;413(4):499–503.

68. Zhu H, Guo S, Zhang Y, Yin J, Yin W, Tao S, Wang Y, Zhang C. Proton-sensing GPCR-YAP signalling promotes cancer-associated fibroblast activation of mesenchymal stem cells. International journal of biological sciences. 2016;12(4):389.

69. Martin AL, Steurer MA, Aronstam RS. Constitutive activity among orphan class-A G protein coupled receptors. PloS one. 2015;10(9):e0138463.

70. Xu J, Mathur J, Vessières E, Hammack S, Nonomura K, Favre J, Grimaud L, Petrus M, Francisco A, Li J. GPR68 Senses Flow and Is Essential for Vascular Physiology. Cell. 2018;173(3):762–75. e16.

71. Wei W-C, Bianchi F, Wang Y-K, Tang M-J, Ye H, Glitsch MD. Coincidence Detection of Membrane Stretch and Extracellular pH by the Proton-Sensing Receptor OGR1 (GPR68). Current Biology. 2018;28(23):3815–23. e4.

72. Ruggeri JM, Franco-Barraza J, Sohail A, Zhang Y, Long D, di Magliano MP, Cukierman E, Fridman R, Crawford HC. Discoidin domain receptor 1 (DDR1) is necessary for tissue homeostasis in pancreatic injury and pathogenesis of pancreatic ductal adenocarcinoma. The American Journal of Pathology. 2020;190(8):1735-51.

73. Schindelin J, Arganda-Carreras I, Frise E, Kaynig V, Longair M, Pietzsch T, Preibisch S, Rueden C, Saalfeld S, Schmid B. Fiji: an open-source platform for biological-image analysis. Nature methods. 2012;9(7):676–82.

74. Bredfeldt JS, Liu Y, Pehlke CA, Conklin MW, Szulczewski JM, Inman DR, Keely PJ, Nowak RD, Mackie TR, Eliceiri KW. Computational segmentation of collagen fibers from second-harmonic generation images of breast cancer. Journal of biomedical optics. 2014;19(1):016007-.

75. Subramanian A, Tamayo P, Mootha VK, Mukherjee S, Ebert BL, Gillette MA, Paulovich A, Pomeroy SL, Golub TR, Lander ES. Gene set enrichment analysis: a knowledge-based approach for interpreting genome-wide expression profiles. Proceedings of the National Academy of Sciences. 2005;102(43):15545–50.

76. Chen EY, Tan CM, Kou Y, Duan Q, Wang Z, Meirelles GV, Clark NR, Ma’ayan A. Enrichr: interactive and collaborative HTML5 gene list enrichment analysis tool. BMC bioinformatics. 2013;14(1):1–14.

77. Steele NG, Biffi G, Kemp SB, Zhang Y, Drouillard D, Syu L, Hao Y, Oni TE, Brosnan E, Elyada E. Inhibition of Hedgehog Signaling Alters Fibroblast Composition in Pancreatic CancerHedgehog Signaling in Pancreatic Cancer. Clinical Cancer Research. 2021;27(7):2023–37.

78. Zheng GX, Terry JM, Belgrader P, Ryvkin P, Bent ZW, Wilson R, Ziraldo SB, Wheeler TD, McDermott GP, Zhu J. Massively parallel digital transcriptional profiling of single cells. Nature communications. 2017;8(1):14049.

79. Hao Y, Hao S, Andersen-Nissen E, Mauck WM, Zheng S, Butler A, Lee MJ, Wilk AJ, Darby C, Zager M. Integrated analysis of multimodal single-cell data. Cell. 2021;184(13):3573–87. e29.

80. Stuart T, Butler A, Hoffman P, Hafemeister C, Papalexi E, Mauck WM, Hao Y, Stoeckius M, Smibert P, Satija R. Comprehensive integration of single-cell data. Cell. 2019;177(7):1888–902. e21.

81. Lee S-K, Boron WF, Parker MD. Monitoring ion activities in and around cells using ion-selective liquid-membrane microelectrodes. Sensors. 2013;13(1):984–1003.

82. Silva RC, Castilho BA, Sattlegger E. A Rapid Extraction Method for mammalian cell cultures, suitable for quantitative immunoblotting analysis of proteins, including phosphorylated GCN2 and eIF2a. MethodsX. 2018;5:75–82.

83. Francescone R, Vendramini-Costa DB, Franco-Barraza J, Wagner J, Muir A, Lau AN, Gabitova L, Pazina T, Gupta S, Luong T. Netrin G1 Promotes Pancreatic Tumorigenesis through Cancer-Associated Fibroblast–Driven Nutritional Support and Immunosuppression. Cancer discovery. 2021;11(2):446–79.

84. Horlbeck MA, Gilbert LA, Villalta JE, Adamson B, Pak RA, Chen Y, Fields AP, Park CY, Corn JE, Kampmann M. Compact and highly active next-generation libraries for CRISPR-mediated gene repression and activation. Elife. 2016;5:e19760.

