## Supplementary material for "Lorazepam stimulates IL-6 production and is associated with poor survival outcomes in pancreatic cancer": Tables and Supplemental Figures

#### Supplemental Figure S1.

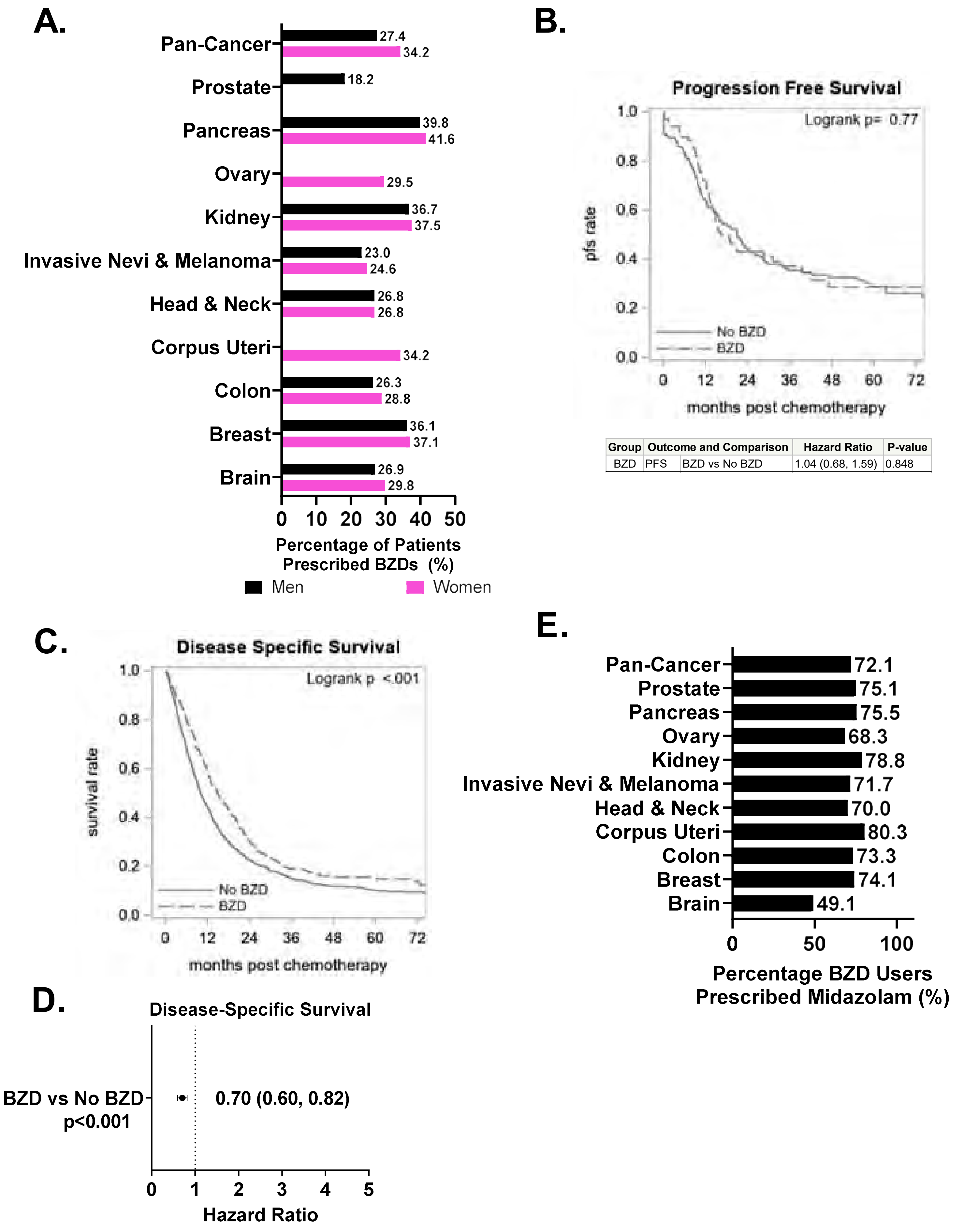


| Group | Outcome and Comparison | | Hazard Ratio | P-value |
|---|---|---|---|---|
| BZD | PFS | BZD vs No BZD | 1.04 (0.68, 1.59) | 0.848 |

#### Supplemental Figure S2.

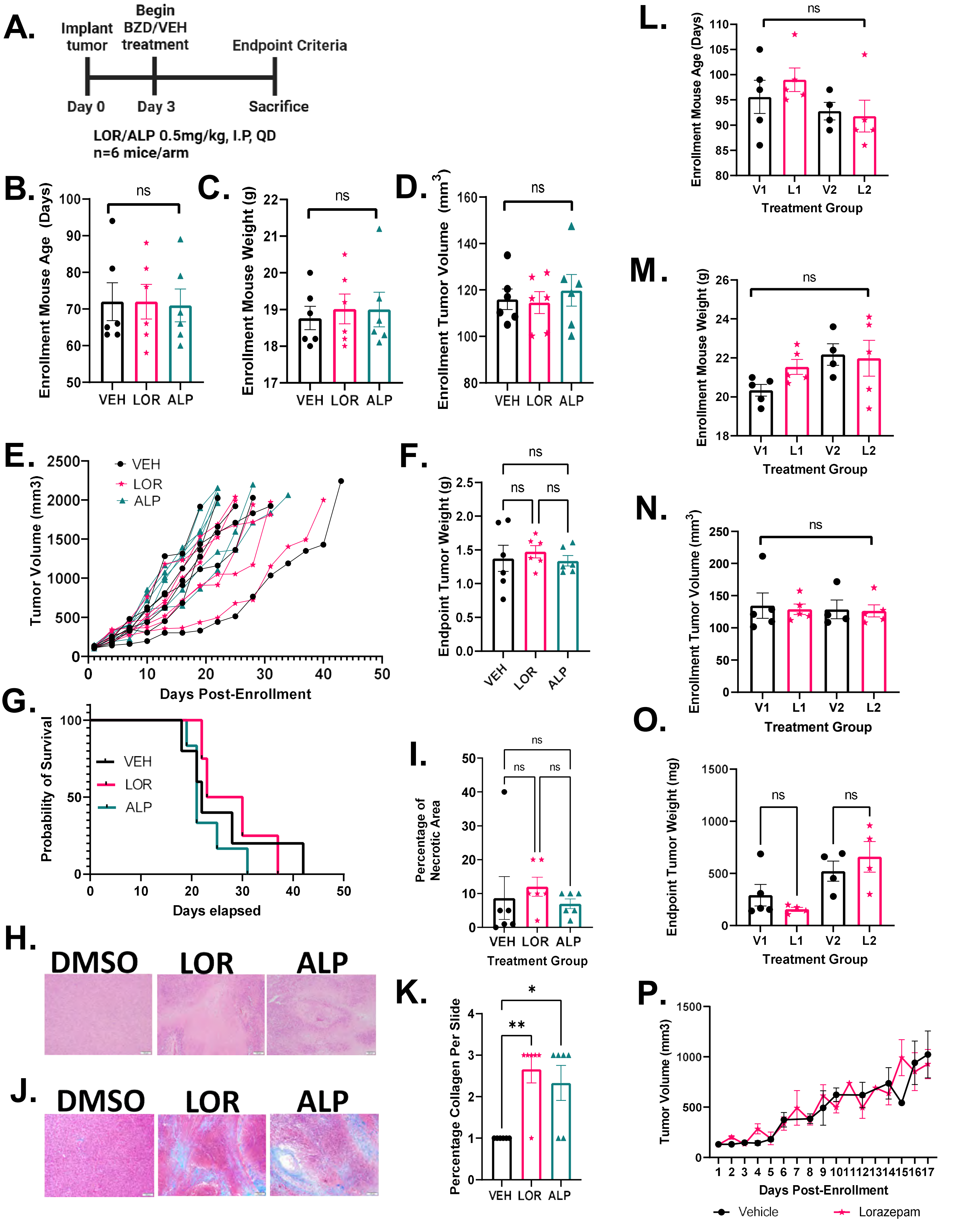

Supplemental Figure S2.

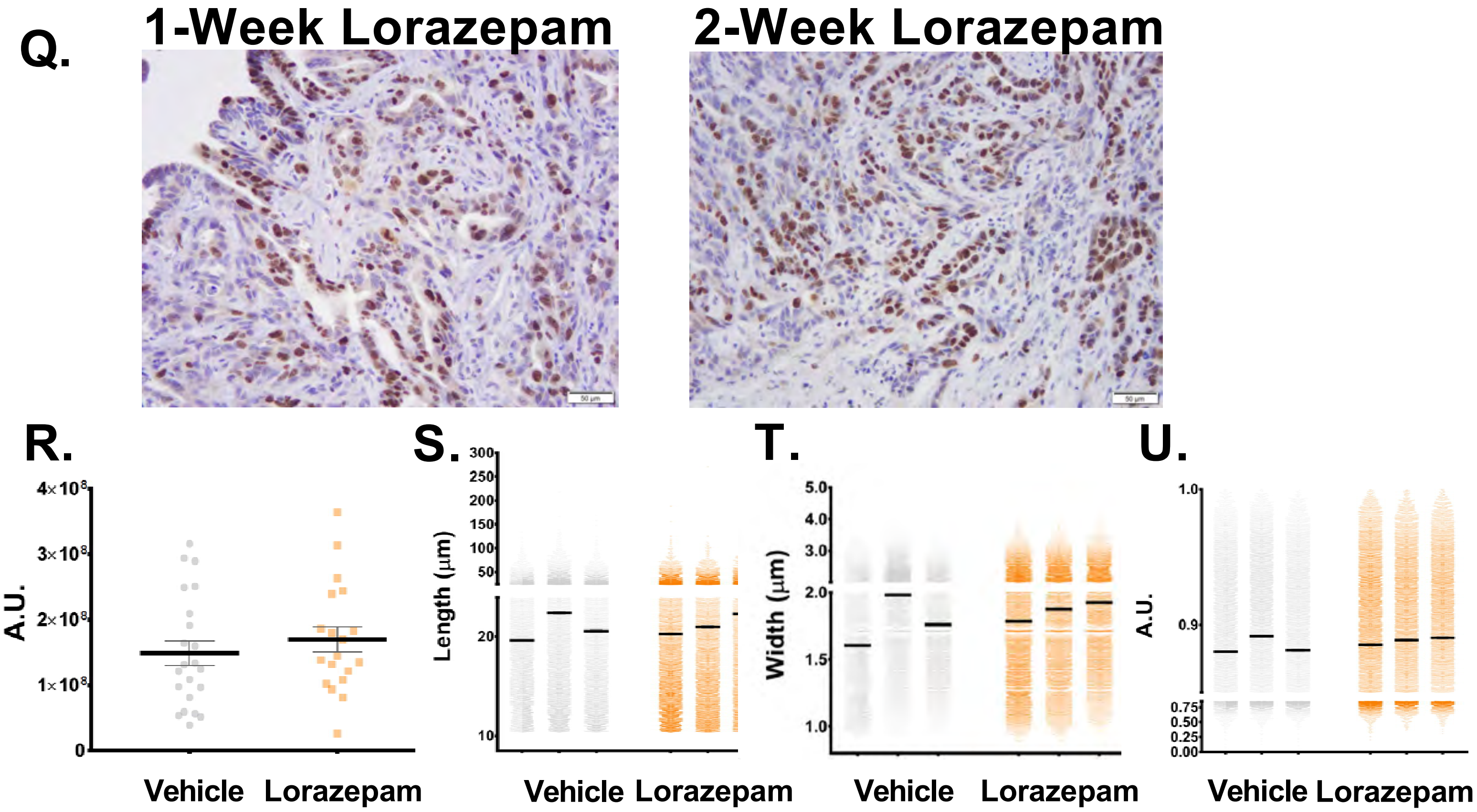

**Supplemental Figure S3.**

**A.**

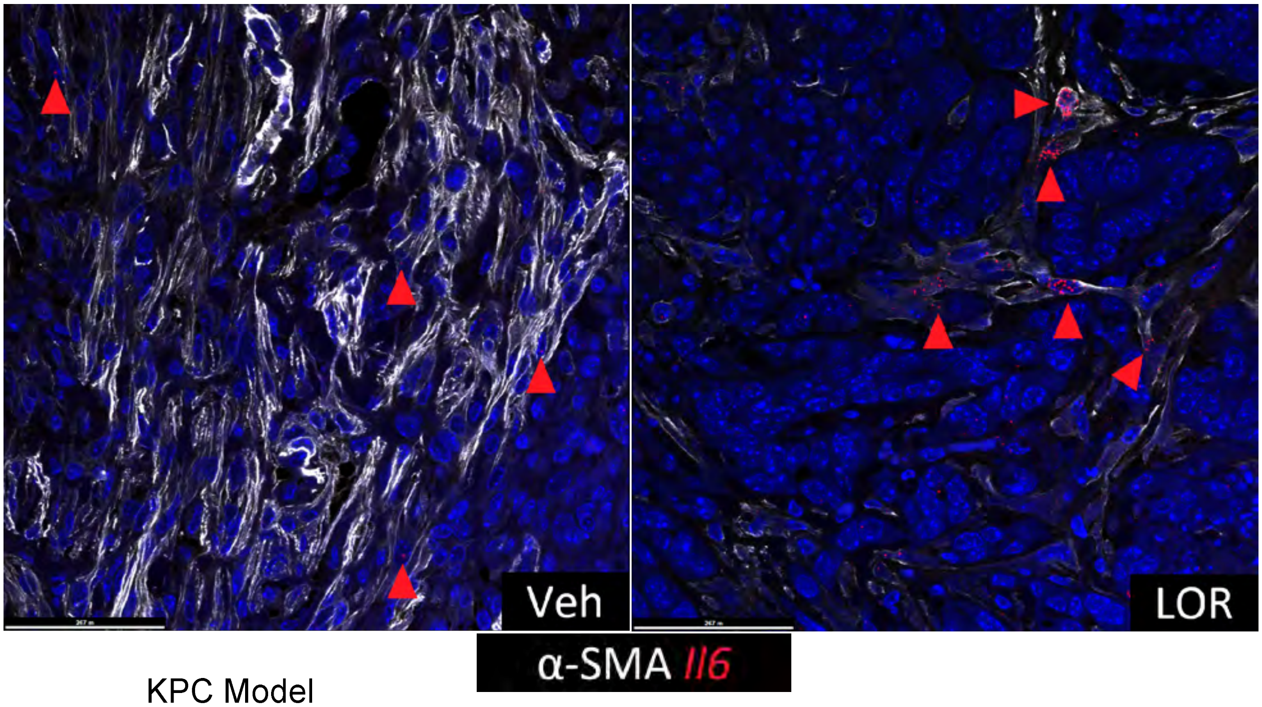


**B.**

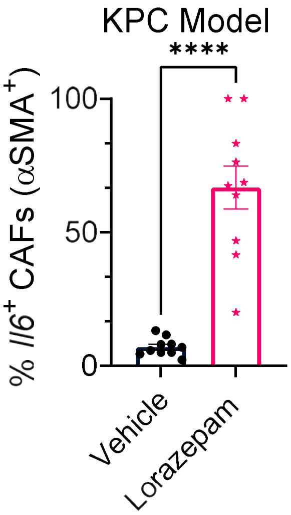

#### Supplemental Figure S4.

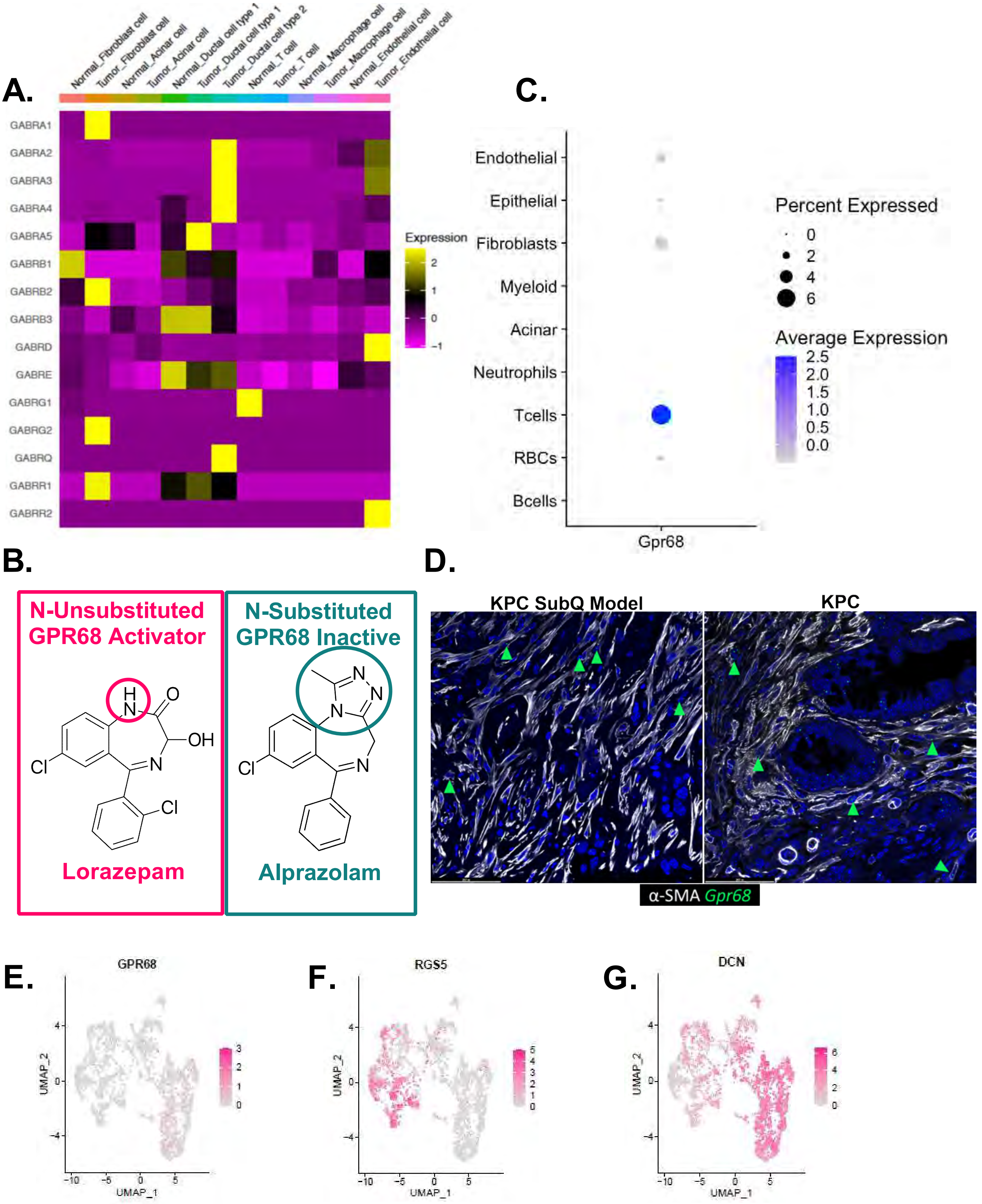

### Supplemental Figure S4.

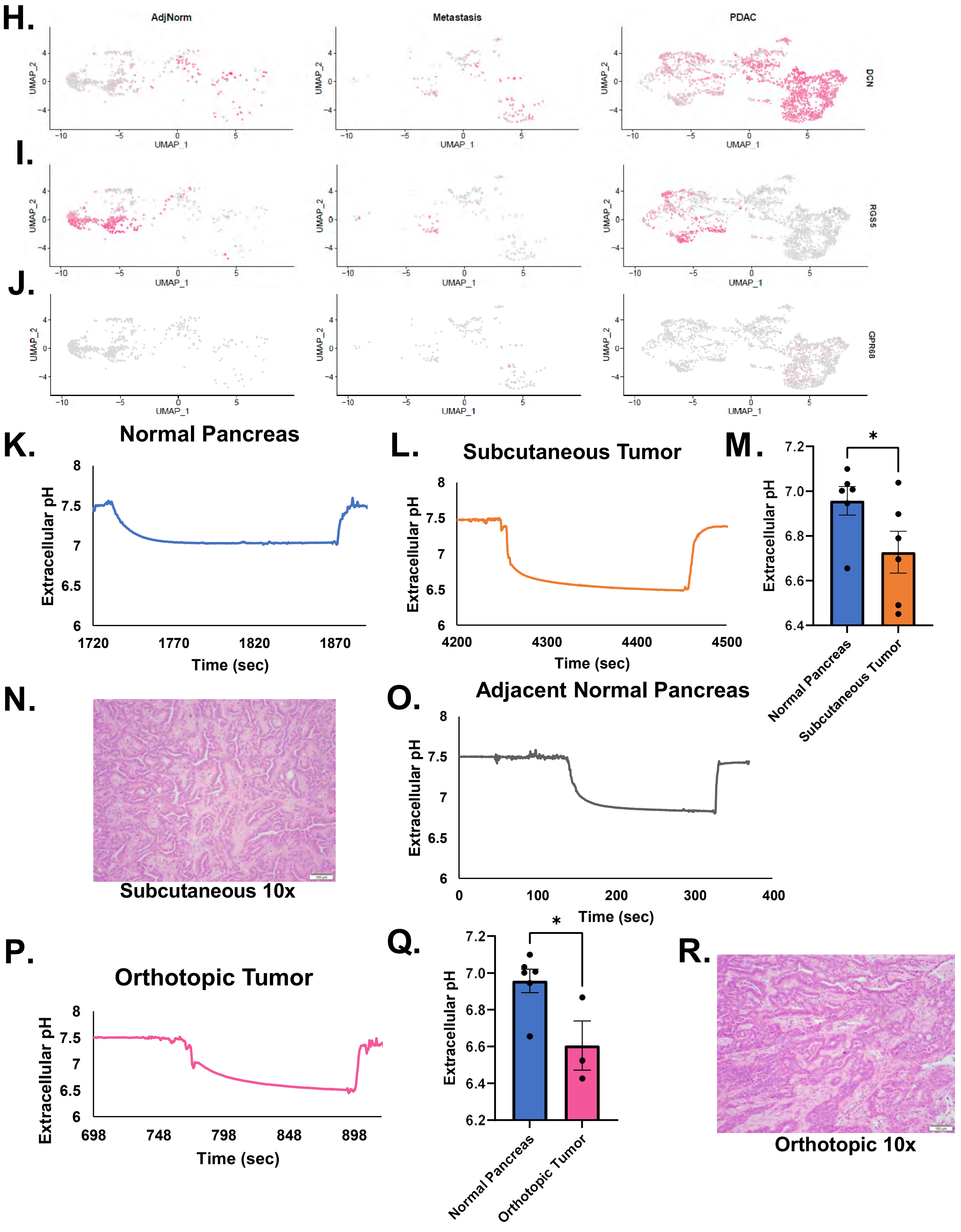

Supplemental Figure S5.

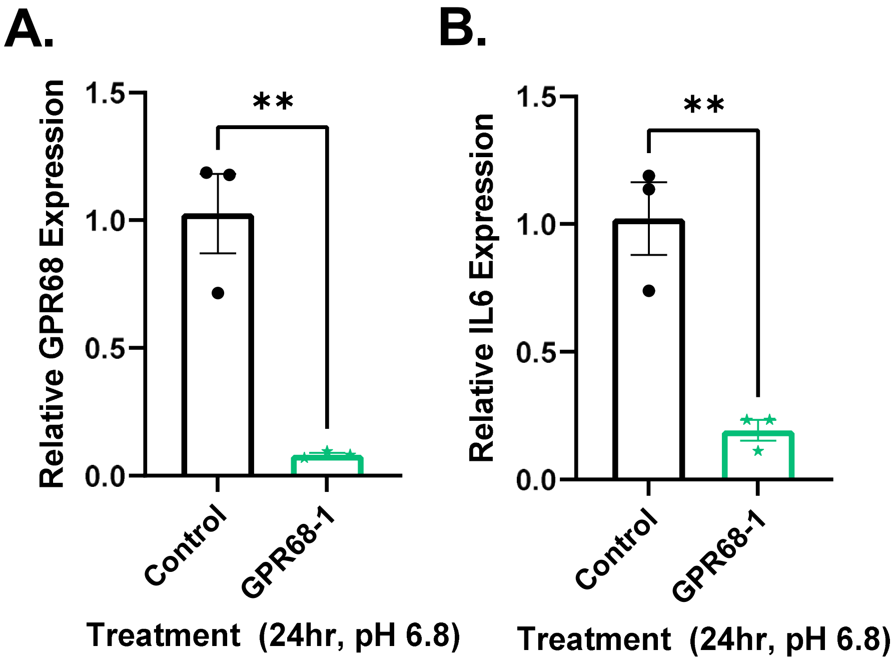

#### Supplemental Figure S6.

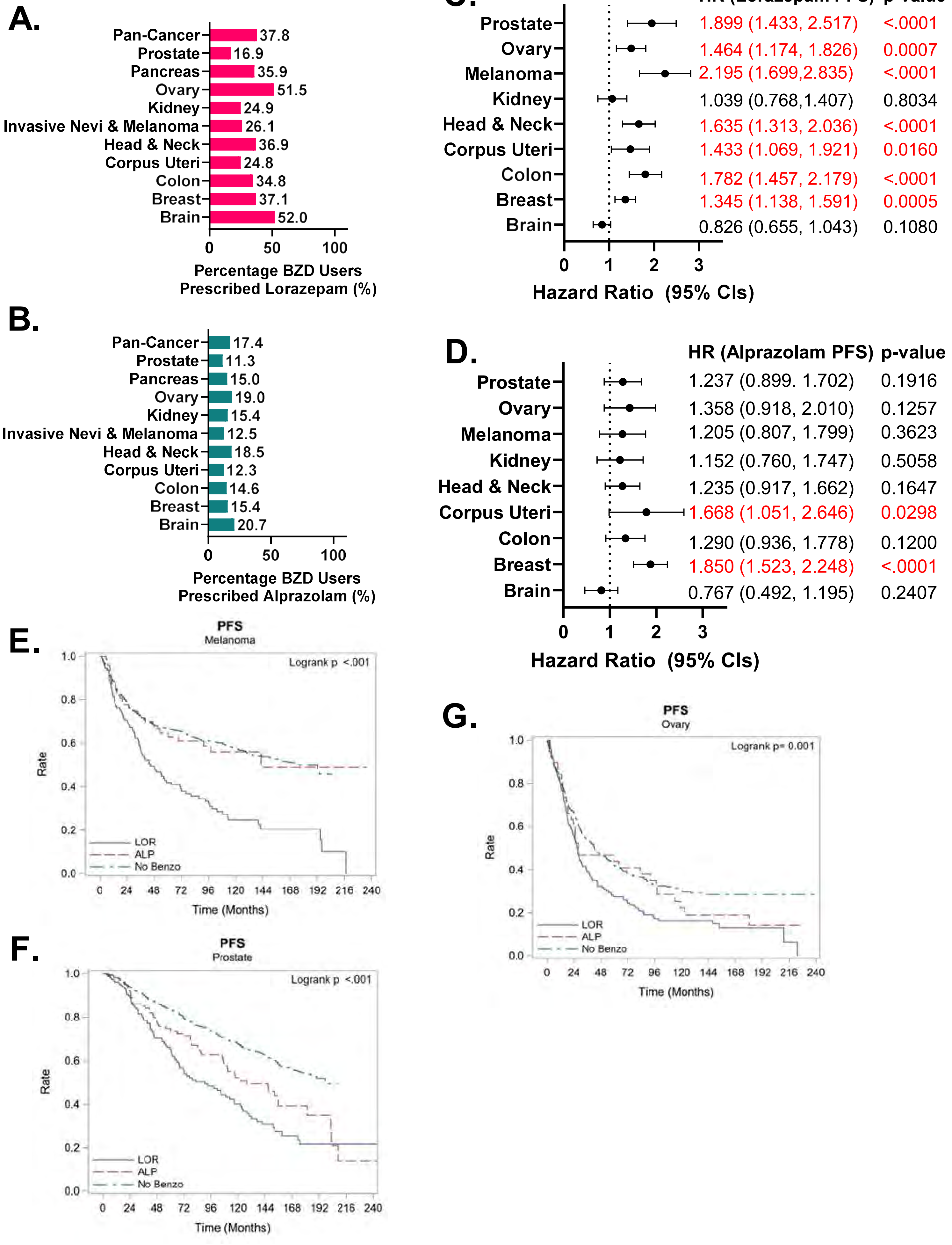

### Table 1. Pancreatic cancer patient characteristics by benzodiazepine prescription records

| | | No BZD | BZD | Overall | P-value |
|---|---|---|---|---|---|
| Overall | N | 1,093 (75.4) | 357 (24.6) | 1,450 (100%) | |
| Age @ Dx | Mean/Std/N | 66.2/10.9/1093 | 63.7/10.6/357 | 65.6/10.9/1450 | <.001 |
| | Median/Min/Max | 66.2/23.2/92.8 | 63.9/24.1/91.6 | 65.6/23.2/92.8 | |
| Age @ Chemo | Mean/Std/N | 66.3/10.9/1093 | 63.9/10.6/357 | 65.7/10.8/1450 | <.001 |
| | Median/Min/Max | 66.3/23.2/93.0 | 64.1/24.1/91.6 | 65.8/23.2/93.0 | |
| Sex | Male | 589 (53.9%) | 173 (48.5%) | 762 (52.6%) | 0.077 |
| | Female | 503 (46.1%) | 184 (51.5%) | 687 (47.4%) | |
| Race | White | 972 (89.9%) | 330 (94.3%) | 1,302 (91.0%) | 0.012 |
| | Black | 95 (8.8%) | 14 (4.0%) | 109 (7.6%) | |
| | Other | 14 (1.3%) | 6 (1.7%) | 20 (1.4%) | |
| Alcohol | Never | 353 (35.6%) | 111 (33.0%) | 464 (35.0%) | 0.679 |
| | Former | 94 (9.5%) | 32 (9.5%) | 126 (9.5%) | |
| | Current | 544 (54.9%) | 193 (57.4%) | 737 (55.5%) | |
| Tobacco | Never | 390 (35.8%) | 133 (37.8%) | 523 (36.3%) | 0.675 |
| | Former | 425 (39.0%) | 138 (39.2%) | 563 (39.1%) | |
| | Current | 274 (25.2%) | 81 (23.0%) | 355 (24.6%) | |
| Grade | I | 9 (2.0%) | 2 (2.1%) | 11 (2.0%) | 0.289 |
| | II | 163 (35.8%) | 39 (40.2%) | 202 (36.6%) | |
| | III | 211 (46.4%) | 48 (49.5%) | 259 (46.9%) | |
| | IV | 72 (15.8%) | 8 (8.2%) | 80 (14.5%) | |
| Clinical Stage | I | 146 (15.3%) | 59 (17.9%) | 205 (16.0%) | 0.604 |
| | II | 164 (17.2%) | 52 (15.8%) | 216 (16.8%) | |
| | III | 170 (17.8%) | 63 (19.1%) | 233 (18.1%) | |
| | IV | 475 (49.7%) | 156 (47.3%) | 631 (49.1%) | |
| Path Stage | 0 | | 2 (1.6%) | 2 (0.4%) | 0.009 |
| | I | 29 (8.7%) | 4 (3.1%) | 33 (7.2%) | |
| | II | 173 (52.1%) | 65 (50.8%) | 238 (51.7%) | |
| | III | 12 (3.6%) | 1 (0.8%) | 13 (2.8%) | |
| | IV | 118 (35.5%) | 56 (43.8%) | 174 (37.8%) | |
| Nodes Examined | Mean/Std/N | 4.6/9.4/1060 | 4.8/10.4/352 | 4.6/9.6/1412 | 0.988 |
| | Median/Min/Max | 0.0/0.0/47.0 | 0.0/0.0/71.0 | 0.0/0.0/71.0 | |
| Nodes Positive | Mean/Std/N | 2.2/3.0/266 | 2.2/2.7/87 | 2.2/3.0/353 | 0.419 |
| | Median/Min/Max | 1.0/0.0/18.0 | 1.0/0.0/12.0 | 1.0/0.0/18.0 | |
| Nodes | Negative | 105 (39.5%) | 27 (31.0%) | 132 (37.4%) | 0.163 |
| | Positive | 161 (60.5%) | 60 (69.0%) | 221 (62.6%) | |
| Persistent Disease | No | 219 (20.0%) | 69 (19.3%) | 288 (19.9%) | 0.819 |
| | Yes | 874 (80.0%) | 288 (80.7%) | 1,162 (80.1%) | |
| Tx: XRT | No | 807 (73.8%) | 290 (81.2%) | 1,097 (75.7%) | 0.004 |
| | Yes | 286 (26.2%) | 67 (18.8%) | 353 (24.3%) | |
| Tx: Immuno | No | 1,042 (95.3%) | 343 (96.1%) | 1,385 (95.5%) | 0.659 |
| | Yes | 51 (4.7%) | 14 (3.9%) | 65 (4.5%) | |
| Tx: Hormone | No | 1,044 (95.5%) | 348 (97.5%) | 1,392 (96.0%) | 0.119 |
| | Yes | 49 (4.5%) | 9 (2.5%) | 58 (4.0%) | |
| Tx: Surgery | No | 48 (4.4%) | 31 (8.7%) | 79 (5.4%) | 0.003 |

### Table 2. Pancreatic cancer patient characteristics by lorazepam prescription records

| | | No | Yes | P-value |
|---|---|---|---|---|
| Overall | N | 141 (39.5) | 216 (60.5) | |
| Age @ Dx | Mean/Std/N | 65.3/9.8/141 | 62.7/11.0/216 | 0.060 |
| | Median/Min/Max | 65.1/36.2/91.6 | 63.1/24.1/84.6 | |
| Age @ Chemo | Mean/Std/N | 65.4/9.7/141 | 62.9/11.0/216 | 0.061 |
| | Median/Min/Max | 65.1/39.3/91.6 | 63.2/24.1/84.7 | |
| Sex | Male | 76 (53.9%) | 97 (44.9%) | 0.105 |
| | Female | 65 (46.1%) | 119 (55.1%) | |
| Race | White | 125 (90.6%) | 205 (96.7%) | 0.037 |
| | Black | 10 (7.2%) | 4 (1.9%) | |
| | Other | 3 (2.2%) | 3 (1.4%) | |
| Alcohol | Never | 39 (29.8%) | 72 (35.1%) | 0.005 |
| | Former | 21 (16.0%) | 11 (5.4%) | |
| | Current | 71 (54.2%) | 122 (59.5%) | |
| Tobacco | Never | 49 (35.5%) | 84 (39.3%) | 0.652 |
| | Former | 54 (39.1%) | 84 (39.3%) | |
| | Current | 35 (25.4%) | 46 (21.5%) | |
| Grade | I | | 2 (3.6%) | 0.618 |
| | II | 18 (43.9%) | 21 (37.5%) | |
| | III | 20 (48.8%) | 28 (50.0%) | |
| | IV | 3 (7.3%) | 5 (8.9%) | |
| Clinical Stage | I | 24 (18.2%) | 35 (17.7%) | 0.999 |
| | II | 21 (15.9%) | 31 (15.7%) | |
| | III | 25 (18.9%) | 38 (19.2%) | |
| | IV | 62 (47.0%) | 94 (47.5%) | |
| Path Stage | 0 | 1 (1.9%) | 1 (1.3%) | 0.756 |
| | I | 2 (3.8%) | 2 (2.6%) | |
| | II | 29 (55.8%) | 36 (47.4%) | |
| | III | | 1 (1.3%) | |
| | IV | 20 (38.5%) | 36 (47.4%) | |
| Nodes Examined | Mean/Std/N | 5.5/11.5/140 | 4.4/9.6/212 | 0.210 |
| | Median/Min/Max | 0.0/0.0/71.0 | 0.0/0.0/47.0 | |
| Nodes Positive | Mean/Std/N | 1.9/2.4/40 | 2.6/2.8/47 | 0.208 |
| | Median/Min/Max | 1.0/0.0/10.0 | 2.0/0.0/12.0 | |
| Nodes | Negative | 14 (35.0%) | 13 (27.7%) | 0.493 |
| | Positive | 26 (65.0%) | 34 (72.3%) | |
| Persistent Disease | No | 29 (20.6%) | 40 (18.5%) | 0.681 |
| | Yes | 112 (79.4%) | 176 (81.5%) | |
| Tx: XRT | No | 117 (83.0%) | 173 (80.1%) | 0.580 |
| | Yes | 24 (17.0%) | 43 (19.9%) | |
| Tx: Immuno | No | 136 (96.5%) | 207 (95.8%) | 1.000 |
| | Yes | 5 (3.5%) | 9 (4.2%) | |
| Tx: Hormone | No | 137 (97.2%) | 211 (97.7%) | 0.744 |
| | Yes | 4 (2.8%) | 5 (2.3%) | |
| Tx: Surgery | No | 11 (7.8%) | 20 (9.3%) | 0.703 |
| | Yes | 130 (92.2%) | 196 (90.7%) | |

| | | | | |
|---|---|---|---|---|
| Timing of BZD | Prior to Start of Chemo | 79 (56.0%) | 78 (36.1%) | <.001 |
| | After Start of Chemo | 59 (41.8%) | 138 (63.9%) | |
| | At Start of Chemo | 3 (2.1%) | | |

### Table 3. Pancreatic cancer patient characteristics by alprazolam prescription records

| | | No | Yes | P-value |
|---|---|---|---|---|
| Overall | N | 225 (63.0) | 132 (37.0) | |
| Age @ Dx | Mean/Std/N | 63.4/10.6/225 | 64.3/10.8/132 | 0.619 |
| | Median/Min/Max | 63.2/24.1/83.5 | 64.1/35.5/91.6 | |
| Age @ Chemo | Mean/Std/N | 63.6/10.5/225 | 64.4/10.8/132 | 0.675 |
| | Median/Min/Max | 63.4/24.1/83.8 | 64.5/35.7/91.6 | |
| Sex | Male | 116 (51.6%) | 57 (43.2%) | 0.154 |
| | Female | 109 (48.4%) | 75 (56.8%) | |
| Race | White | 209 (95.0%) | 121 (93.1%) | 0.588 |
| | Black | 7 (3.2%) | 7 (5.4%) | |
| | Other | 4 (1.8%) | 2 (1.5%) | |
| Alcohol | Never | 70 (32.6%) | 41 (33.9%) | 0.940 |
| | Former | 20 (9.3%) | 12 (9.9%) | |
| | Current | 125 (58.1%) | 68 (56.2%) | |
| Tobacco | Never | 89 (40.1%) | 44 (33.8%) | 0.242 |
| | Former | 88 (39.6%) | 50 (38.5%) | |
| | Current | 45 (20.3%) | 36 (27.7%) | |
| Grade | I | 2 (3.2%) | | 0.354 |
| | II | 27 (43.5%) | 12 (34.3%) | |
| | III | 27 (43.5%) | 21 (60.0%) | |
| | IV | 6 (9.7%) | 2 (5.7%) | |
| Clinical Stage | I | 40 (19.3%) | 19 (15.4%) | 0.097 |
| | II | 33 (15.9%) | 19 (15.4%) | |
| | III | 31 (15.0%) | 32 (26.0%) | |
| | IV | 103 (49.8%) | 53 (43.1%) | |
| Path Stage | 0 | 1 (1.3%) | 1 (2.1%) | 0.908 |
| | I | 2 (2.5%) | 2 (4.2%) | |
| | II | 41 (51.3%) | 24 (50.0%) | |
| | III | 1 (1.3%) | | |
| | IV | 35 (43.8%) | 21 (43.8%) | |
| Nodes Examined | Mean/Std/N | 4.4/9.5/223 | 5.6/11.8/129 | 0.201 |
| | Median/Min/Max | 0.0/0.0/50.0 | 0.0/0.0/71.0 | |
| Nodes Positive | Mean/Std/N | 2.7/2.9/50 | 1.6/2.3/37 | 0.081 |
| | Median/Min/Max | 2.0/0.0/12.0 | 1.0/0.0/10.0 | |
| Nodes | Negative | 14 (28.0%) | 13 (35.1%) | 0.492 |
| | Positive | 36 (72.0%) | 24 (64.9%) | |
| Persistent Disease | No | 42 (18.7%) | 27 (20.5%) | 0.679 |
| | Yes | 183 (81.3%) | 105 (79.5%) | |
| Tx: XRT | No | 177 (78.7%) | 113 (85.6%) | 0.123 |
| | Yes | 48 (21.3%) | 19 (14.4%) | |
| Tx: Immuno | No | 213 (94.7%) | 130 (98.5%) | 0.092 |
| | Yes | 12 (5.3%) | 2 (1.5%) | |
| Tx: Hormone | No | 218 (96.9%) | 130 (98.5%) | 0.494 |
| | Yes | 7 (3.1%) | 2 (1.5%) | |
| Tx: Surgery | No | 19 (8.4%) | 12 (9.1%) | 0.847 |
| | Yes | 206 (91.6%) | 120 (90.9%) | |

| | | | | |
|---|---|---|---|---|
| Timing of BZD | Prior to Start of Chemo | 84 (37.3%) | 73 (55.3%) | <.001 |
| | After Start of Chemo | 141 (62.7%) | 56 (42.4%) | |
| | At Start of Chemo | | 3 (2.3%) | |

### Table 4. Pancreatic Cancer Time-to-Event Outcomes: Multivariate Summaries

| Group | Outcome and Comparison | | Hazard Ratio | P-value |
|---|---|---|---|---|
| BZD | DFS | BZD vs NoBZD | 0.98 (0.86, 1.12) | 0.771 |
| | DSS | BZD vs NoBZD | 0.70 (0.60, 0.82) | <.001 |
| | OS | BZD vs NoBZD | 0.73 (0.63, 0.84) | <.001 |
| | PFS | BZD vs NoBZD | 1.04 (0.68, 1.59) | 0.848 |
| BZD: LOR | DFS | Yes vs No | 1.09 (0.85, 1.39) | 0.497 |
| | DSS | Yes vs No | 1.07 (0.80, 1.42) | 0.667 |
| | OS | Yes vs No | 1.04 (0.79, 1.35) | 0.796 |
| | PFS | Yes vs No | 3.83 (1.53, 9.57) | 0.004 |
| BZD: ALP | DFS | Yes vs No | 0.88 (0.69, 1.12) | 0.305 |
| | DSS | Yes vs No | 0.99 (0.74, 1.31) | 0.921 |
| | OS | Yes vs No | 0.97 (0.75, 1.26) | 0.838 |
| | PFS | Yes vs No | 0.38 (0.16, 0.92) | 0.032 |

DFS: Disease-Free Survival, DSS: Disease-Specific Survival, OS:Overall Survival, PFS: Progression-Free Survival

### Table 5. Brain cancer patient characteristics, median survival, and Multivariate Cox Regression Modeling

| Patient Characteristics (Brain) | | LOR | ALP | No Benzo | Overall | P-value |
|---|---|---|---|---|---|---|
| | N | 153 (23.0) | 34 (5.1) | 477 (71.8) | 664 (100%) | |
| Sex | Female | 74 (48.4%) | 13 (38.2%) | 212 (44.4%) | 299 (45.0%) | 0.500 |
| | Male | 79 (51.6%) | 21 (61.8%) | 265 (55.6%) | 365 (55.0%) | |
| Grade (Clinical) | I | 1 (0.8%) | | 6 (1.3%) | 7 (1.1%) | 0.379 |
| | II | 7 (5.4%) | | 10 (2.1%) | 17 (2.7%) | |
| | III | 5 (3.9%) | 1 (3.3%) | 16 (3.4%) | 22 (3.5%) | |
| | IV | 32 (24.8%) | 8 (26.7%) | 93 (19.5%) | 133 (20.9%) | |
| | Not Reported | 84 (65.1%) | 21 (70.0%) | 351 (73.7%) | 456 (71.8%) | |
| Grade (Pathological) | I | 2 (1.5%) | | 5 (1.1%) | 7 (1.1%) | 0.003 |
| | II | 8 (6.0%) | | 10 (2.1%) | 18 (2.8%) | |
| | III | 4 (3.0%) | 2 (6.5%) | 16 (3.4%) | 22 (3.4%) | |
| | IV | 44 (33.1%) | 11 (35.5%) | 94 (19.7%) | 149 (23.3%) | |
| | Not Reported | 75 (56.4%) | 18 (58.1%) | 351 (73.7%) | 444 (69.4%) | |
| Stage (Clinical) | I | 16 (10.5%) | | | 16 (2.4%) | <.001 |
| | IV | | 1 (3.0%) | | 1 (0.2%) | |
| | Not Reported | 137 (89.5%) | 32 (97.0%) | 477 (100.0%) | 646 (97.4%) | |
| Stage (Pathological) | I | 1 (0.7%) | | | 1 (0.2%) | <.001 |
| | IV | | 1 (3.0%) | | 1 (0.2%) | |
| | Not Reported | 152 (99.3%) | 32 (97.0%) | 477 (100.0%) | 661 (99.7%) | |
| Overall Survival Indicator | Alive | 48 (31.4%) | 10 (29.4%) | 94 (19.7%) | 152 (22.9%) | 0.007 |
| | Dead | 105 (68.6%) | 24 (70.6%) | 383 (80.3%) | 512 (77.1%) | |
| Progression-Free Survival Indicator | No Progression | 45 (29.4%) | 9 (26.5%) | 89 (18.7%) | 143 (21.5%) | 0.015 |
| | Progression | 108 (70.6%) | 25 (73.5%) | 388 (81.3%) | 521 (78.5%) | |

| Overall Survival (OS) | Median Surv. (95% CI) | Sample: Brain | Log Rank P-value |
|---|---|---|---|
| Total | 16.6 (14.8, 18.3) | E=512 C=152 T=664 | p= 0.038 |
| ALP | 13.4 (7.9, 56.2) | E=24 C=10 T=34 | |
| LOR | 20.6 (15.6, 34.4) | E=105 C=48 T=153 | |
| No Benzo | 16.1 (14.2, 17.7) | E=383 C=94 T=477 | |

| Progression-Free Survival (PFS) | Median Surv. (95% CI) | Sample: Brain | Log Rank P-value |
|---|---|---|---|
| Total | 14.9 (13.6, 16.7) | E=521 C=143 T=664 | p= 0.117 |
| ALP | 13.4 (7.9, 24.9) | E=25 C=9 T=34 | |
| LOR | 16.7 (13.8, 28.5) | E=108 C=45 T=153 | |
| No Benzo | 14.5 (12.7, 16.4) | E=388 C=89 T=477 | |

| Overall Survival (OS) | | |
|---|---|---|
| Predictors: Cohort, Sex, Clinical Grade, and Clinical Stage | | |
| Cohort | HR (95% CI) | P-value |
| No Benzo | Ref. | 0.0357 |
| ALP | 0.661 (0.414 – 1.055) | 0.0829 |
| LOR | 0.779 (0.616 – 0.986) | 0.0381 |
| Progression-Free Survival (PFS) | | |
| No Benzo | Ref. | 0.1633 |
| ALP | 0.767 (0.492 – 1.195) | 0.2407 |
| LOR | 0.826 (0.655 – 1.043) | 0.1080 |

E=event, C=censored, T=Total

### Table 6. Breast cancer patient characteristics, median survival, and Multivariate Cox Regression Modeling

| Patient Characteristics (Breast) | | LOR | ALP | No Benzo | Overall | P-value |
|---|---|---|---|---|---|---|
| | N | 559 (24.0) | 362 (15.5) | 1,411 (60.5) | 2,332 (100%) | |
| Sex | Female | 555 (99.3%) | 360 (99.4%) | 1,404 (99.5%) | 2,319 (99.4%) | 0.840 |
| | Male | 4 (0.7%) | 2 (0.6%) | 7 (0.5%) | 13 (0.6%) | |
| Grade (Clinical) | I | 51 (9.3%) | 78 (21.9%) | 259 (18.7%) | 388 (16.9%) | <.001 |
| | II | 193 (35.1%) | 134 (37.6%) | 535 (38.5%) | 862 (37.6%) | |
| | III | 258 (46.9%) | 100 (28.1%) | 382 (27.5%) | 740 (32.3%) | |
| | IV | | | 2 (0.1%) | 2 (0.1%) | |
| | Not Reported | 48 (8.7%) | 44 (12.4%) | 210 (15.1%) | 302 (13.2%) | |
| Grade (Pathological) | I | 49 (8.9%) | 76 (21.3%) | 257 (18.5%) | 382 (16.6%) | <.001 |
| | II | 186 (33.8%) | 130 (36.5%) | 515 (37.1%) | 831 (36.2%) | |
| | III | 249 (45.3%) | 94 (26.4%) | 362 (26.1%) | 705 (30.7%) | |
| | IV | | | 2 (0.1%) | 2 (0.1%) | |
| | Not Reported | 66 (12.0%) | 56 (15.7%) | 253 (18.2%) | 375 (16.3%) | |
| Stage (Clinical) | 0 | 24 (4.3%) | 32 (8.9%) | 120 (8.5%) | 176 (7.6%) | <.001 |
| | I | 137 (24.5%) | 97 (27.0%) | 395 (28.1%) | 629 (27.1%) | |
| | II | 127 (22.7%) | 37 (10.3%) | 189 (13.4%) | 353 (15.2%) | |
| | III | 29 (5.2%) | 6 (1.7%) | 51 (3.6%) | 86 (3.7%) | |
| | IV | 47 (8.4%) | 8 (2.2%) | 75 (5.3%) | 130 (5.6%) | |
| | Not Reported | 195 (34.9%) | 179 (49.9%) | 576 (41.0%) | 950 (40.9%) | |
| Stage (Pathological) | 0 | 16 (2.9%) | 28 (7.8%) | 130 (9.4%) | 174 (7.6%) | <.001 |
| | I | 124 (22.4%) | 104 (29.1%) | 459 (33.1%) | 687 (29.9%) | |
| | II | 124 (22.4%) | 36 (10.1%) | 208 (15.0%) | 368 (16.0%) | |
| | III | 62 (11.2%) | 10 (2.8%) | 80 (5.8%) | 152 (6.6%) | |
| | IV | 11 (2.0%) | 1 (0.3%) | 25 (1.8%) | 37 (1.6%) | |
| | Not Reported | 216 (39.1%) | 179 (50.0%) | 483 (34.9%) | 878 (38.2%) | |
| Overall Survival Indicator | Alive | 349 (62.4%) | 227 (62.7%) | 1,021 (72.4%) | 1,597 (68.5%) | <.001 |
| | Dead | 210 (37.6%) | 135 (37.3%) | 390 (27.6%) | 735 (31.5%) | |
| Progression-Free Survival Indicator | No Progression | 332 (59.4%) | 220 (60.8%) | 1,000 (70.9%) | 1,552 (66.6%) | <.001 |
| | Progression | 227 (40.6%) | 142 (39.2%) | 411 (29.1%) | 780 (33.4%) | |

| Overall Survival (OS) | Median Surv. (95% CI) | Sample: Breast | Log Rank P-value |
|---|---|---|---|
| Total | 194.1 (183.5, NR) | E=735 C=1597 T=2332 | p= <.001 |
| ALP | 142.7 (114.0, 171.5) | E=135 C=227 T=362 | |
| LOR | 157.5 (133.2, 165.9) | E=210 C=349 T=559 | |
| No Benzo | NR (NR, NR) | E=390 C=1021 T=1411 | |

| Progression-Free Survival (PFS) | Median Surv. (95% CI) | Sample: Breast | Log Rank P-value |
|---|---|---|---|
| Total | 191.3 (172.5, NR) | E=780 C=1552 T=2332 | p= <.001 |
| ALP | 136.6 (92.7, 161.5) | E=142 C=220 T=362 | |
| LOR | 133.2 (117.1, 168.3) | E=227 C=332 T=559 | |
| No Benzo | NR (NR, NR) | E=411 C=1000 T=1411 | |

| Overall Survival (OS) | | |
|---|---|---|
| **Predictors: Cohort, Sex, Clinical Grade, and Clinical Stage** | | |
| **Cohort** | **HR (95% CI)** | **P-value** |
| No Benzo | Ref. | <.0001 |
| ALP | 1.867 (1.528 – 2.281) | <.0001 |
| LOR | 1.248 (1.050 – 1.484) | 0.0119 |
| **Progression-Free Survival (PFS)** | | |
| No Benzo | Ref. | <.0001 |
| ALP | 1.850 (1.523 – 2.248) | <.0001 |
| LOR | 1.345 (1.138 – 1.591) | 0.0005 |

E=event, C=censored, T=Total

### Table 7. Corpus uterine cancer patient characteristics, median survival, and Multivariate Cox Regression Modeling

| Patient Characteristics: Corpus Uteri | | LOR | ALP | No Benzo | Overall | P-value |
|---|---|---|---|---|---|---|
| | N | 120 (19.0) | 40 (6.3) | 470 (74.6) | 630 (100%) | |
| Sex | Female | 120 (100.0%) | 40 (100.0%) | 470 (100.0%) | 630 (100.0%) | |
| Grade (Clinical) | I | 21 (17.5%) | 9 (22.5%) | 213 (45.4%) | 243 (38.6%) | <.001 |
| | II | 21 (17.5%) | 8 (20.0%) | 104 (22.2%) | 133 (21.1%) | |
| | III | 37 (30.8%) | 10 (25.0%) | 97 (20.7%) | 144 (22.9%) | |
| | IV | 9 (7.5%) | 3 (7.5%) | 6 (1.3%) | 18 (2.9%) | |
| | Not Reported | 32 (26.7%) | 10 (25.0%) | 49 (10.4%) | 91 (14.5%) | |
| Grade (Pathological) | I | 23 (19.2%) | 9 (22.5%) | 213 (45.4%) | 245 (39.0%) | <.001 |
| | II | 21 (17.5%) | 6 (15.0%) | 104 (22.2%) | 131 (20.8%) | |
| | III | 38 (31.7%) | 12 (30.0%) | 96 (20.5%) | 146 (23.2%) | |
| | IV | 9 (7.5%) | 3 (7.5%) | 6 (1.3%) | 18 (2.9%) | |
| | Not Reported | 29 (24.2%) | 10 (25.0%) | 50 (10.7%) | 89 (14.1%) | |
| Stage (Clinical) | I | 29 (24.2%) | 12 (30.0%) | 160 (34.0%) | 201 (31.9%) | 0.043 |
| | II | 2 (1.7%) | | 9 (1.9%) | 11 (1.7%) | |
| | III | 2 (1.7%) | | 9 (1.9%) | 11 (1.7%) | |
| | IV | 6 (5.0%) | 2 (5.0%) | 4 (0.9%) | 12 (1.9%) | |
| | Not Reported | 81 (67.5%) | 26 (65.0%) | 288 (61.3%) | 395 (62.7%) | |
| Stage (Pathological) | I | 49 (41.2%) | 11 (28.2%) | 235 (50.0%) | 295 (47.0%) | <.001 |
| | II | 8 (6.7%) | 2 (5.1%) | 25 (5.3%) | 35 (5.6%) | |
| | III | 16 (13.4%) | 2 (5.1%) | 35 (7.4%) | 53 (8.4%) | |
| | IV | 14 (11.8%) | 5 (12.8%) | 15 (3.2%) | 34 (5.4%) | |
| | Not Reported | 32 (26.9%) | 19 (48.7%) | 160 (34.0%) | 211 (33.6%) | |
| Overall Survival Indicator | Alive | 48 (40.0%) | 19 (47.5%) | 296 (63.0%) | 363 (57.6%) | <.001 |
| | Dead | 72 (60.0%) | 21 (52.5%) | 174 (37.0%) | 267 (42.4%) | |
| Progression-Free Survival Indicator | No Progression | 46 (38.3%) | 18 (45.0%) | 286 (60.9%) | 350 (55.6%) | <.001 |
| | Progression | 74 (61.7%) | 22 (55.0%) | 184 (39.1%) | 280 (44.4%) | |

| Overall Survival (OS) | Median Surv. (95% CI) | Sample: Corpus Uteri | Log Rank P-value |
|---|---|---|---|
| Total | 150.5 (123.2, 181.2) | E=267 C=363 T=630 | p= <.001 |
| ALP | 52.9 (37.5, 123.2) | E=21 C=19 T=40 | |
| LOR | 62.9 (42.7, 90.3) | E=72 C=48 T=120 | |
| No Benzo | NR (157.8, NR) | E=174 C=296 T=470 | |

| Progression-Free Survival (PFS) | Median Surv. (95% CI) | Sample: Corpus Uteri | Log Rank P-value |
|---|---|---|---|
| Total | 138.3 (108.6, 162.0) | E=280 C=350 T=630 | p= <.001 |
| ALP | 38.1 (22.1, 113.6) | E=22 C=18 T=40 | |
| LOR | 40.6 (23.6, 76.3) | E=74 C=46 T=120 | |
| No Benzo | 178.3 (150.1, NR) | E=184 C=286 T=470 | |

| Overall Survival (OS) | | |
|---|---|---|
| **Predictors: Cohort, Clinical Grade, and Clinical Stage** | | |
| **Cohort** | **HR (95% CI)** | **P-value** |
| No Benzo | Ref. | 0.0564 |
| ALP | 1.497 (0.931 – 2.405) | 0.0956 |
| LOR | 1.376 (1.021 – 1.854) | 0.0362 |
| **Progression-Free Survival (PFS)** | | |
| No Benzo | Ref. | 0.0149 |
| ALP | 1.668 (1.051 – 2.646) | 0.0298 |
| LOR | 1.433 (1.069 – 1.921) | 0.0160 |

E=event, C=censored, T=Total

### Table 8. Head and neck cancer patient characteristics, median survival, and Multivariate Cox Regression Modeling

| Patient Characteristics (Head & Neck) | | LOR | ALP | No Benzo | Overall | P-value |
|---|---|---|---|---|---|---|
| | N | 171 (21.3) | 86 (10.7) | 546 (68.0) | 803 (100%) | |
| Sex | Female | 50 (29.2%) | 31 (36.0%) | 174 (31.9%) | 255 (31.8%) | 0.540 |
| | Male | 121 (70.8%) | 55 (64.0%) | 372 (68.1%) | 548 (68.2%) | |
| Grade (Clinical) | I | 13 (9.1%) | 8 (10.1%) | 60 (11.2%) | 81 (10.7%) | 0.559 |
| | II | 67 (46.9%) | 32 (40.5%) | 205 (38.3%) | 304 (40.2%) | |
| | III | 29 (20.3%) | 19 (24.1%) | 150 (28.0%) | 198 (26.2%) | |
| | IV | 3 (2.1%) | 1 (1.3%) | 5 (0.9%) | 9 (1.2%) | |
| | Not Reported | 31 (21.7%) | 19 (24.1%) | 115 (21.5%) | 165 (21.8%) | |
| Grade (Pathological) | I | 12 (8.5%) | 7 (8.9%) | 60 (11.2%) | 79 (10.4%) | 0.493 |
| | II | 61 (43.0%) | 33 (41.8%) | 204 (38.1%) | 298 (39.4%) | |
| | III | 29 (20.4%) | 18 (22.8%) | 151 (28.2%) | 198 (26.2%) | |
| | IV | 3 (2.1%) | 1 (1.3%) | 5 (0.9%) | 9 (1.2%) | |
| | Not Reported | 37 (26.1%) | 20 (25.3%) | 115 (21.5%) | 172 (22.8%) | |
| Stage (Clinical) | 0 | 3 (1.8%) | 1 (1.2%) | 8 (1.5%) | 12 (1.5%) | 0.027 |
| | I | 27 (15.8%) | 13 (15.1%) | 64 (11.7%) | 104 (13.0%) | |
| | II | 27 (15.8%) | 4 (4.7%) | 63 (11.5%) | 94 (11.7%) | |
| | III | 26 (15.2%) | 11 (12.8%) | 53 (9.7%) | 90 (11.2%) | |
| | IV | 48 (28.1%) | 23 (26.7%) | 192 (35.2%) | 263 (32.8%) | |
| | Not Reported | 40 (23.4%) | 34 (39.5%) | 166 (30.4%) | 240 (29.9%) | |
| Stage (Pathological) | 0 | 1 (0.6%) | 1 (1.2%) | 7 (1.3%) | 9 (1.1%) | 0.509 |
| | I | 7 (4.1%) | 5 (5.9%) | 49 (9.0%) | 61 (7.6%) | |
| | II | 5 (2.9%) | 1 (1.2%) | 17 (3.1%) | 23 (2.9%) | |
| | III | 7 (4.1%) | 2 (2.4%) | 20 (3.7%) | 29 (3.6%) | |
| | IV | 21 (12.4%) | 10 (11.8%) | 82 (15.0%) | 113 (14.1%) | |
| | Not Reported | 129 (75.9%) | 66 (77.6%) | 371 (67.9%) | 566 (70.7%) | |
| Overall Survival Indicator | Alive | 53 (31.0%) | 33 (38.4%) | 212 (38.8%) | 298 (37.1%) | 0.175 |
| | Dead | 118 (69.0%) | 53 (61.6%) | 334 (61.2%) | 505 (62.9%) | |
| Progression-Free Survival Indicator | No Progression | 49 (28.7%) | 32 (37.2%) | 199 (36.4%) | 280 (34.9%) | 0.156 |
| | Progression | 122 (71.3%) | 54 (62.8%) | 347 (63.6%) | 523 (65.1%) | |

| Overall Survival (OS) | Median Surv. (95% CI) | Sample: Head & Neck | Log Rank P-value |
|---|---|---|---|
| Total | 55.2 (44.2, 68.0) | E=505 C=298 T=803 | p= 0.012 |
| ALP | 51.3 (30.7, 90.6) | E=53 C=33 T=86 | |
| LOR | 35.0 (23.8, 56.5) | E=118 C=53 T=171 | |
| No Benzo | 63.7 (48.2, 79.1) | E=334 C=212 T=546 | |

| Progression-Free Survival (PFS) | Median Surv. (95% CI) | Sample: Head & Neck | Log Rank P-value |
|---|---|---|---|
| Total | 38.7 (30.6, 52.1) | E=523 C=280 T=803 | p= 0.006 |
| ALP | 40.3 (23.7, 75.2) | E=54 C=32 T=86 | |
| LOR | 29.2 (18.0, 44.0) | E=122 C=49 T=171 | |
| No Benzo | 46.8 (32.0, 61.3) | E=347 C=199 T=546 | |

| Overall Survival (OS) | | |
|---|---|---|
| **Predictors: Cohort, Sex, Clinical Grade, and Clinical Stage** | | |
| **Cohort** | **HR (95% CI)** | **P-value** |
| No Benzo | Ref. | <.0001 |
| ALP | 1.227 (0.907 – 1.660) | 0.1845 |
| LOR | 1.629 (1.304 – 2.035) | <.0001 |
| **Progression-Free Survival (PFS)** | | |
| No Benzo | Ref. | <.0001 |
| ALP | 1.235 (0.917 – 1.662) | 0.1647 |
| LOR | 1.635 (1.313 – 2.036) | <.0001 |

E=event, C=censored, T=Total

### Table 9. Invasive nevi and melanoma patient characteristics, median survival, and Multivariate Cox Regression Modeling

| Patient Characteristics: Invasive Nevi & Melanoma | | LOR | ALP | No Benzo | Overall | P-value |
|---|---|---|---|---|---|---|
| | N | 130 (18.6) | 76 (10.9) | 494 (70.6) | 700 (100%) | |
| Sex | Female | 54 (41.5%) | 43 (56.6%) | 231 (46.8%) | 328 (46.9%) | 0.113 |
| | Male | 76 (58.5%) | 33 (43.4%) | 263 (53.2%) | 372 (53.1%) | |
| Grade (Clinical) | I | | | 1 (0.2%) | 1 (0.1%) | 0.707 |
| | III | | 1 (1.3%) | 2 (0.4%) | 3 (0.4%) | |
| | IV | 1 (0.8%) | | 1 (0.2%) | 2 (0.3%) | |
| | Not Reported | 129 (99.2%) | 75 (98.7%) | 490 (99.2%) | 694 (99.1%) | |
| Grade (Pathological) | I | | | 1 (0.2%) | 1 (0.1%) | 0.527 |
| | III | | 1 (1.3%) | 1 (0.2%) | 2 (0.3%) | |
| | IV | 1 (0.8%) | | 1 (0.2%) | 2 (0.3%) | |
| | Not Reported | 129 (99.2%) | 75 (98.7%) | 491 (99.4%) | 695 (99.3%) | |
| Stage (Clinical) | 0 | | | 1 (0.2%) | 1 (0.1%) | <.001 |
| | I | 32 (24.6%) | 34 (44.7%) | 133 (27.0%) | 199 (28.5%) | |
| | II | 20 (15.4%) | 14 (18.4%) | 47 (9.5%) | 81 (11.6%) | |
| | III | 6 (4.6%) | 4 (5.3%) | 9 (1.8%) | 19 (2.7%) | |
| | IV | 4 (3.1%) | 1 (1.3%) | 13 (2.6%) | 18 (2.6%) | |
| | Not Reported | 68 (52.3%) | 23 (30.3%) | 290 (58.8%) | 381 (54.5%) | |
| Stage (Pathological) | 0 | | 1 (1.3%) | 3 (0.6%) | 4 (0.6%) | 0.002 |
| | I | 36 (27.7%) | 28 (36.8%) | 219 (44.4%) | 283 (40.5%) | |
| | II | 16 (12.3%) | 15 (19.7%) | 70 (14.2%) | 101 (14.4%) | |
| | III | 18 (13.8%) | 13 (17.1%) | 44 (8.9%) | 75 (10.7%) | |
| | IV | 1 (0.8%) | | 9 (1.8%) | 10 (1.4%) | |
| | Not Reported | 59 (45.4%) | 19 (25.0%) | 148 (30.0%) | 226 (32.3%) | |
| Overall Survival Indicator | Alive | 49 (37.7%) | 49 (64.5%) | 297 (60.1%) | 395 (56.4%) | <.001 |
| | Dead | 81 (62.3%) | 27 (35.5%) | 197 (39.9%) | 305 (43.6%) | |
| Progression-Free Survival Indicator | No Progression | 43 (33.1%) | 47 (61.8%) | 287 (58.1%) | 377 (53.9%) | <.001 |
| | Progression | 87 (66.9%) | 29 (38.2%) | 207 (41.9%) | 323 (46.1%) | |

| Overall Survival (OS) | Median Surv. (95% CI) | Sample: Invasive Nevi & Melanoma | Log Rank P-value |
|---|---|---|---|
| Total | 144.7 (120.0, 193.7) | E=305 C=395 T=700 | p= <.001 |
| ALP | NR (69.4, NR) | E=27 C=49 T=76 | |
| LOR | 72.2 (54.1, 97.4) | E=81 C=49 T=130 | |
| No Benzo | 193.7 (150.3, NR) | E=197 C=297 T=494 | |

| Progression-Free Survival (PFS) | Median Surv. (95% CI) | Sample: Invasive Nevi & Melanoma | Log Rank P-value |
|---|---|---|---|
| Total | 132.7 (103.7, 165.9) | E=323 C=377 T=700 | p= <.001 |
| ALP | 142.3 (69.4, NR) | E=29 C=47 T=76 | |
| LOR | 44.7 (34.4, 63.5) | E=87 C=43 T=130 | |
| No Benzo | 192.2 (134.3, NR) | E=207 C=287 T=494 | |

| Overall Survival (OS) | | |
|---|---|---|
| **Predictors: Cohort, Sex, Clinical Grade, and Clinical Stage** | | |
| **Cohort** | **HR (95% CI)** | **P-value** |
| No Benzo | Ref. | <.0001 |
| ALP | 1.194 (0.791 – 1.803) | 0.3984 |
| LOR | 1.978 (1.519 – 2.576) | <.0001 |
| **Progression-Free Survival (PFS)** | | |
| No Benzo | Ref. | <.0001 |
| ALP | 1.205 (0.807 – 1.799) | 0.3623 |
| LOR | 2.195 (1.699 – 2.835) | <.0001 |

E=event, C=censored, T=Total

### Table 10. Renal cancer patient characteristics, median survival, and Multivariate Cox Regression Modeling

| Patient Characteristics: Kidney | | LOR | ALP | No Benzo | Overall | P-value |
|---|---|---|---|---|---|---|
| | N | 73 (12.4) | 38 (6.5) | 477 (81.1) | 588 (100%) | |
| Sex | Female | 25 (34.2%) | 21 (55.3%) | 185 (38.8%) | 231 (39.3%) | 0.087 |
| | Male | 48 (65.8%) | 17 (44.7%) | 292 (61.2%) | 357 (60.7%) | |
| Grade (Clinical) | I | | 3 (7.9%) | 23 (4.9%) | 26 (4.5%) | 0.288 |
| | II | 18 (24.7%) | 11 (28.9%) | 131 (28.0%) | 160 (27.6%) | |
| | III | 24 (32.9%) | 13 (34.2%) | 135 (28.8%) | 172 (29.7%) | |
| | IV | 6 (8.2%) | 2 (5.3%) | 59 (12.6%) | 67 (11.6%) | |
| | Not Reported | 25 (34.2%) | 9 (23.7%) | 120 (25.6%) | 154 (26.6%) | |
| Grade (Pathological) | I | | 3 (7.9%) | 23 (4.9%) | 26 (4.5%) | 0.435 |
| | II | 18 (24.7%) | 10 (26.3%) | 129 (27.5%) | 157 (27.1%) | |
| | III | 23 (31.5%) | 13 (34.2%) | 136 (29.0%) | 172 (29.7%) | |
| | IV | 8 (11.0%) | 2 (5.3%) | 60 (12.8%) | 70 (12.1%) | |
| | Not Reported | 24 (32.9%) | 10 (26.3%) | 121 (25.8%) | 155 (26.7%) | |
| Stage (Clinical) | I | 13 (17.8%) | 9 (24.3%) | 133 (27.9%) | 155 (26.4%) | 0.412 |
| | II | 4 (5.5%) | 1 (2.7%) | 44 (9.2%) | 49 (8.3%) | |
| | III | 4 (5.5%) | 1 (2.7%) | 24 (5.0%) | 29 (4.9%) | |
| | IV | 16 (21.9%) | 7 (18.9%) | 78 (16.4%) | 101 (17.2%) | |
| | Not Reported | 36 (49.3%) | 19 (51.4%) | 198 (41.5%) | 253 (43.1%) | |
| Stage (Pathological) | I | 10 (13.7%) | 7 (18.9%) | 106 (22.2%) | 123 (21.0%) | 0.482 |
| | II | 3 (4.1%) | 2 (5.4%) | 32 (6.7%) | 37 (6.3%) | |
| | III | 5 (6.8%) | 2 (5.4%) | 47 (9.9%) | 54 (9.2%) | |
| | IV | 9 (12.3%) | 5 (13.5%) | 39 (8.2%) | 53 (9.0%) | |
| | Not Reported | 46 (63.0%) | 21 (56.8%) | 253 (53.0%) | 320 (54.5%) | |
| Overall Survival Indicator | Alive | 26 (35.6%) | 16 (42.1%) | 229 (48.0%) | 271 (46.1%) | 0.124 |
| | Dead | 47 (64.4%) | 22 (57.9%) | 248 (52.0%) | 317 (53.9%) | |
| Progression-Free Survival Indicator | No Progression | 20 (27.4%) | 13 (34.2%) | 205 (43.0%) | 238 (40.5%) | 0.030 |
| | Progression | 53 (72.6%) | 25 (65.8%) | 272 (57.0%) | 350 (59.5%) | |

| Overall Survival (OS) | Median Surv. (95% CI) | Sample: Kidney | Log Rank P-value |
|---|---|---|---|
| Total | 82.4 (65.6, 96.0) | E=317 C=271 T=588 | p= 0.272 |
| ALP | 66.9 (24.0, 120.6) | E=22 C=16 T=38 | |
| LOR | 66.9 (48.3, 96.0) | E=47 C=26 T=73 | |
| No Benzo | 86.8 (66.2, 105.9) | E=248 C=229 T=477 | |

| Progression-Free Survival (PFS) | Median Surv. (95% CI) | Sample: Kidney | Log Rank P-value |
|---|---|---|---|
| Total | 49.1 (38.3, 66.8) | E=350 C=238 T=588 | p= 0.051 |
| ALP | 35.6 (13.9, 95.7) | E=25 C=13 T=38 | |
| LOR | 36.8 (26.1, 53.8) | E=53 C=20 T=73 | |
| No Benzo | 59.8 (39.1, 79.2) | E=272 C=205 T=477 | |

| Overall Survival (OS) | | |
|---|---|---|
| **Predictors: Cohort, Sex, Clinical Grade, and Clinical Stage** | | |
| **Cohort** | **HR (95% CI)** | **P-value** |
| No Benzo | Ref. | 0.6198 |
| ALP | 1.153 (0.741 – 1.795) | 0.5268 |
| LOR | 0.898 (0.651 – 1.240) | 0.5132 |
| **Progression-Free Survival (PFS)** | | |
| No Benzo | Ref. | 0.7907 |
| ALP | 1.152 (0.760 – 1.747) | 0.5058 |
| LOR | 1.039 (0.768 – 1.407) | 0.8034 |

E=event, C=censored, T=Total

### Table 11. Ovarian cancer patient characteristics, median survival, and Multivariate Cox Regression Modeling

| Patient Characteristics: Ovary | | LOR | ALP | No Benzo | Overall | P-value |
|---|---|---|---|---|---|---|
| | N | 179 (31.4) | 41 (7.2) | 350 (61.4) | 570 (100%) | |
| Sex | Female | 179 (100.0%) | 41 (100.0%) | 350 (100.0%) | 570 (100.0%) | |
| Grade (Clinical) | I | 8 (4.5%) | 2 (4.9%) | 27 (7.8%) | 37 (6.6%) | 0.010 |
| | II | 25 (14.1%) | 3 (7.3%) | 43 (12.5%) | 71 (12.6%) | |
| | III | 77 (43.5%) | 12 (29.3%) | 103 (29.9%) | 192 (34.2%) | |
| | IV | 15 (8.5%) | 8 (19.5%) | 28 (8.1%) | 51 (9.1%) | |
| | Not Reported | 52 (29.4%) | 16 (39.0%) | 143 (41.6%) | 211 (37.5%) | |
| Grade (Pathological) | I | 8 (4.5%) | 2 (4.9%) | 29 (8.5%) | 39 (7.0%) | 0.021 |
| | II | 25 (14.0%) | 3 (7.3%) | 44 (12.9%) | 72 (12.9%) | |
| | III | 77 (43.3%) | 12 (29.3%) | 105 (30.9%) | 194 (34.7%) | |
| | IV | 15 (8.4%) | 8 (19.5%) | 28 (8.2%) | 51 (9.1%) | |
| | Not Reported | 53 (29.8%) | 16 (39.0%) | 134 (39.4%) | 203 (36.3%) | |
| Stage (Clinical) | I | 12 (6.8%) | 5 (12.2%) | 30 (8.6%) | 47 (8.3%) | 0.339 |
| | II | 4 (2.3%) | 2 (4.9%) | 11 (3.1%) | 17 (3.0%) | |
| | III | 16 (9.1%) | 4 (9.8%) | 29 (8.3%) | 49 (8.6%) | |
| | IV | 9 (5.1%) | | 33 (9.4%) | 42 (7.4%) | |
| | Not Reported | 135 (76.7%) | 30 (73.2%) | 247 (70.6%) | 412 (72.7%) | |
| Stage (Pathological) | I | 20 (11.4%) | 6 (14.6%) | 70 (20.2%) | 96 (17.0%) | <.001 |
| | II | 5 (2.8%) | 3 (7.3%) | 26 (7.5%) | 34 (6.0%) | |
| | III | 70 (39.8%) | 11 (26.8%) | 75 (21.6%) | 156 (27.7%) | |
| | IV | 22 (12.5%) | 1 (2.4%) | 39 (11.2%) | 62 (11.0%) | |
| | Not Reported | 59 (33.5%) | 20 (48.8%) | 137 (39.5%) | 216 (38.3%) | |
| Overall Survival Indicator | Alive | 47 (26.3%) | 12 (29.3%) | 161 (46.0%) | 220 (38.6%) | <.001 |
| | Dead | 132 (73.7%) | 29 (70.7%) | 189 (54.0%) | 350 (61.4%) | |
| Progression-Free Survival Indicator | No Progression | 43 (24.0%) | 11 (26.8%) | 145 (41.4%) | 199 (34.9%) | <.001 |
| | Progression | 136 (76.0%) | 30 (73.2%) | 205 (58.6%) | 371 (65.1%) | |

| Overall Survival (OS) | Median Surv. (95% CI) | Sample: Ovary | Log Rank P-value |
|---|---|---|---|
| Total | 56.3 (47.3, 66.8) | E=350 C=220 T=570 | p= <.001 |
| ALP | 64.2 (27.8, 97.2) | E=29 C=12 T=41 | |
| LOR | 40.5 (32.9, 47.9) | E=132 C=47 T=179 | |
| No Benzo | 70.5 (54.7, 88.8) | E=189 C=161 T=350 | |

| Progression-Free Survival (PFS) | Median Surv. (95% CI) | Sample: Ovary | Log Rank P-value |
|---|---|---|---|
| Total | 33.7 (28.5, 41.8) | E=370 C=199 T=569 | p= 0.001 |
| ALP | 27.8 (18.5, 97.2) | E=30 C=11 T=41 | |
| LOR | 26.4 (22.6, 31.1) | E=135 C=43 T=178 | |
| No Benzo | 42.2 (33.3, 54.7) | E=205 C=145 T=350 | |

| Overall Survival (OS) | | |
|---|---|---|
| **Predictors: Cohort, Clinical Grade, and Clinical Stage** | | |
| **Cohort** | **HR (95% CI)** | **P-value** |
| No Benzo | Ref. | 0.0011 |
| ALP | 1.343 (0.901 – 2.002) | 0.1481 |
| LOR | 1.521 (1.212 – 1.907) | 0.0003 |
| **Progression-Free Survival (PFS)** | | |
| No Benzo | Ref. | 0.0025 |
| ALP | 1.358 (0.918 – 2.010) | 0.1257 |
| LOR | 1.464 (1.174 – 1.826) | 0.0007 |

E=event, C=censored, T=Total

### Table 12. Colon cancer patient characteristics, median survival, and Multivariate Cox Regression Modeling

| Patient Characteristics: Colon | | LOR | ALP | No Benzo | Overall | P-value |
|---|---|---|---|---|---|---|
| | N | 177 (22.7) | 64 (8.2) | 539 (69.1) | 780 (100%) | |
| Sex | Female | 96 (54.2%) | 43 (67.2%) | 267 (49.5%) | 406 (52.1%) | 0.023 |
| | Male | 81 (45.8%) | 21 (32.8%) | 272 (50.5%) | 374 (47.9%) | |
| Grade (Clinical) | I | 8 (4.7%) | 3 (4.7%) | 23 (4.3%) | 34 (4.4%) | 0.948 |
| | II | 98 (57.6%) | 42 (65.6%) | 332 (61.8%) | 472 (61.2%) | |
| | III | 34 (20.0%) | 10 (15.6%) | 103 (19.2%) | 147 (19.1%) | |
| | IV | 1 (0.6%) | 1 (1.6%) | 4 (0.7%) | 6 (0.8%) | |
| | Not Reported | 29 (17.1%) | 8 (12.5%) | 75 (14.0%) | 112 (14.5%) | |
| Grade (Pathological) | I | 8 (4.7%) | 3 (4.8%) | 23 (4.3%) | 34 (4.4%) | 0.881 |
| | II | 97 (57.1%) | 41 (65.1%) | 330 (61.6%) | 468 (60.9%) | |
| | III | 32 (18.8%) | 9 (14.3%) | 102 (19.0%) | 143 (18.6%) | |
| | IV | 1 (0.6%) | 1 (1.6%) | 4 (0.7%) | 6 (0.8%) | |
| | Not Reported | 32 (18.8%) | 9 (14.3%) | 77 (14.4%) | 118 (15.3%) | |
| Stage (Clinical) | 0 | 2 (1.1%) | 1 (1.6%) | 2 (0.4%) | 5 (0.6%) | 0.618 |
| | I | 8 (4.5%) | 4 (6.3%) | 16 (3.0%) | 28 (3.6%) | |
| | II | 3 (1.7%) | 1 (1.6%) | 13 (2.4%) | 17 (2.2%) | |
| | III | 3 (1.7%) | 1 (1.6%) | 4 (0.7%) | 8 (1.0%) | |
| | IV | 29 (16.4%) | 6 (9.5%) | 86 (16.0%) | 121 (15.5%) | |
| | Not Reported | 132 (74.6%) | 50 (79.4%) | 418 (77.6%) | 600 (77.0%) | |
| Stage (Pathological) | 0 | | | 5 (0.9%) | 5 (0.6%) | <.001 |
| | I | 4 (2.3%) | 4 (6.3%) | 28 (5.2%) | 36 (4.6%) | |
| | II | 4 (2.3%) | 3 (4.8%) | 60 (11.1%) | 67 (8.6%) | |
| | III | 14 (8.0%) | 6 (9.5%) | 77 (14.3%) | 97 (12.5%) | |
| | IV | 22 (12.5%) | 5 (7.9%) | 76 (14.1%) | 103 (13.2%) | |
| | Not Reported | 132 (75.0%) | 45 (71.4%) | 293 (54.4%) | 470 (60.4%) | |
| Overall Survival Indicator | Alive | 45 (25.4%) | 21 (32.8%) | 201 (37.3%) | 267 (34.2%) | 0.015 |
| | Dead | 132 (74.6%) | 43 (67.2%) | 338 (62.7%) | 513 (65.8%) | |
| Progression-Free Survival Indicator | No Progression | 36 (20.3%) | 20 (31.3%) | 179 (33.2%) | 235 (30.1%) | 0.005 |
| | Progression | 141 (79.7%) | 44 (68.8%) | 360 (66.8%) | 545 (69.9%) | |

| Overall Survival (OS) | Median Surv. (95% CI) | Sample: Colon | Log Rank P-value |
|---|---|---|---|
| Total | 44.4 (39.2, 49.0) | E=513 C=267 T=780 | p= 0.005 |
| ALP | 57.7 (35.1, 85.3) | E=43 C=21 T=64 | |
| LOR | 43.0 (31.1, 50.3) | E=132 C=45 T=177 | |
| No Benzo | 43.9 (38.9, 50.0) | E=338 C=201 T=539 | |

| Progression-Free Survival (PFS) | Median Surv. (95% CI) | Sample: Colon | Log Rank P-value |
|---|---|---|---|
| Total | 31.0 (27.8, 33.6) | E=545 C=235 T=780 | p= <.001 |
| ALP | 38.1 (22.3, 59.8) | E=44 C=20 T=64 | |
| LOR | 23.8 (20.2, 30.5) | E=141 C=36 T=177 | |
| No Benzo | 33.1 (29.2, 38.1) | E=360 C=179 T=539 | |

| **Overall Survival (OS)** | | |
|---|---|---|
| **Predictors: Cohort, Sex, Clinical Grade, and Clinical Stage** | | |
| **Cohort** | **HR (95% CI)** | **P-value** |
| No Benzo | Ref. | <.0001 |
| ALP | 1.369 (0.988 – 1.897) | 0.0593 |
| LOR | 1.620 (1.317 – 1.993) | <.0001 |
| **Progression-Free Survival (PFS)** | | |
| No Benzo | Ref. | <.0001 |
| ALP | 1.290 (0.936 – 1.778) | 0.1200 |
| LOR | 1.782 (1.457 – 2.179) | <.0001 |

E=event, C=censored, T=Total

### Table 13. Prostate cancer patient characteristics, median survival, and Multivariate Cox Regression Modeling

| Patient Characteristics: Prostate | | LOR | ALP | No Benzo | Overall | P-value |
|---|---|---|---|---|---|---|
| | N | 175 (21.3) | 118 (14.4) | 528 (64.3) | 821 (100%) | |
| Sex | Male | 175 (100.0%) | 118 (100.0%) | 528 (100.0%) | 821 (100.0%) | |
| Grade (Clinical) | I | 14 (8.2%) | 2 (1.7%) | 10 (1.9%) | 26 (3.2%) | <.001 |
| | II | 61 (35.7%) | 35 (30.2%) | 212 (40.8%) | 308 (38.2%) | |
| | III | 78 (45.6%) | 73 (62.9%) | 282 (54.2%) | 433 (53.7%) | |
| | IV | 3 (1.8%) | 1 (0.9%) | 8 (1.5%) | 12 (1.5%) | |
| | Not Reported | 15 (8.8%) | 5 (4.3%) | 8 (1.5%) | 28 (3.5%) | |
| Grade (Pathological) | I | 9 (5.2%) | 1 (0.9%) | | 10 (1.2%) | <.001 |
| | II | 53 (30.8%) | 33 (28.2%) | 204 (38.9%) | 290 (35.7%) | |
| | III | 75 (43.6%) | 73 (62.4%) | 277 (52.9%) | 425 (52.3%) | |
| | IV | 1 (0.6%) | | 1 (0.2%) | 2 (0.2%) | |
| | Not Reported | 34 (19.8%) | 10 (8.5%) | 42 (8.0%) | 86 (10.6%) | |
| Stage (Clinical) | 0 | | | 1 (0.2%) | 1 (0.1%) | <.001 |
| | I | 17 (9.8%) | 11 (9.5%) | 5 (0.9%) | 33 (4.0%) | |
| | II | 61 (35.1%) | 46 (39.7%) | 414 (78.4%) | 521 (63.7%) | |
| | III | 2 (1.1%) | 1 (0.9%) | 8 (1.5%) | 11 (1.3%) | |
| | IV | 25 (14.4%) | 10 (8.6%) | 19 (3.6%) | 54 (6.6%) | |
| | Not Reported | 69 (39.7%) | 48 (41.4%) | 81 (15.3%) | 198 (24.2%) | |
| Stage (Pathological) | I | | | 2 (0.4%) | 2 (0.2%) | <.001 |
| | II | 21 (12.1%) | 20 (17.1%) | 161 (30.5%) | 202 (24.7%) | |
| | III | 11 (6.3%) | 7 (6.0%) | 44 (8.3%) | 62 (7.6%) | |
| | IV | 5 (2.9%) | 4 (3.4%) | 14 (2.7%) | 23 (2.8%) | |
| | Not Reported | 137 (78.7%) | 86 (73.5%) | 307 (58.1%) | 530 (64.7%) | |
| Overall Survival Indicator | Alive | 98 (56.0%) | 68 (57.6%) | 383 (72.5%) | 549 (66.9%) | <.001 |
| | Dead | 77 (44.0%) | 50 (42.4%) | 145 (27.5%) | 272 (33.1%) | |
| Progression-Free Survival Indicator | No Progression | 87 (49.7%) | 63 (53.4%) | 339 (64.2%) | 489 (59.6%) | 0.001 |
| | Progression | 88 (50.3%) | 55 (46.6%) | 189 (35.8%) | 332 (40.4%) | |

| Overall Survival (OS) | Median Surv. (95% CI) | Sample: Prostate | Log Rank P-value |
|---|---|---|---|
| Total | 202.2 (188.0, NR) | E=272 C=549 T=821 | p= <.001 |
| ALP | 155.6 (117.6, 202.2) | E=50 C=68 T=118 | |
| LOR | 116.5 (82.2, 136.6) | E=77 C=98 T=175 | |
| No Benzo | NR (200.1, NR) | E=145 C=383 T=528 | |

| Progression-Free Survival (PFS) | Median Surv. (95% CI) | Sample: Prostate | Log Rank P-value |
|---|---|---|---|
| Total | 157.3 (146.7, 184.4) | E=332 C=489 T=821 | p= <.001 |
| ALP | 127.5 (107.0, 181.1) | E=55 C=63 T=118 | |
| LOR | 90.6 (67.4, 116.7) | E=88 C=87 T=175 | |
| No Benzo | 197.0 (174.1, NR) | E=189 C=339 T=528 | |

| Overall Survival (OS) | | |
|---|---|---|
| **Predictors: Cohort, Clinical Grade, and Clinical Stage** | | |
| **Cohort** | **HR (95% CI)** | **P-value** |
| No Benzo | Ref. | <.0001 |
| ALP | 1.464 (1.038 – 2.064) | 0.0298 |
| LOR | 2.160 (1.589 – 2.936) | <.0001 |
| **Progression-Free Survival (PFS)** | | |
| No Benzo | Ref. | <.0001 |
| ALP | 1.237 (0.899 – 1.702) | 0.1916 |
| LOR | 1.899 (1.433 – 2.517) | <.0001 |

E=event, C=censored, T=Total
